# Benign-Ex: Delineating Regions of the Human Genome Benign to Copy Number Variation

**DOI:** 10.1101/2022.10.17.22280252

**Authors:** Alyssa S. Wetzel, Heather Major, Mrutyunjaya Parida, J. Robert Manak, Benjamin W. Darbro

## Abstract

While copy number variants (CNVs) have been identified as an important cause of rare genetic disorders, they have also been identified in unaffected control populations, making clinical interpretation of these lesions challenging. Discriminating benign CNVs from those pathogenic for rare genetic disorders, therefore, relies on understanding what regions of the human genome are tolerant to copy number variation. Benign-Ex is a python-based program that uses information from databases of CNVs to generate one or more benign interval map(s) and then identifies the optimal map by computing the overlap with known pathogenic regions. We utilized Benign-Ex to identify the optimal set of benign intervals from two distinct CNV databases: Database of Genomic Variants (DGV) and Clinical Genome Resource (ClinGen). Benign-Ex called 41.1% of the genome benign using data from DGV and 37.6% of the genome benign using data from ClinGen. The benign regions from DGV and ClinGen were *not* spatially correlated, underscoring the importance of integrating both research and clinical databases for determining CNV benignity.

## Introduction

Copy number variants (CNVs) are a class of unbalanced structural variation that alters the physical architecture of the human genome and results in a net gain (duplication) or loss (deletion) of genetic material. While CNVs have long been known to be associated with human disorders, until 2004, it was presumed that with few exceptions, CNVs always confer a phenotypic effect ^1^. The reports by Iafrate *et al*. ^*2*^ and Sebat *et al*. ^*3*^, which identified on average 11-12 CNVs per individual in unrelated and healthy populations, drastically changed our understanding of CNVs. Since then, over seventy-five papers have been published investigating structural variation in the human genome, and these variants are cataloged in the Database of Genomic Variants (DGV) ^4^. At the same time, chromosomal microarrays became the first-line diagnostic test in the evaluation of patients with non-syndromic developmental delay, intellectual disability, autism spectrum disorders, and multiple congenital anomalies not specific to a well-delineated genetic syndrome ^1,5-9^; thus further increasing our knowledge about CNVs in clinical populations. Together, this has led to an increasing need to classify CNVs better according to potential pathogenicity.

Historically, the classification of the pathogenicity of CNVs has been a manual and subjective process that relies heavily on the size of the CNV, the genes present within the CNV interval, what those genes are known or believed to do, and the phenotype of the patient. Recurrent CNVs are easier to interpret as, by definition, they have been observed before in patient populations with similar clinical phenotypes (pathogenic) or at a high frequency (>1%) in control populations or within in-house laboratory databases (benign). In addition, the American College of Medical Genetics and Genomics (ACMGG) and the Clinical Genome Resource (ClinGen) have established criteria that allow rare non-recurrent CNVs to be interpreted as pathogenic, including multiple independent reports of similarly sized CNVs in patients with overlapping phenotypic features, an inheritance pattern within families that supports segregation of the CNV with affected status, and gene-phenotype correlations ^10-12^. In contrast, benign CNVs are only classified as such if they overlap with known copy number polymorphic sites or the CNV has significant peer-reviewed evidence indicating the variation is benign. Further, there is no “gold-standard” set of benign CNVs clinicians and researchers can use when evaluating rare CNVs. Recently, the ACMGG and ClinGen updated their classification guidelines to include quantitative thresholds for classification and recommended uncoupling pathogenicity classifications and direct clinical relevance. Therefore, delineating regions of the human genome which are benign to copy number variation is imperative to accurately and consistently assign benign pathogenicity classifications.

To answer the question of what the most reliable set of ‘benign’ regions is, we must first determine how to best leverage both clinical and population-level databases. Here we report on the development and performance of a new computational program – Benign-Ex – which was created to address the limitations in non-phenotype driven benign CNV discrimination/classification. Unlike prior estimates of benign copy number variation in the human genome, Benign-Ex (1) incorporates clinically curated benign and pathogenic CNV data from ClinGen (2) assesses and minimizes the overlap between “benign” called regions and known microduplication and microdeletion syndromes (MMS), and (3) provides an automated, high-throughput, and customizable system which allows the end-user to choose both the level of evidence required for a region to be called “benign” and test the performance against any end-user created “pathogenic” interval list (e.g. known microdeletion/duplication syndromes or MMS). Notably, Benign-Ex takes pathogenic information into account and uses this information to penalize regions and/or maps that call known pathogenic regions benign. Finally, Benign-Ex outputs the identified “benign” regions as a BED file which can be used to aid in the real-time identification of benign CNVs.

## Methods

### Overview of Benign-Ex

Benign-Ex utilizes data from either frequency-based or clinical classification-based datasets to identify regions of the human genome that are copy number variable in control populations (“benign”). Its goal is to maximize the amount of the genome called benign, while simultaneously minimizing the overlap of benign regions with known pathogenic regions (*e*.*g*. MMS). For our purposes, we leveraged both the Database of Genomic Variants (DGV) and the Clinical Genome Resource (ClinGen), however, so long as the datasets meet the requirements of the input data, alternative datasets such as dbVar and/or clinical laboratory databases of pathogenicity classifications are supported. To account for the breadth of variants and potential discrepancies in pathogenicity classifications, Benign-Ex implements a set of filtering criteria, allowing the user to customize which regions of the genome are considered benign separately for duplications and deletions, and frequency-based and classification-based databases. In addition, Benign-Ex has the built-in flexibility to incorporate and assess multiple sets of filtering criteria at once.

Genome-wide structural variation data was collected from the Clinical Genome Resource (ClinGen; www.clinicalgenome.org) and Database of Genomic Variants (DGV) ^4,13^. Data from DGV was downloaded for the 2020-02-25 release of DGV to obtain a total of 826,661 (hg19), and 827,036 (GRCh38) CNV intervals. Data from ClinGen was downloaded via the UCSC Table Browser for the 2020-05-14 release of ClinGen to obtain a total of 27,963 (hg19), and 30,560 (GRCh38) CNVs annotated as Benign (iscaBenign), Likely Benign (iscaLikelyBenign), or Uncertain Significance (iscaUncertain) ^14,15^.

DGV is a curated database that contains structural variants (*e*.*g*. deletions, duplications, and inversions) found within control populations, and its primary goal is to catalog normal structural variation in the human genome^4^. As the individual variants within DGV cover >86% of the human genome^4^, including regions known to be associated with human disease (e.g. 22q11.2), Benign-Ex leverages information contained within the data set to filter the individual variants. The user specifies the filtering parameters which include: (a) the minimum number of independent CNV events in the same direction (*i*.*e*. gain or loss; “qualifying variants”) (b) the minimum sample size of the study in which a CNV event was discovered, (c) the directionality of the CNV event (*i*.*e*. gain or loss), (d) the minimum frequency of the CNV event within the study in which it was discovered, (e) if only CNVs detected by specific technologies (BAC aCGH, SNP array, Oligo aCGH and/or Sequencing) should be considered, and/or (f) the year in which the study was published. Therefore, the user could implement a set of filtering criteria for DGV which calls a region of the human genome benign when duplicated <u>*if and only if*</u> three or more overlapping GAIN variants are found, where each individual GAIN variant is found within a study at a minimum frequency of 0.1%, and that study was published in or after 2009 with a minimum sample size of 100 individuals.

ClinGen is a curated database that contains structural variants obtained from clinical populations, and its primary goal is to standardize the interpretation of clinically identified structural variants. To account for discrepancies in CNV pathogenicity classifications within the ClinGen database ^11^, the user supplies the following filtering criteria: (a) the directionality of the CNV event (*i*.*e*. gain or loss), (b) the minimum number of individual CNV events in the same direction (qualifying variants), and (c) the acceptable CNV classifications to consider (*e*.*g*. benign, uncertain significance, likely benign). Thus, the user could implement a set of filtering criteria that calls a region of the genome benign when deleted only if two or more overlapping LOSS variants are found within ClinGen, and the pathogenicity classifications of those individual variants are “benign” or “likely benign”.

The set of known pathogenic regions used in Benign-Ex to evaluate the performance of Benign-Ex’s called benign regions was identified from the following databases and papers: DatabasE of genomiC varIation and Phenotype in Humans using Ensembl Resources (DECIPHER; https://decipher.sanger.ac.uk/) ^16^, Clinical Genome Resource (ClinGen; www.clinicalgenome.org) ^13^, Online Mendelian Inheritance in Man (OMIM; www.omim.org) ^17^, Table 1 of Wetzel and Darbro (in press) ^18^, Table 1 of Wiese *et al*. (2012) ^19^, Supplemental Table 7 of Cooper *et al*. (2011) ^20^ and Datasets S1 and S2 of Petrovski *et al*. (2013) ^21^ (**Supplementary Methods**).

**Table 1:** Summary of Benign-Ex Identified Benign Regions in the Human Genome (HG19)

|  | DGV |  |  | ClinGen |  |  | DGV + ClinGen |  |  |
| --- | --- | --- | --- | --- | --- | --- | --- | --- | --- |
|  | GAIN | LOSS | BOTH | GAIN | LOSS | BOTH | GAIN | LOSS | BOTH |
| Genome Coverage (%) | 29.6 | 24.5 | 41.1 | 30.8 | 14.8 | 37.6 | 46.3 | 32.0 | 58.0 |
| Genome Coverage (Mb) | 916.4 | 758.2 | 1,273.8 | 954.0 | 456.7 | 1,164.8 | 1,433.8 | 992.0 | 1,796.8 |
| Total Number of Benign Regions | 20,785 | 62,599 | 56,901 | 1,331 | 1,118 | 1,782 | 16,193 | 56,602 | 42,047 |
| Benign Region Length (Range) | 1 bp<br>8.5 Mb | 1 bp<br>8.5 Mb | 1 bp<br>8.5 Mb | 50 bp<br>80.6 Mb | 82 bp<br>35.6 Mb | 50 bp<br>87.7 Mb | 1 bp<br>91.2 Mb | 1 bp<br>35.6 Mb | 1 bp<br>91.2 Mb |
| Median Benign Region Length (kb) | 3.00±1.25 | 1.21±0.31 | 1.40±0.53 | 225.8±96.1 | 174.6±47.2 | 212.1±76.1 | 3.32±8.60 | 1.22±1.01 | 1.44±3.4 |

To identify benign regions, Benign-Ex interrogates the genome in 500-bp windows to determine if any CNVs from the specified database (*e*.*g*. DGV or ClinGen) meet the filtering criteria (**Supplementary Methods**). If the number of variants that meet the filtering criteria meets or exceeds the minimum number of qualifying variants, then Benign-Ex will designate the overlapping region of the qualifying variants as “benign” (**Figure 1**). After identifying the qualifying benign regions which meet the user’s filtering criteria, Benign-Ex will merge benign regions which are within 500bp of one another.

**Figure 1:**
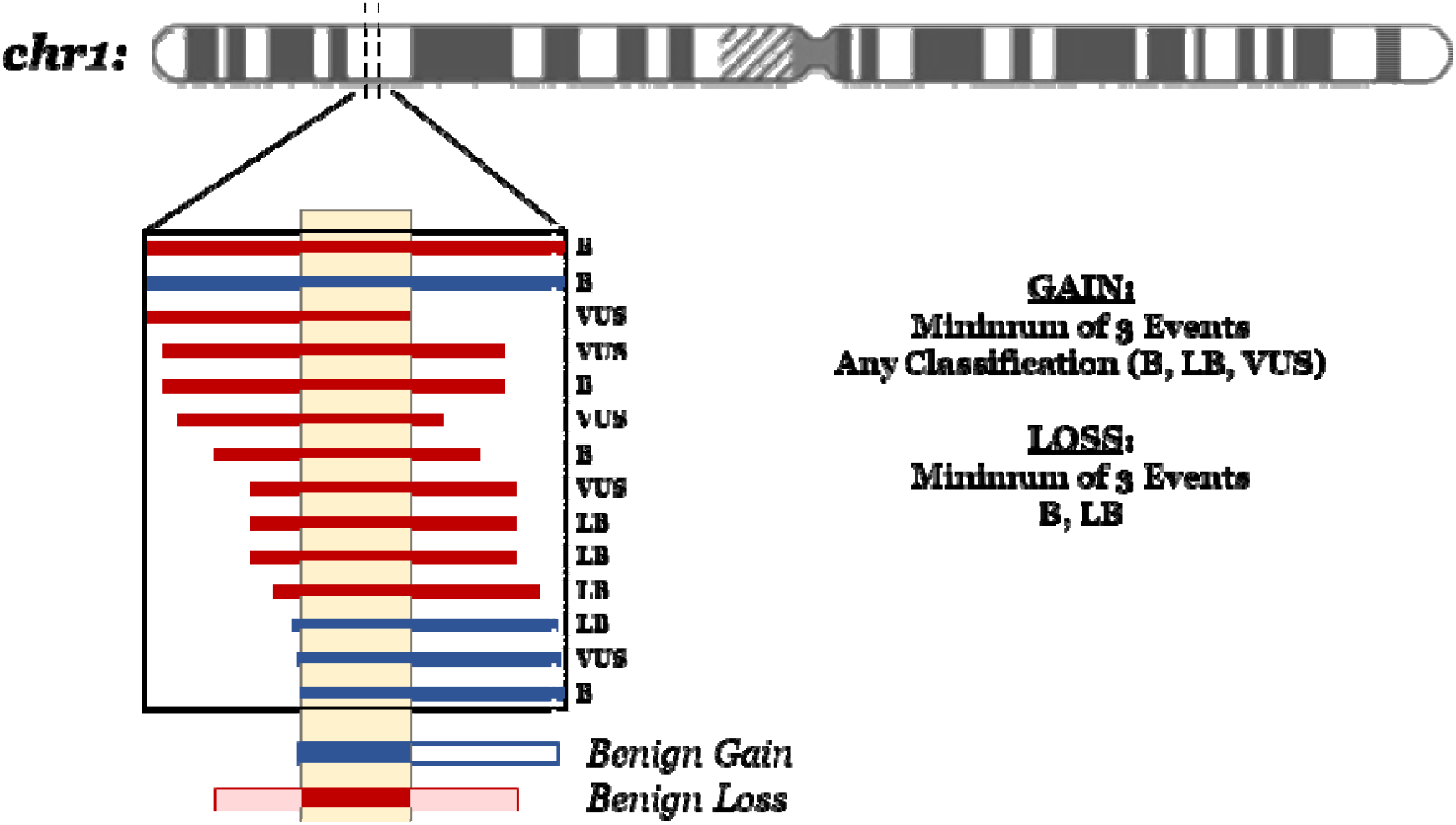
Identifying Benign Regions of the Human Genome via Benign-Ex. Benign-Ex interrogates the genome in 500-bp windows to determine if any CNVs from the specified database meet the filtering cr teria. In this example, both the GAIN and LOSS filtering criteria requires three or more qualifying variants however, the acceptable classifications differ between GAIN (benign, likely benign, or variant of unknown significance) and LOSS (benign or likely benign). The yellow region represents the 500-bp window Benign-Ex is currently evaluating. Within this window, all four GAIN variants meet the filtering criteria, while six of the ten LOSS variants meet the filtering criteria. Therefore, this entire 500-bp window is ‘benign’ to both gains and losses. The lighter blue and red regions which expand beyond the current 500-bp window represent the final merged benign intervals. Blue variants represent gains, while red variants represent losses. The classification of each individual variant is listed on the left: benign (B), likely benign (LB), variant of unknown significance (VUS). NOTE: Depiction for illustrative purposes, not drawn to scale or representative of actual data.

### Benign-Ex Optimization

Optionally, users can supply a series of different filtering criteria to generate multiple sets of benign intervals. The relative performance of each set of benign regions can be then compared by calculating the dissimilarity between each benign interval list and a set of known pathogenic intervals (**Supplementary Methods**).

To identify the optimal set of criteria for calling benign regions via Benign-Ex for both ClinGen and DGV (against a specific set of lists with pathogenic loci), we established a set of simulation values for each parameter of the inclusion criteria (**Supplemental Table 1**). For DGV, we performed simulations with respect to the direction of the CNV event for the minimum number of qualifying events and minimum within study allele frequency with and without year and methodology filters applied, for a total of 576 unique parameter combinations (288 gain; 288 loss). For ClinGen, the minimum number of qualifying variants and the acceptable classification(s) were varied with respect to the direction of the CNV event for a total of 30 unique parameter combinations (15 gain; 15 loss).

### Evaluation of Optimal Benign Regions

The most recent commonly accepted map of benign copy number variation in the human genome was published by Zarrei *et al*. ^22^ in 2015 and used the 2013-07-23 release of DGV. Their inclusive map required each individual variant to be supported by a minimum of two subjects, while the stringent map required each individual variant to be supported by a minimum of two subjects and two different studies. The hg19 versions of the stringent and inclusive maps were downloaded from DGV’s website. The reldist and fisher functions from the Bedtools suite ^23^ were used to evaluate the spatial correlation between benign interval sets (**Supplementary Methods**).

### Statistical Methods

Statistical analyses (Kruskal-Wallis one-way-analysis of variance and Wilcoxon Rank Sum test) were performed with R v4.1.2.

## Results

### Benign-Ex Optimization

The optimal parameter combination was the set of benign intervals that had the best average performance across all pathogenic lists tested. We tested a total of 2560 parameter combinations during our preliminary testing to confirm the functionality of Benign-Ex and ensure that the optimal parameter setting is dependent on the set of user-supplied parameters (**Supplementary Methods; Supplemental Table 2**). To determine the optimal set of benign intervals for both the Database of Genomic Variants (DGV) and the Clinical Genome Resource (ClinGen), we focused on a subset of 606 parameter settings (**Supplemental Table 1**).

For DGV, Benign-Ex chose 2_30_0.004_N_2009 (gain) and 2_30_0.006_N_2009 (loss) as the optimal parameter sets (**Figure 2; Supplemental Figure 1**). Therefore, Benign-Ex requires a minimum of two variants with an allele frequency (AF) greater than 0.004% (Gain Events) or 0.006% (Loss Events). Each of these 2+ variants must also come from a study published in or after 2009 with a sample size greater than 30.

**Figure 2:**
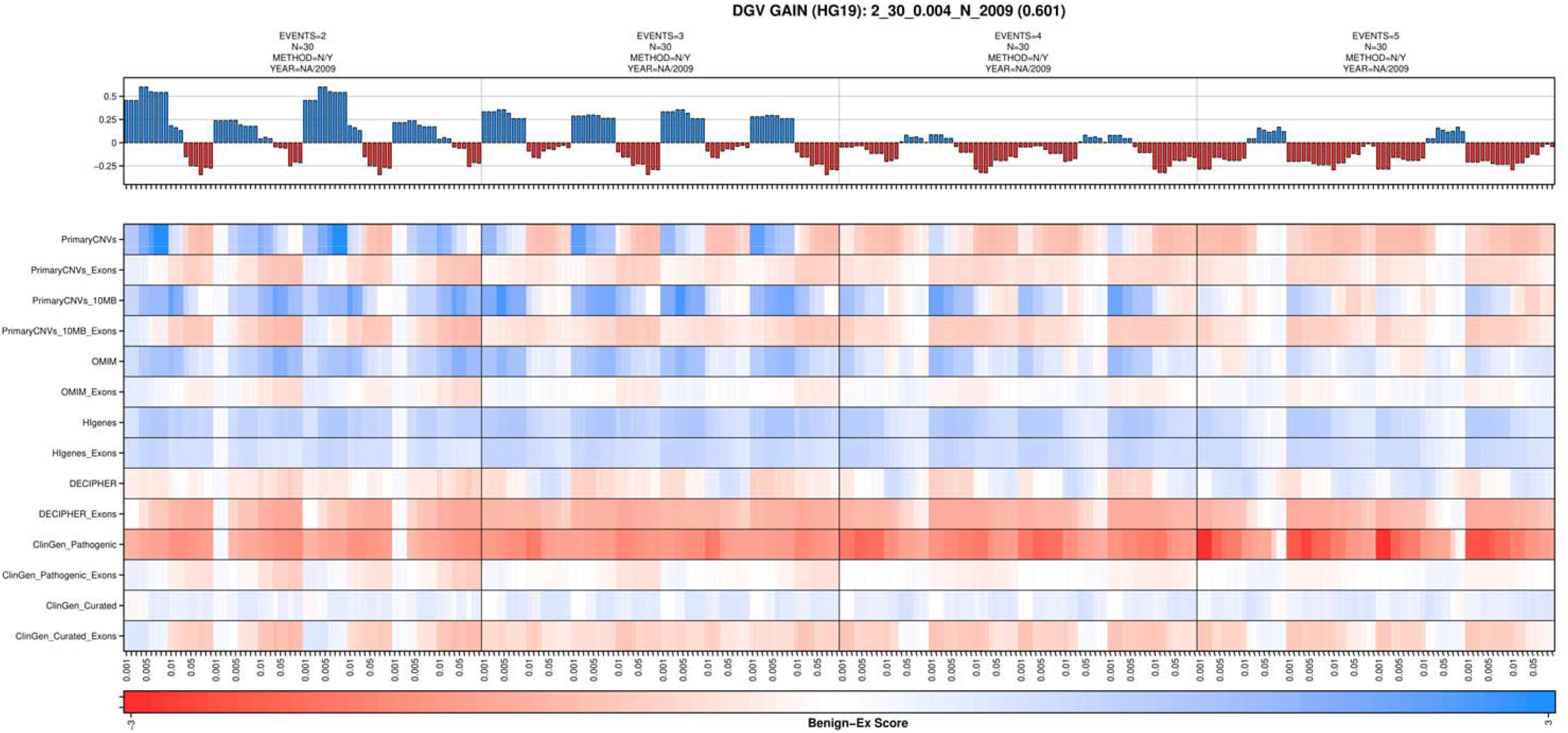
Benign-Ex Heatmap for DGV and Duplications. To visually demonstrate how different Benign-Ex parameter sets perform across multiple pathogenic lists, Benign-Ex produces a multi-panel heatmap. Top: The histogram represents the average performance of that parameter set across all pathogenic lists. The maximum value is the optimal parameter setting across all pathogenic lists. Middle: The heatmap plots the final summary statistic for each pathogenic list and Benign-Ex parameter set combination. The Benign-Ex parameter simulations are sorted first by the minimum number of events, then left to right by the combination of the year and methdology filter applications: (N/2009, N/NA, Y/2009, Y/NA). Finally, the simulations are sorted by allele frequency (AF) which are indicated. Bottom: Color scale for the final summary statistic value. Negative scores are in red, positive scores are in blue, and ‘0’ scores are white.

For ClinGen, Benign-Ex chose 1_B_USLB (gain) and 1_B_USLB (loss) as the optimal parameter sets (**Supplemental Figure 2**). Therefore, Benign-Ex only requires a single variant classified as benign or likely benign. Despite the minimum number of events being set at one, the Benign-Ex did *not* choose the most lenient criteria possible which would have also allowed for the inclusion of variants of unknown significance.

### Benign-Ex Benign Region Characteristics

Individually, Benign-Ex called 41.1% of the genome benign using data from DGV (29.6% gain; 24.5% loss) and 37.6% of the genome benign using data from ClinGen (30.8% gain; 14.8% loss; **Table 1**; **Figure 3**). A total of 10,699 regions (17.8% genome) were called benign by both DGV and ClinGen in both directions. Individually, 5,923 regions (14.1% genome) and 7,115 regions (7.2% genome) were called benign by both DGV and ClinGen for gain and loss events, respectively. After merging the benign regions from DGV and ClinGen, Benign-Ex called 46.3% of the genome benign with respect to gain events, 32.0% of the genome benign with respect to loss events, and 58.0% overall.

**Figure 3:**
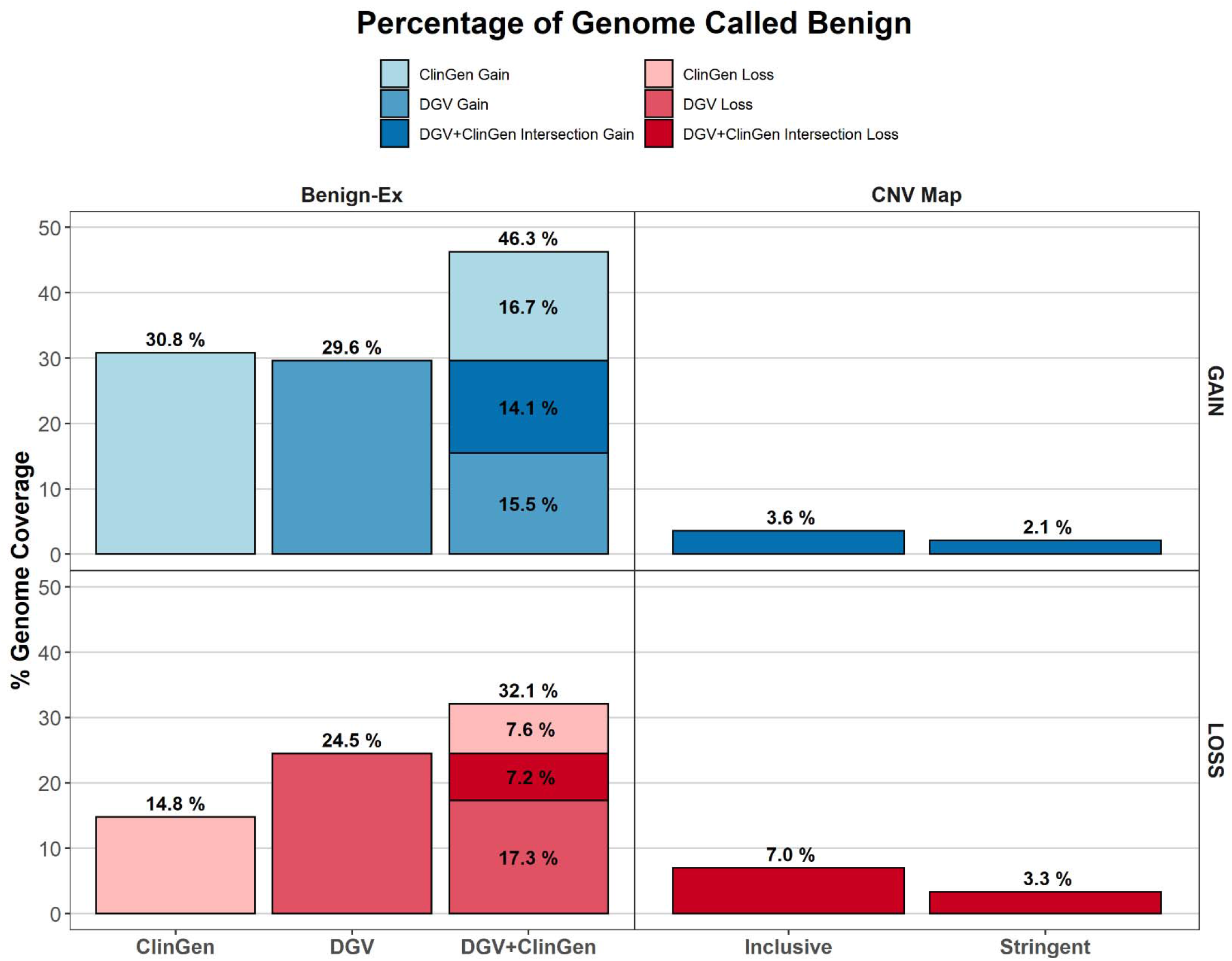
Percent of the Human Genome Called “Benign” to Copy Number Variants. The percent of the human genome that was found to be copy number variable in control population databases and thus “benign” to copy number variation. Stringent and Inclusive CNV Map percentages were obtained from Zarrei *et al*.^22^. Benign-Ex percentages for ClinGen and DGV individually, as well as the merged interval set are shown. The dark blue (gain) and dark red (loss) blocks represent the intersection subset wherein both DGV and ClinGen called the region benign.

A total of 85,833 individual regions of the genome (16,193 distinct gain regions and 56,602 distinct loss regions) were called benign using the merged DGV and ClinGen benign regions (**Supplemental Figure 3C**). DGV was the larger contributor to these merged regions with nearly all the merged regions being covered by DGV benign regions (Gain: 98.9%; Loss: 99.9%) and <10% of the merged regions covered by the ClinGen benign regions (Gain: 7.37%; Loss: 1.81%; **Supplemental Figure 3D**). Interestingly, the benign regions from DGV and ClinGen were *not* spatially correlated (**Figure 4**; **Supplemental Table 1**), indicating that the combination of the two datasets is informative and provides unique regions (bedtools fisher: p=1; **Supplementary Methods**) ^23^.

**Figure 4:**
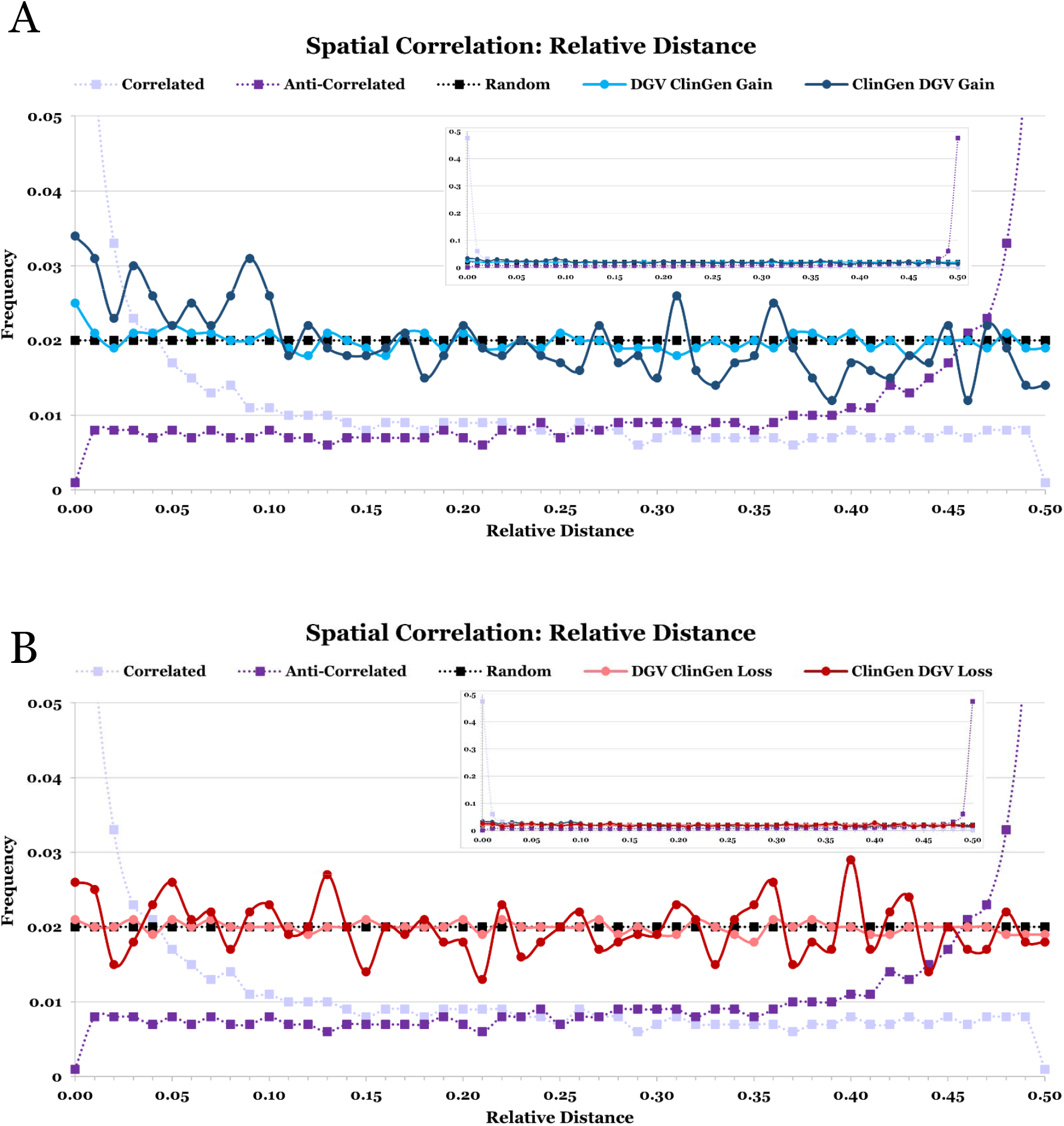
Spatial Correlation Between the Benign-Ex’s DGV and ClinGen Benign Regions. Spline graph depicting the spatial correlation between Benign-Ex’s DGV and ClinGen benign regions as assessed by the Bedtools reldist function for (A) gains and (B) losses. For each graph, both reciprocal comparisons are plotted, and the inset shows the full view of the expected relative distance distributions for correlated and anti-correlated interval sets. Light Purple: Correlated interval sets; Dark Purple: Anti-correlated interval sets; Black: Random unrelated interval sets.

The length of duplication benign regions was significantly longer than deletion benign regions (**Table 1**; p<0.001) for both DGV (Duplication: 3.00±1.25 kb; Deletion: 1.21±0.31 kb; **Supplemental Figure 3A**) and ClinGen (Duplication: 225.8±96.1 kb; Deletion: 174.6±47.2 kb; **Supplemental Figure 3B**). Further, the median length of benign regions was significantly smaller for DGV than ClinGen for both duplication and deletions (p<0.001; **Table 1**). Duplication benign regions were significantly larger than the deletion benign regions for the merged set of intervals as well (p<0.001; **Supplemental Figure 3C**).

The merged benign regions are not evenly distributed across the genome (**Supplemental Figure 5**) and there is greater variability between chromosomes for the set of benign regions identified by the ClinGen data (Gain: 15.8-96.9%; Loss: 4.0-72.1%) than DGV (Gain: 19.6-39.6%; Loss: 8.1-33.0%). For the ClinGen dataset, the Y chromosome had the highest percentage of benign region coverage (Gain: 96.9%; Loss: 72.1%), while chromosome 14 was on the low end of benign region coverage (Gain: 16.1%; Loss: 8.5%). For the DGV dataset, the Y chromosome had the *lowest* percentage of benign region coverage (Gain: 19.6%; Loss: 8.1%), and chromosomes 7, 9, and 16 have the highest percentage of benign region coverage (Gain: 37.1-.39.6%; Loss: 31.9-33.0%). Except for chromosomes 3 (DGV) and 15 (ClinGen), each chromosome had a higher percentage of coverage for benign gain regions than benign loss regions.

As the Stringent and Inclusive CNV Maps were derived from the same DGV data with different thresholds for variant inclusion, we would expect to observe a high spatial correlation between the two interval sets. In fact, because the Stringent Map is entirely contained within the Inclusive Map, the Overlap Coefficient (for both gains and losses) is 1.0 (**Supplemental Table 3**). This high spatial correlation can also be observed with the Bedtools suite tools reldist and fisher ^23^ (**Supplemental Figure 6**).

The Stringent and Inclusive CNV Maps contain less genomic content than the Benign-Ex intervals derived from the new DGV dataset (**Supplemental Table 4**). Therefore, the Overlap Coefficient can be used as a measure to determine how much the new Benign-Ex benign region intervals spatially overlap the old CNV Map intervals. While the Overlap Coefficient values are higher for the Stringent CNV Map (Gain: 0.963; Loss: 0.941) than the Inclusive CNV Map (Gain: 0.878; Loss: 0.810), the reldist and fisher exact tests support a high spatial correlation between all benign region interval sets (**Supplemental Table 4; Supplemental Figure 7**).

Furthermore, most of the intervals in the Stringent and Inclusive CNV Maps (85.2-97.5%; **Supplemental Table 5**) were overlapped by at least one Benign-Ex benign interval, and a small percentage (1.3-3.7%) were overlapped by two or more Benign-Ex benign regions (**Supplementary Methods**). There was a statistically significant difference in benign region length by direction when comparing Benign-Ex, Stringent, and Inclusive Maps for all intervals (Gain: χ^2^=489.2, p<0.001; Loss: χ^2^ =149.95, p<0.001), and just those regions which overlap (Gain: χ^2^=242.14, p<0.001; Loss: χ^2^ =143.79, p<0.001). However, the benign region length was *not* significantly different between the Benign-Ex Map and Stringent CNV Map for either gains or losses when looking solely at the overlapping intervals (Gain: p=0.27; Loss: p=0.59). This suggests that the Benign-Ex set of benign intervals not only overlaps that of the Stringent CNV Map but that these intervals are similarly sized (**Supplemental Figure 8**). Thus, the Benign-Ex map is not only re-identifying the same regions found by Zarrei *et al*. but also identifying novel regions (**Figure 3**).

## Discussion

The first map of human copy number variation came out shortly after the discovery that CNVs are present in the genomes of control individuals. Utilizing the HapMap population, Redon *et al*. ^24^ determined that 12% of the human genome was copy number variable. Conrad *et al*. ^25^ expanded on this analysis, using high-resolution comparative genomic hybridization arrays to perform fine-scale mapping of copy number variation in the HapMap population. By their estimates, they were able to ascertain copy number polymorphisms >500bp with minor allele frequency (MAF) greater than 5%. More recently, Zarrei *et al*. ^22^ leveraged the entire DGV database to create a map of copy number variation and identified over 100 genes that could be deleted without apparent phenotypic consequence. Since this last iteration of a human genome CNV map, DGV has increased 3-fold, and databases for clinical CNVs with pathogenicity classifications have been established. To date, over seven million unique CNVs which cover 86.14% of the genome have been found in presumably-healthy control individuals ^4^, and together the ClinGen and ClinVar databases contain over 100,000 unique clinically ascertained CNVs ^14,15,26-28^. We ran over two thousand different simulations against a series of known pathogenic region interval sets to identify which set of parameter settings optimally calls benign regions from DGV and ClinGen data. We determined that the intersection of these regions called 17.8% of the human genome benign (14.1% Gain; 7.2% Loss) while the union of these regions called 58.0% of the human genome benign (46.3% Gain; 32.0% Loss). While the percentage of the human genome that was called benign via the intersection interval set is more in line with prior estimates of benign coverage ^22,24^, we focused on the union interval set. The lack of spatial correlation between the DGV and ClinGen datasets underscores the importance of integrating clinical data in the assessment of benignity.

This large increase in the genome coverage from the merged interval track while unexpected, can be explained by both the large disparity between coding and non-coding regions, and the increase in the amount and type of data that was utilized to generate the benign maps. Given that an estimated 98% of the human genome is non-coding and known recurrent MMS account for only 25% of the human genome, it isn’t unreasonable for a large percentage of the human genome to be benign with respect to copy number variation. Further, there was a four-fold increase in the size of the DGV database, and the addition of clinically ascertained benign variants.

The greater variability in the inter-chromosomal benign region percentages identified by Benign-Ex for the ClinGen database relative to the DGV database (**Supplemental Figure 5**) is likely related to the sizes and nature of the databases themselves. While the ClinGen database is 29.5 times smaller than the DGV database, because the database consists of curated variants, we have greater confidence in the benign nature of individual variants within ClinGen, despite there being fewer variants overall. However, due to its relative size and the inclusion of all benign and likely benign variants by Benign-Ex, it is less robust against effects from individual variants and their distribution across the genome. Similarly, the observed size disparities between the benign regions from the DGV and ClinGen datasets (**Supplemental Figure 3**) can likely be attributed to methodological differences. A large proportion of the most recent data within DGV used sequencing technologies, while the clinical data from ClinGen continued to be primarily ascertained via array-based methodologies. Therefore, the more precise breakpoints and subsequently smaller CNV intervals within the DGV database are reflected in the average benign interval length.

Clinically, the evaluation of individual copy number variants is a limiting step in the efficiency and speed of chromosomal microarray testing. Each variant requires manual review to determine if it is real or an artifact and to determine the pathogenicity. With the ACMGG’s new evaluation criteria, complete overlap with an established benign region (Table 2: Section 2C-D,F [gain]; Table 1: Section 2F [loss]) ^29,30^ is sufficient evidence to call a clinically ascertained CNV benign. Therefore, these benign regions can be used clinically, as part of the manual review process, or in classification algorithms like DISCRIMINATOR (manuscript in preparation) ^31^ which prioritize assessment of pathogenicity over benignity.

Benign-Ex has several built-in functions which allow it to be of particular use in this regard. First, it is configured to easily allow for the generation of new benign interval maps via parameter simulations with the release of new database versions. This in turn allows for the continual refinement of benign regions. Secondly, while the curated nature of the ClinGen database means that we have greater confidence in the benign nature of any individual variant, through UCSC Table Browser we are unable to ascertain the date on which the variant was submitted or determine if the variant classification has been reviewed. Should this information become available for classification-based variants, Benign-Ex is uniquely poised to be able to implement additional filters to allow further refinement of benign intervals and assess the effects of initial classification and/or review date.

A major limitation in the identification of benign regions is the lack of a current gold standard. Without concrete knowledge about which regions of the human genome tolerate gains and losses without phenotypic effect, the interpretation of clinical CNVs remains difficult, contributing to the ever-increasing number of VUS. While the regions identified by Benign-Ex are spatially correlated with those identified by Zarrei *et al*. ^22^, and there is a large-scale pattern of benign region reproduction (**Supplemental Table 4; Supplemental Figure 7**), how small-scale differences in benign intervals manifest themselves on the large-scale was not investigated in the current work. We would expect to see that with the increase in the size of the DGV database, the benign intervals would collapse into more precise intervals. However, it is unclear if this collapsing effect is occurring and/or how these ‘new’ Benign-Ex intervals are centered on the original intervals from the Stringent and Inclusive maps.

In addition to the evaluation of different benign region maps, the ability to do these small-scale comparisons would allow for a better evaluation of benign region map overlap with known pathogenic regions. At this time, an ultra-conservative estimate for a new ‘gold-standard’ of benign regions could be gathered by taking the intersection of Zarrei *et al*.’s Stringent Map, and the Benign-Ex intervals for DGV and ClinGen and perhaps further refined by removing regions of pathogenicity such as known MMS. Within our own institution, such BED file tracks have been uploaded to our clinical CNV detection and interpretation software (NxClinical; BioDiscovery Inc., El Segundo, CA) to aid in the real-time classification of patient CNVs.

## Data Availability

Benign-Ex is publicly available on GitHub at https://github.com/aswetzel/Benign-Ex. Full simulation data is available upon request. All other data produced and analyzed in the present study are included in this published article and its supplementary information.

http://dgv.tcag.ca/dgv/app/downloads?ref=GRCh37/hg19

http://genome.ucsc.edu/index.html

## Acknowledgements

This study makes use of data generated by the DECIPHER community. A full list of centres who contributed to the generation of the data is available from http://decipher.sanger.ac.uk and via email from. Funding for the DECIPHER project was provided by Wellcome.^16^ The authors would like to thank the Clinical Genome Resource (ClinGen) for generating curated content used in this project. We would also like to thank the members of the University of Iowa’s Shivanand R. Patil Cytogenetics & Molecular laboratory, including its past directors, Drs. Val Sheffield and Shivanand R. Patil.

## Notes

### Competing Interest Statement

The authors have declared no competing interest.

### Funding Statement

This work was supported by R01DE021071 (MP and JM), the Stead Family Department of Pediatrics (AW, HM, BD) and the Interdisciplinary Genetics T32 Predoctoral Training Grant (T32 GM 008629; AW).

### Author Declarations

This study used ONLY openly available human data that were obtained from the Database of Genomic Variants (http://dgv.tcag.ca/dgv/app/home) and the Clinical Genome Resource via the UCSC Genome Table Browser (http://genome.ucsc.edu/index.html)

## References

1. Shen, Y. et al. Clinical genetic testing for patients with autism spectrum disorders. Pediatrics 125, e727–35 (2010).

2. Iafrate, A.J. et al. Detection of large-scale variation in the human genome. Nature Genetics 36, 949–951 (2004).

3. Sebat, J. et al. Large-Scale Copy Number Polymorphism in the Human Genome. Science (New York, N.Y.) 305, 525–8 (2004).

4. MacDonald, J.R., Ziman, R., Yuen, R.K., Feuk, L. & Scherer, S.W. The Database of Genomic Variants: a curated collection of structural variation in the human genome. Nucleic Acids Res 42, D986–92 (2014).

5. Manning, M. & Hudgins, L. Array-based technology and recommendations for utilization in medical genetics practice for detection of chromosomal abnormalities. Genet Med 12, 742–5 (2010).

6. Hochstenbach, R. et al. Array analysis and karyotyping: workflow consequences based on a retrospective study of 36,325 patients with idiopathic developmental delay in the Netherlands. Eur J Med Genet 52, 161–9 (2009).

7. Miller, D.T. et al. Consensus statement: chromosomal microarray is a first-tier clinical diagnostic test for individuals with developmental disabilities or congenital anomalies. Am J Hum Genet 86, 749–64 (2010).

8. Rauch, A. et al. Diagnostic yield of various genetic approaches in patients with unexplained developmental delay or mental retardation. Am J Med Genet A 140, 2063–74 (2006).

9. Sagoo, G.S. et al. Array CGH in patients with learning disability (mental retardation) and congenital anomalies: updated systematic review and meta-analysis of 19 studies and 13,926 subjects. Genetics in Medicine 11, 139–146 (2009).

10. Kearney, H.M., Thorland, E.C., Brown, K.K., Quintero-Rivera, F. & South, S.T. American College of Medical Genetics standards and guidelines for interpretation and reporting of postnatal constitutional copy number variants. Genet Med 13, 680–5 (2011).

11. Riggs, E.R. et al. Technical standards for the interpretation and reporting of constitutional copy-number variants: a joint consensus recommendation of the American College of Medical Genetics and Genomics (ACMG) and the Clinical Genome Resource (ClinGen). Genetics in Medicine (2019).

12. Riggs, E.R. et al. Towards an evidence-based process for the clinical interpretation of copy number variation. Clinical Genetics 81, 403–412 (2012).

13. Rehm, H.L. et al. ClinGen — The Clinical Genome Resource. New England Journal of Medicine 372, 2235–2242 (2015).

14. Kent, W.J. et al. The human genome browser at UCSC. Genome Res 12, 996–1006 (2002).

15. Karolchik, D. et al. The UCSC Table Browser data retrieval tool. Nucleic Acids Res 32, D493–6 (2004).

16. Firth, H.V. et al. DECIPHER: Database of Chromosomal Imbalance and Phenotype in Humans Using Ensembl Resources. Am J Hum Genet 84, 524–33 (2009).

17. Online Mendelian Inheritance in Man, OMIM®. (McKusick-Nathans Institute of Genetic Medicine, Johns Hopkins University, Baltimore, MD).

18. Wetzel, A.S. & Darbro, B.W. A Comprehensive List of Human Microdeletion and Microduplication Syndromes. BMC Genomic Data (In Press).

19. Weise, A. et al. Microdeletion and Microduplication Syndromes. Journal of Histochemistry & Cytochemistry 60, 346–358 (2012).

20. Cooper, G.M. et al. A copy number variation morbidity map of developmental delay. Nat Genet 43, 838–46 (2011).

21. Petrovski, S., Wang, Q., Heinzen, E.L., Allen, A.S. & Goldstein, D.B. Genic Intolerance to Functional Variation and the Interpretation of Personal Genomes. PLOS Genetics 9, e1003709 (2013).

22. Zarrei, M., MacDonald, J.R., Merico, D. & Scherer, S.W. A copy number variation map of the human genome. Nat Rev Genet 16, 172–83 (2015).

23. Quinlan, A.R. & Hall, I.M. BEDTools: a flexible suite of utilities for comparing genomic features. Bioinformatics 26, 841–842 (2010).

24. Redon, R. et al. Global variation in copy number in the human genome. Nature 444, 444–54 (2006).

25. Conrad, D.F. et al. Origins and functional impact of copy number variation in the human genome. Nature 464, 704–712 (2010).

26. Kaminsky, E.B. et al. An evidence-based approach to establish the functional and clinical significance of copy number variants in intellectual and developmental disabilities. Genet Med 13, 777–84 (2011).

27. Lappalainen, I. et al. DbVar and DGVa: public archives for genomic structural variation. Nucleic Acids Res 41, D936–41 (2013).

28. Landrum, M.J. et al. ClinVar: improving access to variant interpretations and supporting evidence. Nucleic Acids Res 46, D1062–d1067 (2018).

29. Riggs, E.R. et al. Technical standards for the interpretation and reporting of constitutional copy-number variants: a joint consensus recommendation of the American College of Medical Genetics and Genomics (ACMG) and the Clinical Genome Resource (ClinGen). Genetics in Medicine 22, 245–257 (2020).

30. Riggs, E.R. et al. Correction: Technical standards for the interpretation and reporting of constitutional copy-number variants: a joint consensus recommendation of the American College of Medical Genetics and Genomics (ACMG) and the Clinical Genome Resource (ClinGen). Genet Med 23, 2230 (2021).

31. Wetzel, A.S.M., Heather Parida, M., Manak, J.R. & Darbro, B.W. DISCRIMINATOR: Assigning Cohort-Wide Provisional Pathogenicity Classifications to CNVs. medRxiv (2022).

