## Supplemental Materials for "Benign-Ex: Delineating Regions of the Human Genome Benign to Copy Number Variation"

**Supplementary Methods:**

**Supplemental Figures:**

**Supplemental Tables:**

**1** **Known Pathogenic Region Lists**

**:**

As there is no gold standard for benign regions, the performance of Benign-Ex was assessed by its dissimilarity to a series of lists containing genes and/or regions associated with human disease/disorders. CNV-based lists (Primary CNV List, ClinGen, DECIPHER) were used to account specifically for dosage-sensitive regions, while gene-based lists (Haploinsufficient Genes, OMIM Morbid Map, OMIM All Genes) were used to account for more specific dosage-sensitive genes and gene regions associated with human disorders.

Coordinates for the Primary CNV List correspond to the manually curated list of microduplication and microdeletion syndromes from Wetzel and Darbro (in press) {Wetzel, 2022 #65}. A secondary Primary CNV List excluded intervals that exceeded 10MB in length. Coordinates for the ClinGen Curated Pathogenic CNV List were downloaded from UCSC Table Browser using the iscaCuratedPathogenic track (2020-05-14 release) ^1-3^. Coordinates for the ClinGen Pathogenic CNV List were downloaded from UCSC Table Browser using the iscaPathogenic track (2020-05-14 release) ^1-3^. Coordinates for the DECIPHER CNV List were downloaded from DECIPHER’s CNV Syndrome List ^4^. Coordinates for the Haploinsufficiency list correspond to the curated OMIM Haploinsufficiency gene list (Dataset S1 from Petrovski *et al*. ^5^) supplemented with the intersection of genes with the top 20% of p(HI) scores (DECIPHER Haploinsufficiency Predictions Version 3; [https://‌decipher.‌sanger‌.ac‌.uk‌/](https://decipher.sanger.ac.uk/)) ^4^ and the bottom 25% of RVIS scores (Dataset S2 from Petrovski *et al*. ^5^). Coordinates for the OMIM List were obtained from the OMIM Morbid Map List (morbidmap.txt) and OMIM All Genes List (gene2map.txt) with permission from OMIM ^6^. Filtering was performed to restrict to OMIM entries with a known molecular basis. Included phenotypic entries were annotated with either (3) or (4), and all entries containing the following annotations were excluded: [], {}, ? (see the FAQ section of the OMIM website for annotation descriptions). Unless otherwise specified, UCSC’s LiftOver tool ^7^ was used to convert coordinates between hg18, hg19, and GRCh38.

**2** **Detailed Benign-Ex Methodology**

**:**

Benign-Ex is a python-based program that calls regions of the human genome benign based on user-provided classification or frequency-based data. Benign-Ex is functionally divided into two parts: (1) generation of benign intervals based on user-provided parameters and (2) evaluation of the dissimilarity between the generated benign interval sets and the set of known pathogenic intervals.

Benign-Ex runs from the wrapper file ‘INTERFACE_BENIGNEX.py’, which calls each of the sub-scripts (**Supplemental Figure 10)**. First, Benign-Ex reads the input files (AUTOMATION, DGV_PARAMETERS, CLINGEN_PARAMETERS) to determine which database(s) to use, which portion(s) of Benign-Ex to run, and which set of simulation parameter(s) to use. Benign-Ex is configured to run in one of three ways: only generate the benign intervals, only compare different parameter sets, or generate benign intervals, then compare the parameter sets.

For each parameter set provided by DGV_PARAMETERS, the DGV database file is read, and variants that do not meet the specified year, sample size, methodology, and frequency filters are removed. Similarly, for each parameter set provided by CLINGEN_PARAMETERS, the CLINGEN database file is read, and variants that do not meet the specified classification filter are removed. The coordinates of the remaining variants are compared to identify sets of overlapping variants. For each set, the start and end coordinates of the individual variants are extracted, then sorted to generate a series of intervals across the overlapping region. These regions are further subdivided into 500-bp windows. That window is recorded if the number of variants supporting a 500-bp window exceeds the specified minimum number of events. Once all the windows have been evaluated, all recorded windows are merged if they are within 500-bp of one another to generate the final benign interval. While the coordinates for the final benign intervals are based upon the coordinates of the underlying variants in the database, a 500-bp evaluation window was chosen to remain consistent with our threshold determination analysis (see **Determining the Threshold Values for Benign-Ex Simulations**).

Benign-Ex is currently configured to generate the benign region files for each chromosome individually and then merge the set of chromosome files for each parameter setting to create the genome-wide file. Thus, Benign-Ex will generate ( $N$ ) files based on the number of chromosome * parameter combinations.

$$N=\left( 24 chromosome files \right)*(x parameter settings)$$

Eq. 1

Once all the individual ($N$ ) files have been created, Benign-Ex will merge the individual chromosome files by parameter setting to create the genome-wide file.

For the DGV database only, an optional secondary step (suspicious regions) currently under beta-testing is performed to account for segmental duplications where duplications or deletions may be favored by the calling software used in the original DGV study or ClinGen laboratory or the presence of a CNV within the control genome if the study in question was performed using comparative genomic hybridization. If a region (e.g. GAIN) is supported by at least double the minimum required number of events, all with variants with 1% frequency or greater, and there was at least one overlapping variant in the opposite direction (e.g. LOSS), then the region in the opposite direction (e.g. LOSS) to the original region (e.g. GAIN) is also called benign. A final merging step is then implemented to yield unique benign regions.

For each benign interval list ( $b_{i}$) and known pathogenic interval list ( $p_{i}$ ) combination in the set of known pathogenic intervals ( $P=\left\{ p_{1},p_{2},p_{3} \right\}$; **Supplemental Table 2**) and benign intervals generated by Benign-Ex ( $B=\{b_{1},b_{2},\ldots, b_{n}\}$), the dissimilarity score ($D_{b_{i}, p_{i}}$) is calculated using the overlap coefficient as follows.

$$D_{{(b}_{i}, p_{i)}}= 1-{OC(b}_{i},p_{i})= 1-\left( \frac{b_{i}\cap p{}_{i}}{\min\left( b_{i},p_{i} \right)} \right)$$

*Eq. 2*

The dissimilarity scores are then normalized ( $D_{norm}$ ) across the entire set of generated benign interval lists ( $B=\{b_{1},b_{2},\ldots, b_{n}\}$) for each pathogenic interval list ( $p_{i}$ ).

$$D_{norm}(b_{i}, p_{1})= \frac{D_{{(b}_{i}, p_{1)}}-\min\left( D_{(B, p_{1})} \right)}{\max\left( D_{(B, p_{1})} \right)-\min\left( D_{(B, p_{1})} \right)}$$

*Eq. 3*

Benign-Ex also calculates the sequence content ( $SC$ ) or total base pairs which were called benign for the interval list ( $b_{i}$), and normalizes the sequence content ( $SC$ ) across the set of benign intervals which were generated ( $B=\{b_{1},b_{2},\ldots, b_{n}\}$).

$$SC{}_{norm}\left( b_{i} \right)= \frac{SC\left( b_{i} \right)-\min\left( SC\left( B \right) \right)}{\max\left( SC\left( B \right) \right)-\min\left( SC\left( B \right) \right)}$$

*Eq. 4*

The normalized dissimilarity ( $D_{norm}$ ) and sequence content ( $SC_{norm}$ ) scores for each benign interval list ( $b_{i}$) and known pathogenic interval list ( $p_{i}$ ) combination are then averaged to produce the raw summary statistic ( $BX$ ).

$$BX\left( b_{i},p_{i} \right)=\frac{1}{2}\left( D_{norm}\left( b_{i}, p_{1} \right)+SC{}_{norm}\left( b_{i} \right) \right)$$

*Eq. 5*

To account for the presence of multiple pathogenic region lists and standardize raw values, Benign-Ex applies an Ordered Quantile (ORQ) normalization transformation to the raw values using the “bestNormalize” R package ^8^ to yield the final summary statistic ( ${BX}_{norm}$ ). The optimal Benign-Ex parameter setting for a given known pathogenic region list is that which has the maximal final summary statistic ( $max\left( BX_{norm}\left( B,p_{i} \right) \right) for p_{i} where B=\left\{ b_{1},b_{2},\ldots, b_{n} \right\}$ ). If multiple pathogenic region lists are supplied, then Benign-Ex will average the summary statistic across all the pathogenic lists for each parameter setting.

$$BX{}_{avg}\left( b_{i},P \right)=\frac{1}{n}\sum_{i=1}^{n} BX\_norm\left( b_{i},p_{n} \right)$$

*Eq. 6*

Then, the optimal Benign-Ex parameter setting (across the supplied pathogenic lists and parameter settings) is that which has the maximal average across all pathogenic lists ( $\max\left( BX{}_{avg} \right)$ ). The default configuration of Benign-Ex is to assess the performance of Benign-Ex across all lists; however, users have the option to choose specific lists(s) of interest.

**3** **Determining the Threshold Values for Benign-Ex Simulations**

:

In clinical genetics, variants with a frequency of >1% are often considered too common to be the sole cause of a rare genetic disorder, and variants with a within-study frequency of >10% have a high likelihood of being artifacts or clearly polymorphic. Therefore, we set the upper bound for the minimum allele frequency (AF) at 5%. As the smallest non-zero allele frequency within the DGV dataset was 3.4E-05 or 0.0034%, we set the lower bound for the minimum allele frequency at 0.001% to capture all non-zero allele frequencies. The gain and loss frequencies for variants within DGV are non-normally distributed, with a significant proportion of variants having an AF below 0.01% (77% gain; 65% loss; **Supplemental Figure 11**). During our initial testing, we used a series of allele frequency steps ( [0.01, 0.1, 1, 2] then [0.001, 0.01, 0.1, 1]; data not shown ), however, as Benign-Ex consistently chose the lowest AF threshold possible, this indicated to us that we needed to include more variability around the lower AF thresholds. Therefore, we tested a series of 32 minimum allele frequencies (**Supplemental Table 2**).

A minimum sample size of 30 was chosen to eliminate studies within DGV that only contain data from single individuals or families. The Y/N parameter for study technology is used to restrict DGV entries to those identified by methodologies with higher resolution and/or were developed for genome-wide evaluation of CNVs. The list of acceptable methodologies when the filter is in place include: ‘BAC aCGH’, ‘SNP array’, ‘Oligo aCGH’ and/or ‘Sequencing’. The year ‘2009’ was chosen for the year filter analysis, as papers published in 2008 and 2009 re-defined the breakpoints of previously reported CNVs in DGV and indicated that the actual CNV size was 80-90% smaller than what was identified by the prior technologies ^9,10^. Therefore, the use of the year ‘2009’ provides a second way of filtering for higher-resolution methodologies for CNV detection. Lastly, we varied the minimum number of events from 1 to 10 (**Supplemental Table 2**).

The ClinGen dataset we are using to call benign regions consists of variants that were called benign, likely benign, or a variant of unknown significance. We chose to use all the possible combinations of variants in our simulation set (**Supplemental Table 2**).

First, we performed a preliminary analysis to confirm the functionality of Benign-Ex and ensure it was identifying a reasonable optimal parameter set and not just choosing the most lenient or stringent criteria possible (data not shown). In this analysis, only the minimum number of events was varied, the minimum allele frequency to the lowest possible value (0.001), the sample size to 1, and no methodology or year filters were applied. Benign-Ex *did not* choose the most lenient parameter set as optimal, confirming our summary statistic calculations are performing as intended.

Next, we tested the 2560 unique parameter combinations (filtering criteria and minimum number of qualifying events) for the DGV dataset (1280 gain; 1280 loss) and 120 unique parameter combinations for the ClinGen dataset (60 gain; 60 loss; **Supplemental Table 2**). Benign-Ex chose 2_30_0.004_N_2009 (gain) and 2_30_0.001_N_2009 (loss) as the optimal parameter sets for DGV (**Supplemental Figure 12**) and 1_B (gain) and 1_B_USLB (loss) as the optimal parameters for ClinGen (**Supplemental Figure 13**). As neither the most lenient criteria (1_30_0.001_N and 1_B_USLB_VUS) nor the most stringent criteria (10_30_5_Y_2009 and 10_B) were chosen, this indicates to us that Benign-Ex is performing as intended.

We compared the optimal parameter set and its associated $\max\left( BX{}_{avg} \right)$ value and determined that the suspicious regions processing had little effect (data not shown). Therefore, we chose to skip this step for the remaining analyses. We calculated the total amount of the genome called benign for each of the preliminary parameter settings within the HG19 dataset for both DGV and ClinGen. The range of genome covered was [0.003 – 67.4%] and [0.8 – 61.0%] for DGV gain and loss respectively (**Supplemental Figure 22**), and [0.3 – 74.5%] and [0.1 – 48.4%] for ClinGen gain and loss respectively (**Supplemental Figure 23**). As expected, the percentage of the genome that was called benign is related to the minimum number of events, as well as the level of filtering applied to the databases.

After establishing the functionality of Benign-Ex, we used domain knowledge to establish a smaller set of simulation values for each parameter of the inclusion criteria. As DGV in its entirety covers 81.15% of the human genome, the minimum number of qualifying variants (n=2) for DGV was chosen to limit false positive benign regions by excluding singleton events. The maximum number of qualifying events (n=5) for DGV was chosen based on a two-part analysis of the distribution of overlapping DGV entries.

In the first analysis, we ran Benign-Ex with the most lenient criteria (only the study size threshold applied; EVENTS=1,30,GAIN,0.001,N; EVENTS=1,30,LOSS,0.001,N), allowing us to generate a comprehensive list of all possible benign regions supported by non-zero allele frequencies for our study size threshold. We then determined how many distinct DGV variants were used to support the calling of that benign region (**Supplemental Figure 14**). Approximately half of all the possible benign intervals were supported by a minimum of two DGV entries (59.6% GAIN; 45.6% LOSS). As additional filters (allele frequency, methodology, and year) are applied, the available pool of DGV entries which meet all criteria will decrease. Therefore, we wanted to set the upper bound at a value that maximizes the retention of DGV entries. At n=5, we still have access to 29.9% of the GAIN dataset and 24.3% of the LOSS dataset, and if we exclude the singleton variants, at n=5, we have access to 50.2% of the GAIN dataset and 53.2% of the LOSS dataset.

In the second analysis, we generated both tiled and sliding (data not shown) 500-bp windows across the human genome, then intersected these windows with variants from DGV. DGV entries were first separated by direction (e.g. gain or loss) and then filtered to remove variants from studies with a sample size less than 30 and a minimum of one observed gain/loss (direction specific). Then we determined how many DGV variants overlapped each 500-bp window (**Supplemental Figure 15**). Approximately 35% of the 500-bp windows for both GAIN and LOSS were overlapped by a single DGV entry (35.9% GAIN; 37.3% LOSS; no difference between tiled or sliding), and 85% of the windows were overlapped by five or fewer DGV entries (85.0% GAIN; 84.3% LOSS; no difference between tiled or sliding). Therefore, both of these analyses support setting the upper threshold for the minimum number of qualifying events to five.

Lastly, based on the allele frequency distribution within DGV (**Supplemental Figure 11**) and the performance of the parameter simulations in the preliminary data (**Supplemental Figure 12**), we chose to further restrict the allele frequency thresholds for the final set of Benign-Ex parameters (**Supplemental Table 1**).

We performed a similar analysis for the ClinGen dataset and found that singleton variants comprised approximately half of the database. 60.8% of the gain and 49.5% of the loss benign intervals were supported by two or more variants (**Supplemental Figure 16**), and 66.8% and 54.7% of the sliding 500-bp windows were supported by two or more gain or loss variants respectively (**Supplemental Figure 17**). However, unlike the DGV database, the ClinGen database is much smaller – containing only 3.4% of the variants within DGV – and the variants are curated before being assigned pathogenicity classifications. That, combined with the preliminary result that the optimal calling criteria consist of a single benign or likely benign event, led us to set the minimum number of events at one and the maximum number of events at five. For this final run, we also simplified the pathogenicity classification combinations to those which are the most biologically relevant: B, B/USLB, and B/USLB/VUS (**Supplemental Table 1**). The eliminated combinations (USLB, VUS, USLB/VUS) were for testing purposes to ensure that B and/or B/USLB combinations were being preferred over VUS by Benign-Ex.

**4** **Benign-Ex Usage Requirements & Configuration**

**:**

Benign-Ex requires Python 2.7 or greater (Python 3 is currently not supported), R version 3.6.0 or greater, and a minimum of 420 MB of disk space. The Benign-Ex created files for all 5360 simulations described herein (2560 DGV simulations for hg19 and GRCh38 plus 120 ClinGen simulations for hg19 and GRCh38) are available for download and will require an additional 44 GB of disk space. The Benign-Ex distribution is available for download at [<https://github.com/aswetzel/Benign-Ex>]. Full simulation data is available upon request.

We ran Benign-Ex on both a standalone computer using VMWare® to establish a virtual machine set-up (Intel® Xenon® CPU E3-1545M v5 @ 2.90GHz; 32.0 GB RAM; 4 Cores; 8 Logical Processors) and a high-performance computing cluster (56 compute slots, 2 x Xeon E5-2680v4 (28 Cores at 2.4GHz), 1TB SSD, 1Gbps Ethernet, 100Gbps Omnipath, HPL benchmark performance of 766.1 GFlops). Below are details regarding the processing time on our configuration; the actual runtime of Benign-Ex may vary depending on the hardware used.

Benign-Ex is currently configured to generate each of the ( $N$ ) chromosome * parameter combination files sequentially; therefore, generating the individual chromosome benign interval files is the rate-limiting step for Benign-Ex. For example, the total processing time to generate these files from the most lenient criteria possible from DGV (1_1_GAIN/LOSS_0.001_N) using the virtual machine set-up was 150m 4.732s, while the total processing time to generate these files from a more stringent set of criteria from DGV (1_1_GAIN/LOSS_1_N) was 67m 28.356s. Further, the run time of Benign-Ex will increase with the number of parameter combinations tested. Future iterations of Benign-Ex may allow for parallel processing, but as of now, parallel processing must be done manually by running multiple iterations of Benign-Ex with different parameter settings simultaneously.

The length of time it takes for Benign-Ex to generate each ( $N$ ) file is dependent on the level of filtering applied to the database (i.e. the total amount of data to analyze), but this is not a linear relationship (**Supplemental Figure 18-Supplemental Figure 21**). For the ClinGen database (**Supplemental Figure 18**), the time it takes to generate the individual chromosome benign interval files is related to which chromosome is being processed, with little effect from the minimum number of events. The time taken to generate files is the longest for the ‘ALL’ dataset relative to the ‘B’ or ‘B/USLB’ datasets and ‘GAIN’ versus ‘LOSS’ files. For the DGV database, the effect of filtering is more pronounced, with the processing time decreasing as the minimum number of events and minimum allele frequency increase (**Supplemental Figure 19**). The method and year filters have less of an effect on processing time (**Supplemental Figure 20**); therefore, these differences in the database and file generation length are likely related to the overall sizes of the databases themselves and the number of variant windows to be processed following the application of filters (**Supplemental Figure 21**).

**5** **DGV and ClinGen Database Utilization (hg19)**

**:**

The percentage of the genome that was called benign for the final parameter set (**Supplemental Table 1**) ranged from [2.19-42.33% Gain; 2.05-37.24% Loss] for DGV (**Supplemental Figure 9A**) and [5.71-74.50% Gain; 2.94-48.35% Loss] for ClinGen (**Supplemental Figure 9B**).

Benign-Ex chose 2_30_0.004_N_2009 (gain) and 2_30_0.006_N_2009 (loss) as the optimal parameter sets for the DGV dataset (**Supplemental Figure 1**). A total of 75 studies are present in the 2020 release of DGV; of these, 24 meet the study size and publication year threshold. There was a total of 826,661 total variants in the 2020-02-25 release of DGV. Initial filtering was performed to remove variants with an observation count of zero, classified as inversion events, present on non-canonical chromosomes, or with a length of 0-bp or 1-bp (603,573 Total; 149,463 Gain; 461,359 Loss). Applying the optimal Benign-Ex parameter setting specific filters left 411,965 variants (107,301 Gain; 309,393 Loss). Therefore, with the study-specific and allele frequency thresholds applied, Benign-Ex utilized 68.3% of DGV’s total variants for calling benign regions of the human genome (71.8% Gain; 67.1% Loss).

Benign-Ex chose 1_B (gain) and 1_B_USLB (loss) as the optimal parameters for the ClinGen gain and loss events respectively (**Supplemental Figure 2**). There was a total of 28,133 variants in the 2021-04-01 release of ClinGen, and initial filtering removed variants present on non-canonical chromosomes (27,964 Total; 16,555 Gain; 11,409 Loss). Applying the optimal Benign-Ex parameter setting specific filters left 17,078 variants (9,686 Gain; 7,392 Loss). In total, 61.1% of the total variants (58.5% Gain; 64.8% Loss) within this ClinGen release

These optimal parameter sets called 30% of the genome benign from DGV (29.6% gain; 30.7% loss) and 15% of the genome benign from ClinGen (16.3% gain; 14.7% loss). The percentage of each chromosome called benign for these optimal gain and loss parameter settings can be found in **Supplemental Figure 5**. The size distribution of benign regions is shown in **Supplemental Figure 3**.

**6** **Spatial Correlation Analyses (hg19)**

:

There are three possible scenarios for the correlation between two sets of genomic intervals: (A) intervals are correlated and thus consistently overlap; (B) intervals are anti-correlated and consistently don’t overlap; (C) intervals are independent. Favorov *et al.*’s ^11^ relative distance metric will identify the midpoint of the query interval and the two closest reference intervals. Then, the distance between the query midpoint and closest reference interval midpoint is scaled by dividing by the distance between the two reference interval midpoints. The relative distance metric (bounded by [0,0.5]) can then be calculated for each query interval in the set against the reference interval set. If the two interval sets are independent, we would expect a uniform distribution of relative distance metric values. In contrast, if the interval sets are correlated or anti-correlated, we would expect to see a higher proportion of low or high relative distance metric values respectively. The Bedtools suite ^12^ reldist function applies this metric to calculate spatial correlations between two sets of genomic intervals. Additionally, the Bedtools suite’s ^12^ fisher function computes the Fisher Exact Test as a second method of evaluating spatial relationships while correcting for the genome size and coverage of both interval sets.

To compare the sets of intervals that were generated individually by Benign-Ex from the DGV and ClinGen databases, we first determined the relative contribution of each database to the merged final benign regions (**Supplemental Figure *3*D**). DGV was the larger contributor to these merged regions, with nearly all the merged regions being covered by DGV benign regions (Gain: 98.9%; Loss: 99.9%) and <10% of the merged regions covered by the ClinGen benign regions (Gain: 7.37%; Loss: 1.81%). Next, we compared the benign regions to each other and determined that there was little spatial correlation between the two datasets. The distribution of relative distance metric values obtained from the Bedtools redldist function for the DGV to ClinGen comparison are relatively evenly distributed around 0.02 – the expected value if there is no spatial correlation – for both gains (**Supplemental Figure 4A**) and losses (**Supplemental Figure 4B**).

We next utilized the Bedtools suite ^12^ to compare the copy number variable regions (CNVRs) identified by Zarrei *et al.*’s Stringent and Inclusive CNV Maps to the benign regions identified Benign-Ex for DGV. First, shared genomic regions were identified by a simple overlap. We found that the majority of the CNVRs were overlapped by at least one Benign-Ex benign interval, and a small percentage were overlapped by multiple benign regions (**Supplemental Table 5**). For the Stringent CNV Map, 96.6% and 97.5% of the gain and loss intervals were overlapped by Benign-Ex benign regions respectively, while 2.7% and 1.3% were overlapped by multiple Benign-Ex benign regions. For the Inclusive CNV Map, 85.2% and 92.7% of the gain and loss intervals were overlapped by Benign-Ex benign regions respectively, with 3.7% and 1.6% overlapped by multiple Benign-Ex benign regions.

**7** **Benign Region Overlap with Known Microdeletion and Microduplication Syndromes**

**:**

The intersection of the DGV and ClinGen benign regions contains a total of 436.7 Mb (14.1% genome coverage) in the gain direction and 222.9 Mb (7.2% genome coverage) in the loss direction (**Supplemental Table 6**). Of these shared benign regions, 127.4 Mb (29.2% of the intersection; 4.12% of the genome) of gain intervals and 65.7 Mb (29.5% of the intersection; 2.12% of the genome) also intersect with known microdeletion and microduplication syndromes (MMS) ^13^.

The union of the DGV and ClinGen benign regions contains 1,433.7 Mb (46.3% genome coverage) in the gain direction and 992.0 Mb (32.0% genome coverage) in the loss direction. Of these merged benign regions, 127.4 Mb (29.2% of the intersection; 4.12% of the genome) of gain intervals and 65.7 Mb (29.5% of the intersection; 2.12% of the genome) also intersect with known MMS ^13^.

Zarrei *et al.*’s Stringent and Inclusive CNV Maps contain a total of 64.7 Mb (2.1%) and 111.4 Mb (3.6%) in the gain direction respectively, and 102.4 Mb (3.3%) and 215.2 Mb (7.0%) in the loss direction. Of the CNV Map benign regions, 14.7 Mb (22.7% of the Stringent Gain Map; 0.47% of the genome), 27.0 Mb (24.2% of the Inclusive Gain Map; 0.87% of the genome), 22.7 Mb (22.1% of the Stringent Loss Map; 0.73% of the genome), and 55.5 Mb (25.8% of the Inclusive Loss Map; 1.79% of the genome) intersect with known MMS ^13^.

The intersection of Benign-Ex’s DGV benign intervals, Benign-Ex’s ClinGen benign intervals, and Zarrei et al.’s Stringent CNV Map contains 46.9 Mb (1.52% of the genome) of benign gain regions and 42.8 Mb (1.38% of the genome) of benign loss regions. Interestingly, there is still overlap with the known MMS (10.0 Mb for gain; 9.8 Mb for loss). Removal of these intervals restricts the ultra-conservative benign region interval set to 36.9 Mb (1.19% genome) for gains and 33.0 Mb (1.07% genome) for losses. Segmental duplications overlap the majority of the ultra-conservative benign region intervals: 27.9 Mb (75.6%) and 20.5 Mb (62.1%) for gain and loss regions respectively.

**8** **Benign-Ex Results for GRCh38**

**:**

The same analysis as described above for HG19 (**Supplemental Figure 11**; **Supplemental Figure 14-Supplemental Figure 15**) was performed for the DGV HG38 dataset to establish the upper and lower bounds for the minimum number of events and allele frequency (**Supplemental Figure 24**). As with HG19, we looked at the distribution of allele frequencies across the DGV dataset to set the minimum (0.001%) and maximum allele frequency (0.09%) thresholds. The sliding window analysis for GRCh38 showed a similar pattern to that of HG19; therefore, we set the upper threshold for the minimum number of events to five (**Supplemental Figure 25**). Benign-Ex chose 2_30_0.007_N_2009 (gain) and 2_30_0.006_N_2009 (loss) as the optimal parameter sets for the DGV dataset (**Supplemental Figure 26**), and 1_B_USLB as the optimal parameters for both ClinGen gain and loss events (**Supplemental Figure 27**).

**9** **Sex and Ethnicity Data for DGV**

**:**

Publicly available sample level data for studies included in the 2020-02-25 release of DGV were downloaded from DGV ^14^ and dbVar ^15^. Two studies (nstd166 ^16^ and nstd41 ^17^) had no publicly available sample data. Reported sex and ethnicity data were compiled from the two sources for all remaining studies, and unique samples were identified. In total, sample level data were available for 44,129 unique samples across 73 studies. Reported sex was available for 93% of unique samples (n=40,951; 92.8%). Ethnicity data was only provided for 27% of samples (n=14,899); however, 114 different ethnicities from 54 different countries were represented (**Supplemental Figure 28-29**). The majority of duplicate samples (n=15,746 of 19,170; 82.13%) were common to both nstd54 ^18^ and nstd100 ^19^ and represented patients who underwent clinical testing for intellectual disability, developmental delay, or related phenotypes at Signature Genomic Libraries, LLC. Most remaining duplicate samples (n=2,707 of 3,424; 79.06%) and all samples found in more than five studies (n=766 of 766) were from large-scale human genetic variation projects: International HapMap Project ^20,21^, 1000 Genomes Project ^22^, and the Human Genome Diversity Project ^23^.

**Supplemental Table 1: Final Benign-Ex Simulation Parameters**

**.** Top: Database of Genomic Variants. Bottom: ClinGen. Tables display the list of simulation values tested for each parameter. ‘B’ denotes benign classifications, ‘B/USLB’ denotes benign and uncertain significance likely benign classifications, and ‘ALL’ denotes benign, uncertain significance likely benign, and variant of unknown significance classifications.

| **Database of Genomic Variants** | **Values** |
| --- | --- |
| Minimum Number of Qualifying Variants | 2, 3, 4, 5 |
| Minimum Sample Size of the Study | 30 |
| Direction of CNV Event | GAIN, LOSS |
| Minimum Frequency of CNV Event within the Study (%) | 0.001 to 0.009 by 0.001 ∷ ∷ 0.01 to 0.09 by 0.01 |
| Exclude CNVs NOT identified by specific methodologies? | Y, N |
| Exclude CNVs NOT identified by Studies published after X Year? | {NA}, 2009 |

| **ClinGen** | **Values** |
| --- | --- |
| Minimum Number of Qualifying Variants | 1, 2, 3, 4, 5 |
| Acceptable CNV Pathogenicity Classification(s) | B, B/USLB, B/USLB/VUS |

**Supplemental Table 2: Preliminary Benign-Ex Simulation Parameters**

**.** Top: Database of Genomic Variants. Bottom: ClinGen. Tables display the list of simulation values tested for each parameter. ‘B’ denotes benign classifications, ‘B/USLB’ denotes benign and uncertain significance likely benign classifications, and ‘ALL’ denotes benign, uncertain significance likely benign, and variant of unknown significance classifications.

| **Database of Genomic Variants** | **Values** |
| --- | --- |
| Minimum Number of Qualifying Variants | 1, 2, 3, 4, 5, 6, 7, 8, 9, 10 |
| Minimum Sample Size of the Study | 30 |
| Direction of CNV Event | GAIN, LOSS |
| Minimum Frequency of CNV Event within the Study (%) | 0.001 to 0.009 by 0.001 ∷ ∷ 0.01 to 0.09 by 0.01  0.1 to 0.9 by 0.1 ∷ ∷ 1 to 5 by 1 |
| Exclude CNVs NOT identified by specific methodologies? | Y, N |
| Exclude CNVs NOT identified by Studies published after X Year? | {NA}, 2009 |

| **ClinGen** | **Values** |
| --- | --- |
| Minimum Number of Qualifying Variants | 1, 2, 3, 4, 5, 6, 7, 8, 9, 10 |
| Acceptable CNV Pathogenicity Classification(s) | B, USLB, VUS, B/USLB, USLB/VUS, B/USLB/VUS |

**Supplemental Table 3: Pairwise Comparisons Between Stringent & Inclusive CNV Map and Benign-Ex.**

|  |  | **INTERSECTION** | **UNION** | **MINIMUM** | **JACCARD** | **OVERLAP** | **AVERAGE** | **FISHER** |
| --- | --- | --- | --- | --- | --- | --- | --- | --- |
| **Stringent vs Inclusive**  **CNV Map** | GAIN | 64,727,102 | 107,551,591 | 64,727,102 | 0.602 | 1.000 | 0.801 | p < $e^{-3,045} \approx$ 0 |
|  | LOSS | 102,414,579 | 215,164,504 | 102,414,579 | 0.476 | 1.000 | 0.738 | p < $e^{-26,007} \approx$ 0 |
| **Benign-Ex DGV vs**  **Inclusive CNV Map** | GAIN | 97,804,075 | 930,109,917 | 111,439,374 | 0.105 | 0.878 | 0.491 | p < ${e,}^{-1748} \approx$ 0 |
|  | LOSS | 174,361,697 | 799,014,183 | 215,164,504 | 0.218 | 0.810 | 0.514 | p < $e^{-\infty} \approx$ 0 |
| **Benign-Ex DGV vs**  **Stringent CNV Map** | GAIN | 62,356,932 | 907,230,693 | 64,727,102 | 0.069 | 0.963 | 0.516 | p < $e^{-\infty} \approx$ 0 |
|  | LOSS | 96,352,527 | 764,273,428 | 102,414,579 | 0.126 | 0.941 | 0.533 | p < $e^{-10,162} \approx$ 0 |

**Supplemental Table 4: Stringent and Inclusive CNV Map Summary Data**

**.**

|  |  | **# Benign Regions** | **Median Benign Region Length (kb)** | **Genome Coverage (Mb; %)** |
| --- | --- | --- | --- | --- |
| **Stringent CNV Map** | GAIN | 1,169 | 9.74±3.95 | 64.7 Mb (2.1%) |
|  | LOSS | 11,530 | 1.14±0.50 | 102.4 Mb (3.3%) |
| **Inclusive CNV Map** | GAIN | 3,132 | 3.33±2.23 | 111.4 Mb (3.6%) |
|  | LOSS | 23,438 | 0.96±0.47 | 215.2 Mb (7.0%) |

**Supplemental Table 5: Overlap Between Benign-Ex Benign Regions and Stringent and Inclusive CNV Map CNVRs**

**.**

|  |  | **# Overlaps with Benign-Ex DGV** | **Median Overlap Length (kb)** | **Median Size Difference (kb)** |
| --- | --- | --- | --- | --- |
| **Stringent CNV Map** | GAIN | 1,195 (96.6%) | 9.49±3.70 | 81.9±41.7 |
|  | LOSS | 11,473 (97.5%) | 0.97±0.47 | 0.05±4.33 |
| **Inclusive CNV Map** | GAIN | 3,063 (85.2%) | 3.35±1.85 | 23.6±22.8 |
|  | LOSS | 23,475 (92.7%) | 0.86±0.32 | 0.00±3.86 |

**Supplemental Table 6: Benign Region Overlap with Known Microdeletion and Microduplication Syndromes**

**.**

|  |  |  |  | **Benign Regions** $\boldsymbol{\cap}$ **MMS** | | |
| --- | --- | --- | --- | --- | --- | --- |
|  |  | **Total Length (bp)** | **% Genome Coverage** | **Total Length (bp)** | **% MMS Coverage** | **% Genome Coverage** |
| **Benign-Ex**  **DGV** $\boldsymbol{\cap}$ **ClinGen** | GAIN | 436,658,619 | 14.1% | 127,392,649 | 29.2% | 4.12% |
|  | LOSS | 222,862,089 | 7.2% | 65,670,810 | 29.5% | 2.12% |
| **Benign-Ex**  **DGV** $\boldsymbol{\cup}$ **ClinGen** | GAIN | 1,433,831,117 | 46.3% | 379,020,736 | 26.4% | 12.24% |
|  | LOSS | 992,030,575 | 32.0% | 257,495,579 | 26.0% | 8.32% |
| **Stringent CNV Map** | GAIN | 64,727,102 | 2.1% | 14,704,242 | 22.7% | 0.47% |
|  | LOSS | 102,414,579 | 3.3% | 22,673,100 | 22.1% | 0.73% |
| **Inclusive CNV Map** | GAIN | 111,439,374 | 3.6% | 27,013,803 | 24.2% | 0.87% |
|  | LOSS | 215,164,504 | 7.0% | 55,519,791 | 25.8% | 1.79% |

**Supplemental Figure 1: Final HG19 Benign-Ex Heatmaps for DGV**

**.** To visually demonstrate how different Benign-Ex parameter sets perform across multiple pathogenic lists, Benign-Ex produces the following heatmap. The histogram on the top represents the average performance of that parameter set across all pathogenic lists. The maximum value is the optimal parameter setting across all pathogenic lists. The histogram on the right represents the average performance of all Benign-Ex parameter sets across that pathogenic list. The highest performing parameter set is indicated for each pathogenic list. The heatmap in the middle plots the final summary statistic for each pathogenic list and Benign-Ex parameter set combination. The Benign-Ex parameter simulations are sorted first by the minimum number of events, then left to right by the combination of the year and methodology filter applications: (N/2009, N/NA, Y/2009, Y/NA). Finally, the simulations are sorted by allele frequency (AF) from smallest to largest. Red indicates negative scores, Blue indicates positive scores, and White indicates a ‘0’ score. (A) Final 288 DGV Gain Simulations, (B) DGV Gain Simulations for Method=N; Year=NA (C) DGV Gain Simulations for Method=N; Year=2009, (D) DGV Gain Simulations for Method=Y; Year=NA; (E) DGV Gain Simulations for Method=Y; Year=2009; (F) Final 288 DGV Loss Simulations, (G) DGV Loss Simulations for Method=N; Year=NA (H) DGV Loss Simulations for Method=N; Year=2009, (I) DGV Loss Simulations for Method=Y; Year=NA; (J) DGV Loss Simulations for Method=Y; Year=2009.

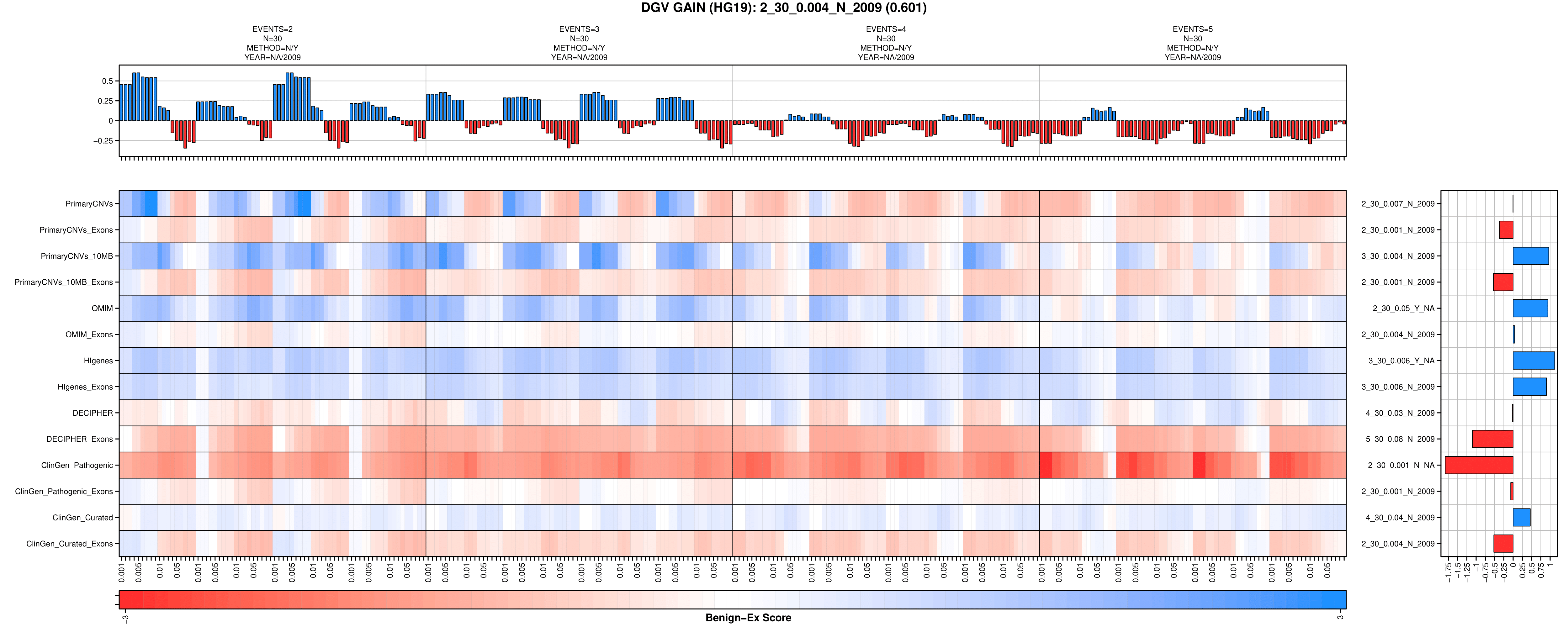

A

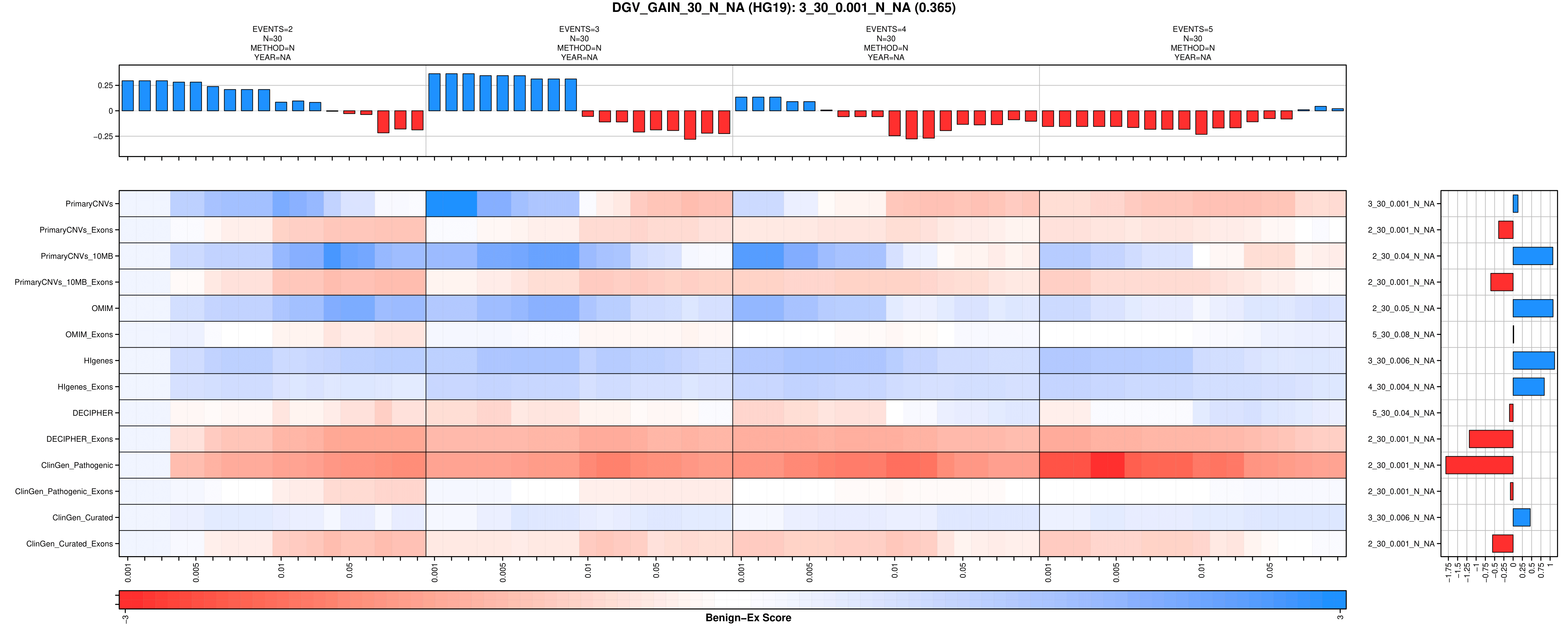

B

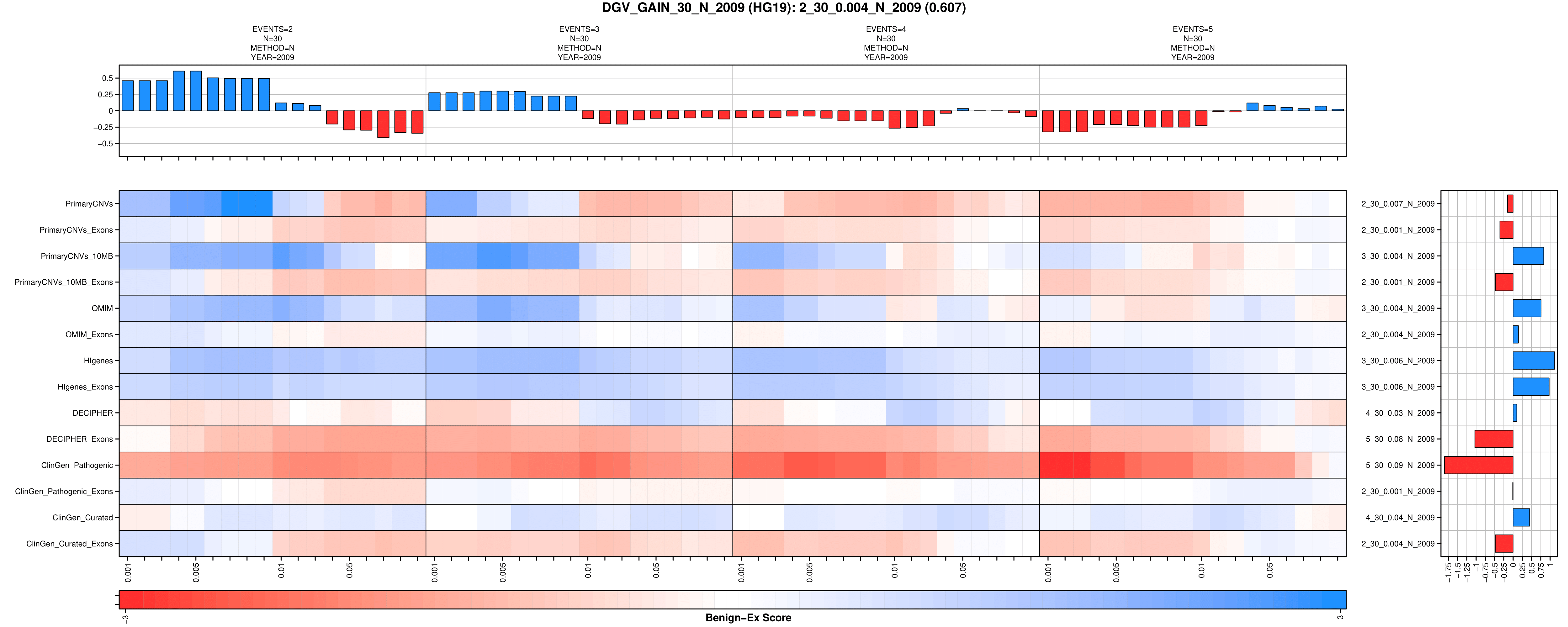

C

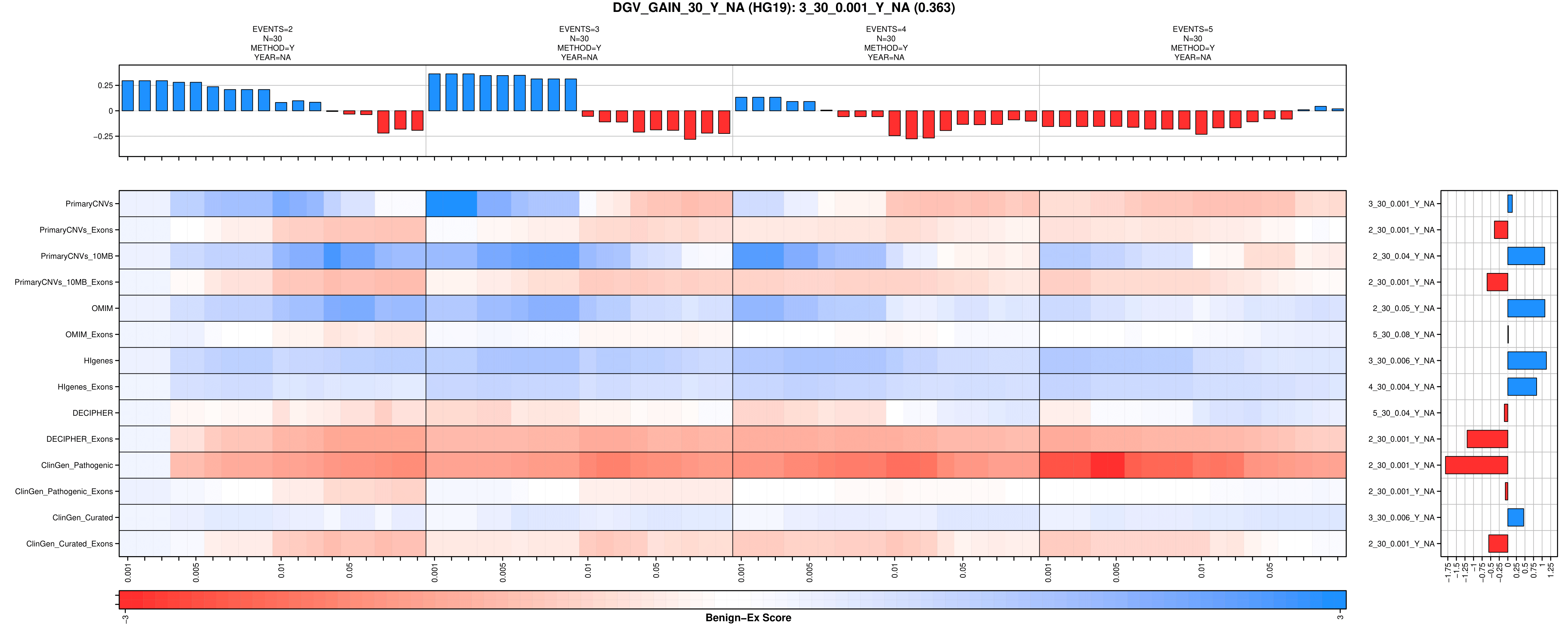

D

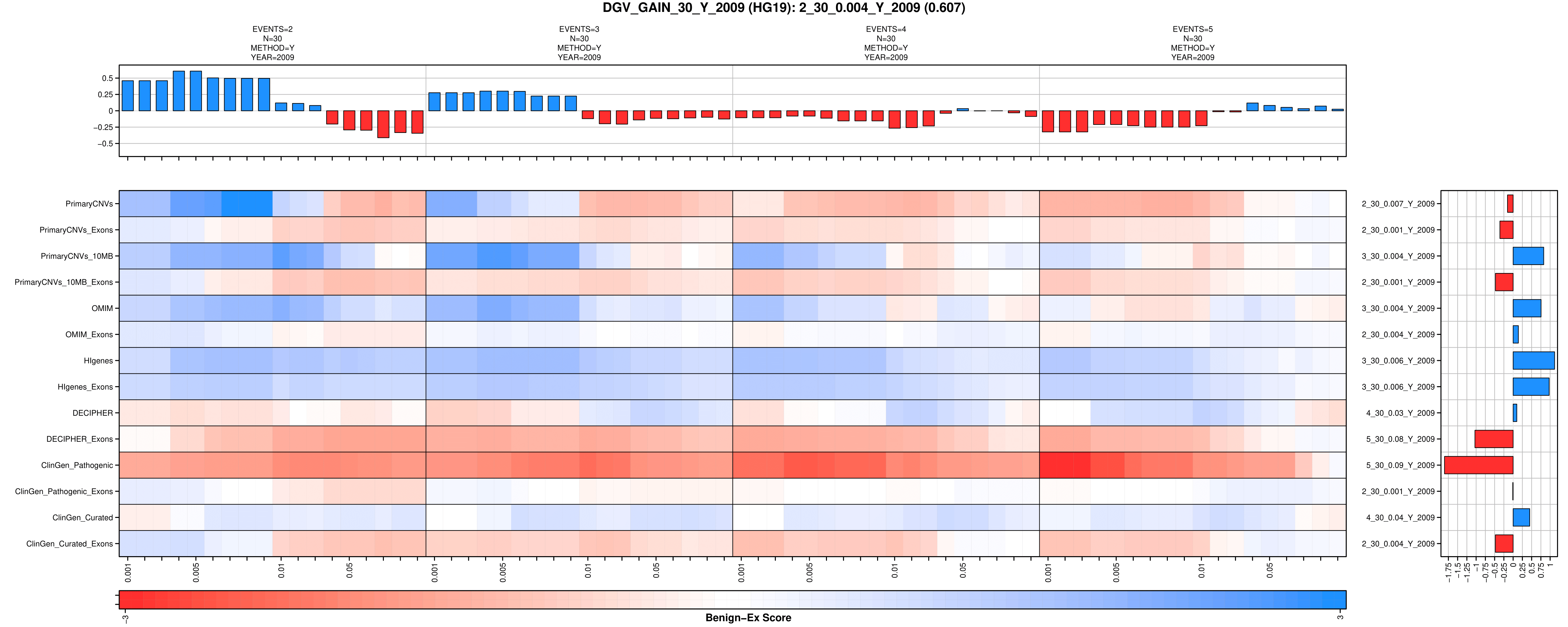

E

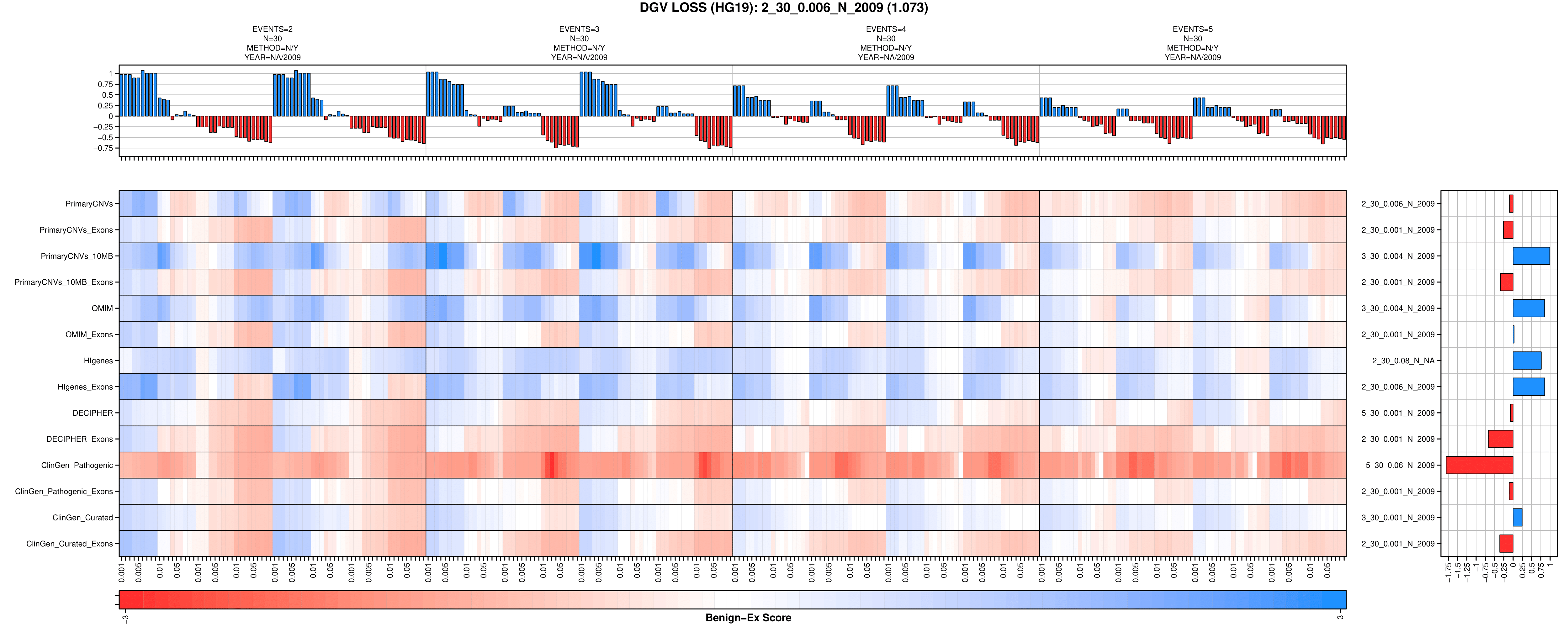

F

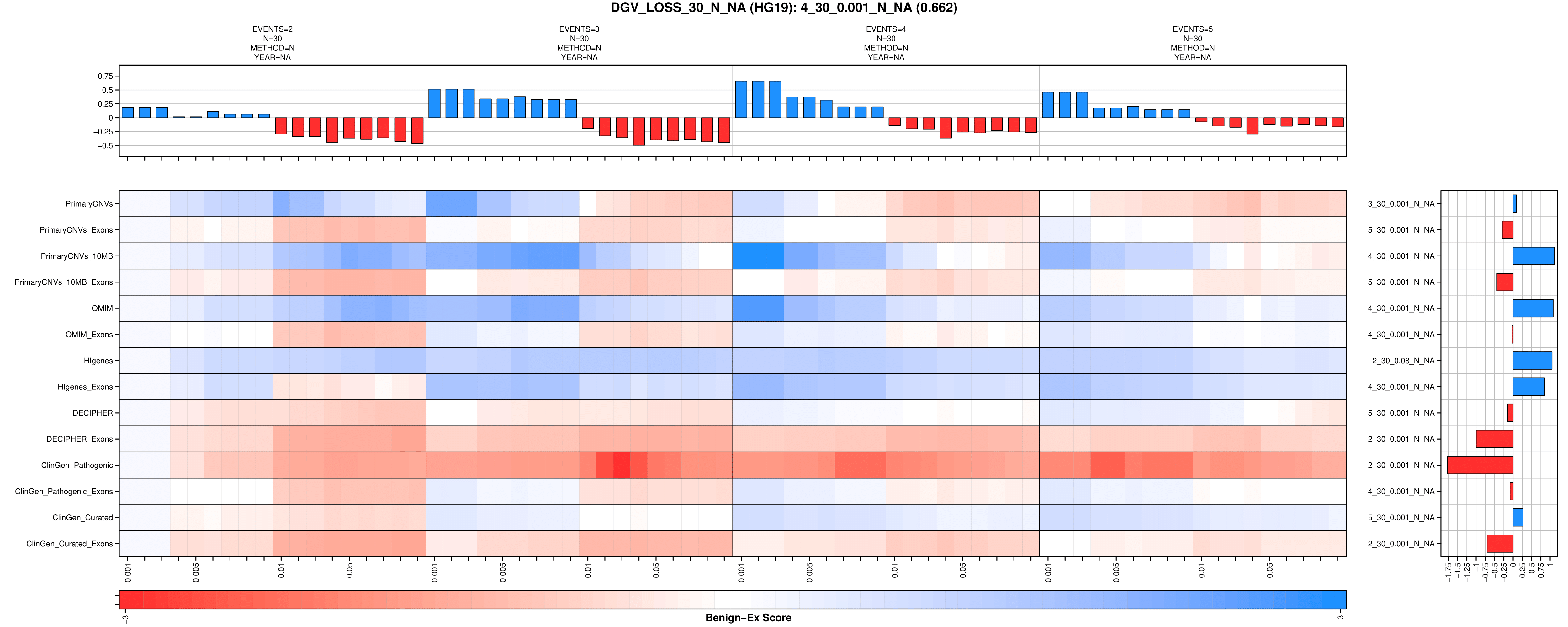

G

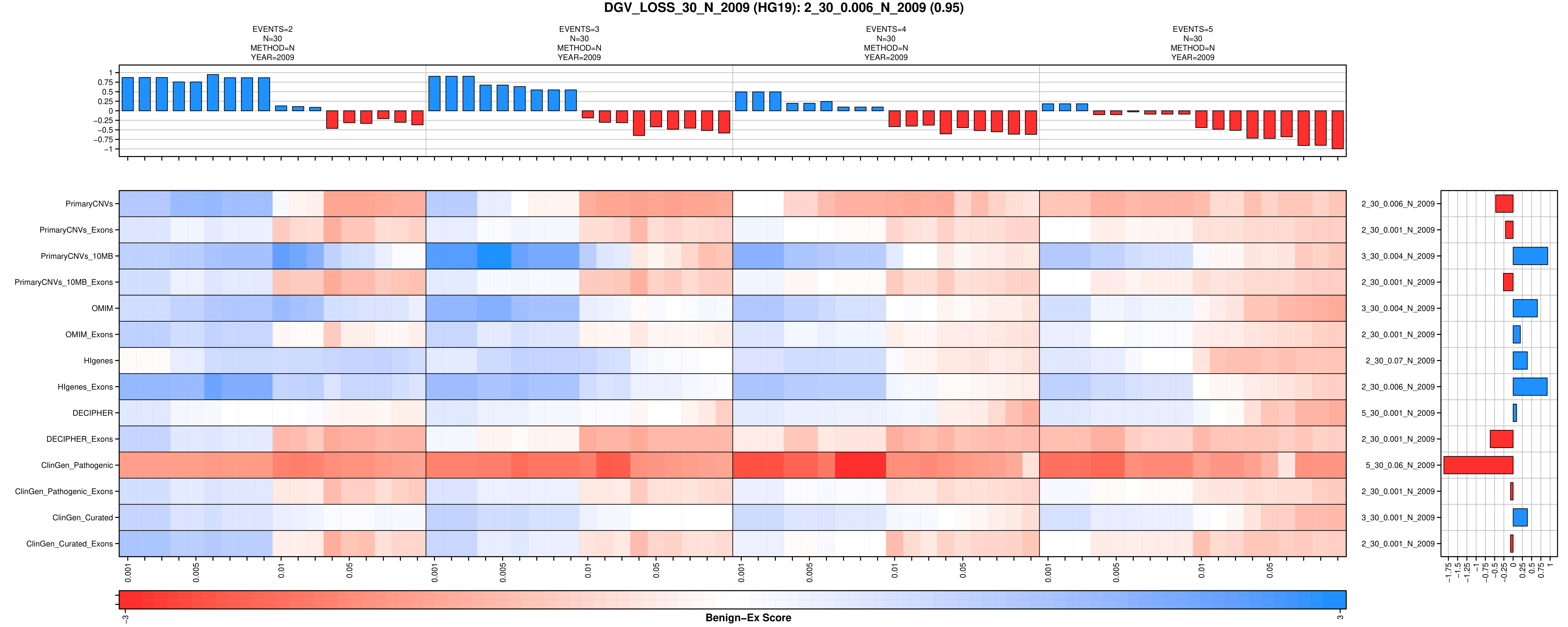

H

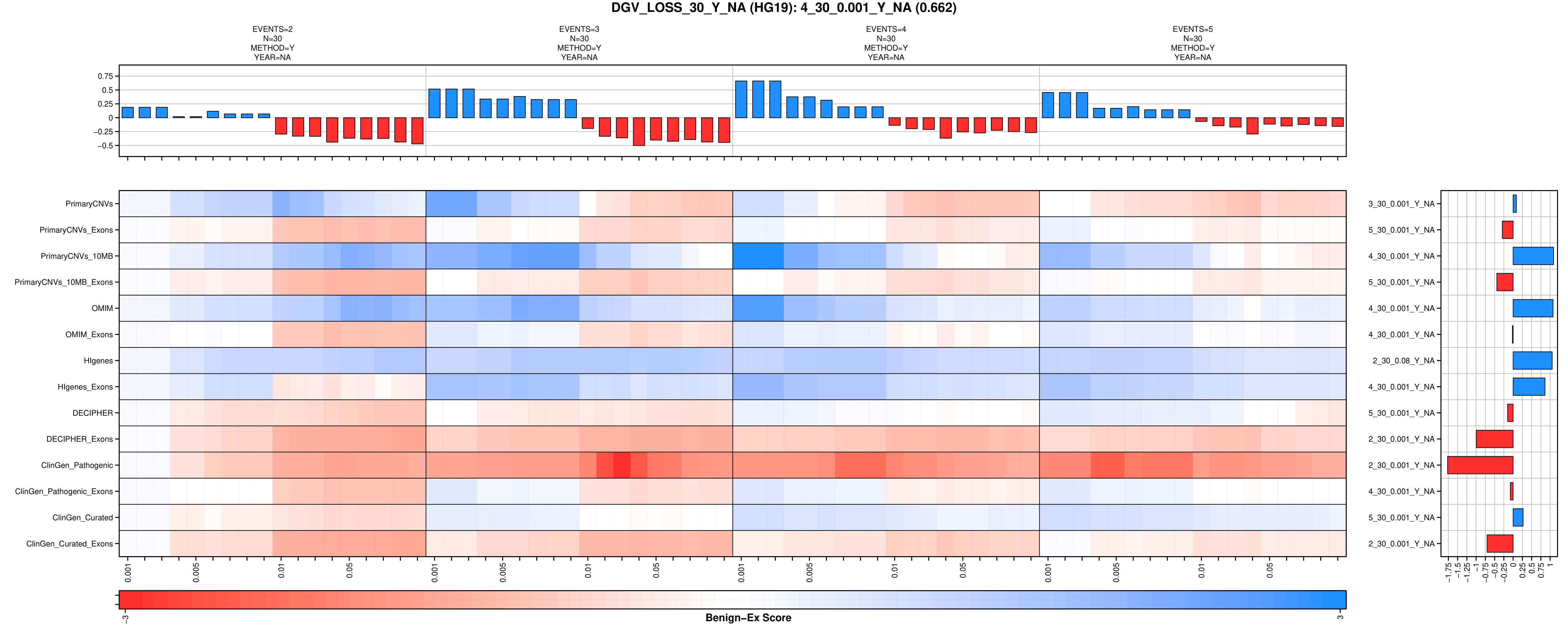

I

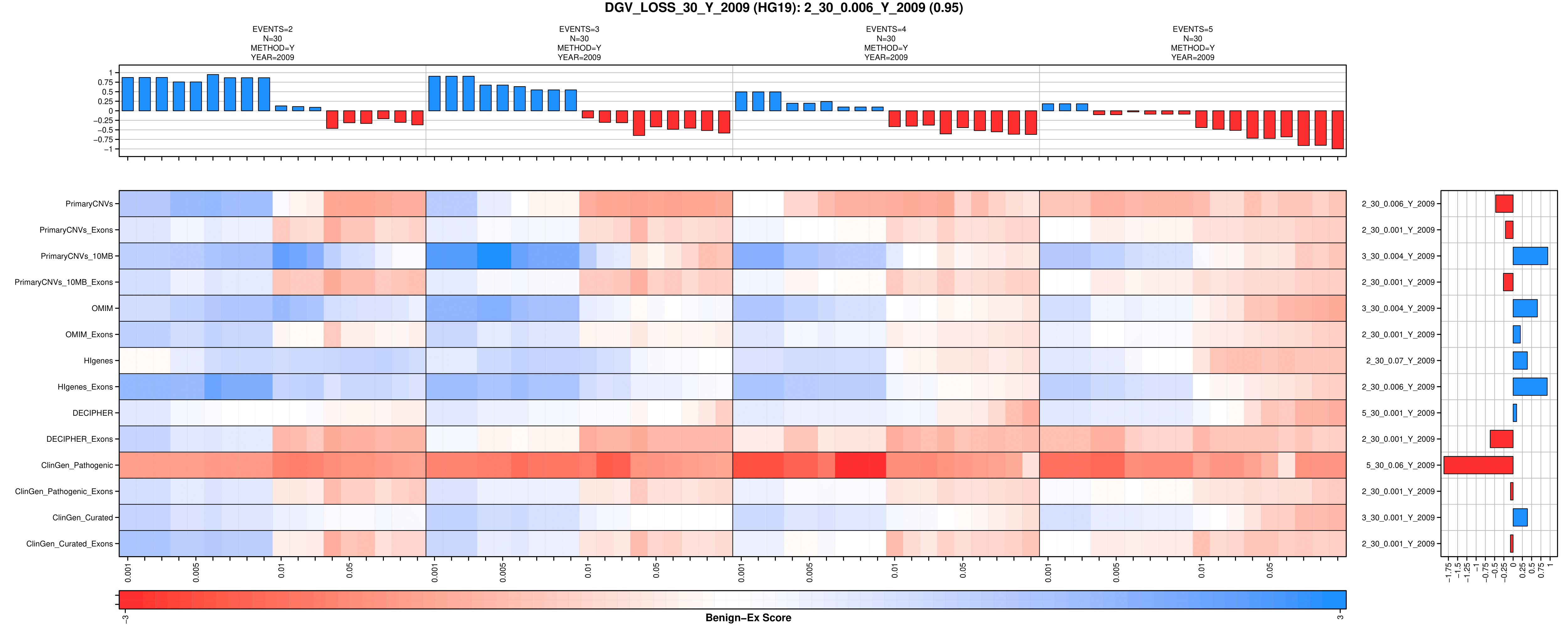

J

**Supplemental Figure 2: Final HG19 Benign-Ex Heatmaps for ClinGen**

**.** To visually demonstrate how different Benign-Ex parameter sets perform across multiple pathogenic lists, Benign-Ex produces the following heatmap. The histogram on the top represents the average performance of that parameter set across all pathogenic lists. The maximum value is the optimal parameter setting across all pathogenic lists. The histogram on the right represents the average performance of all Benign-Ex parameter sets across that pathogenic list. The highest performing parameter set is indicated for each pathogenic list. The heatmap in the middle plots the final summary statistic for each pathogenic list + Benign-Ex parameter set combination. The Benign-Ex parameter simulations are sorted by the acceptable classifications (‘B’, ‘B/USLB’, ‘B/USLB/VUS’). Red indicates negative scores, Blue indicates positive scores, and White indicates a ‘0’ score. (A) Final 15 ClinGen Gain Simulations, (B) All 15 ClinGen Loss Gain Simulations.

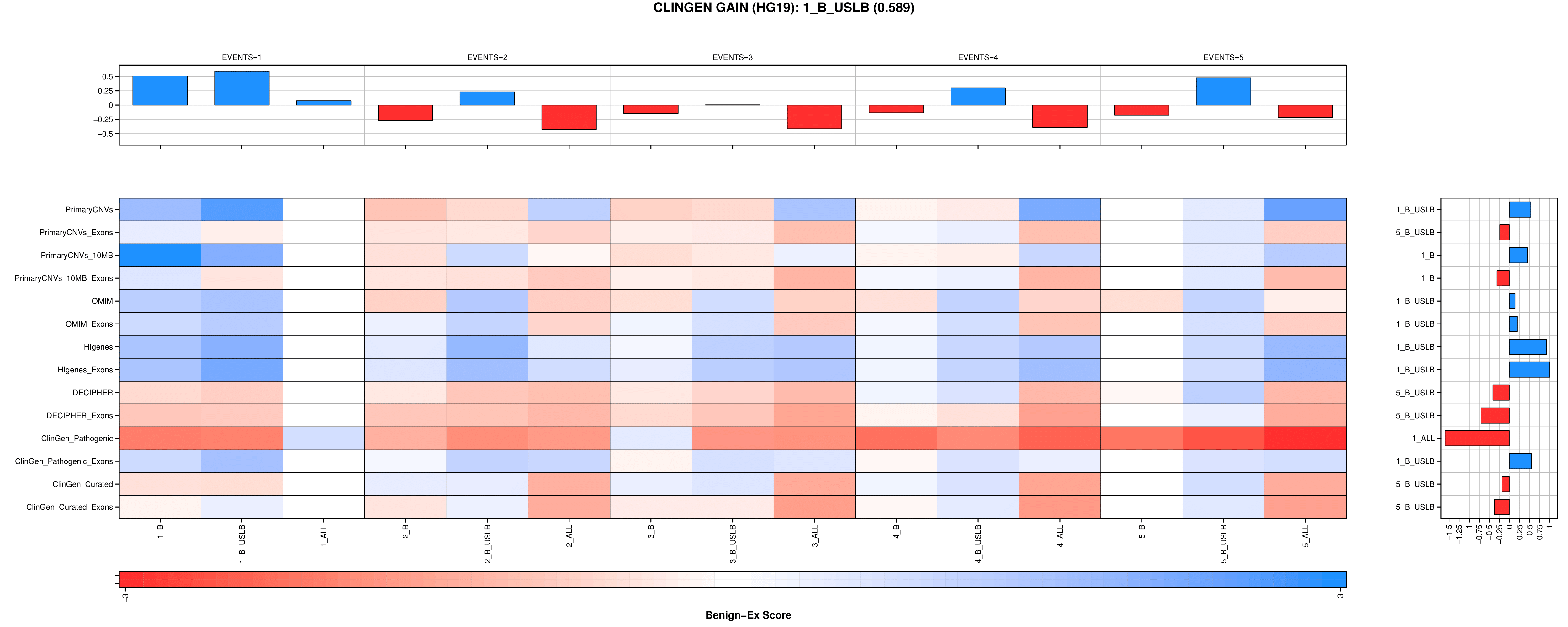

A

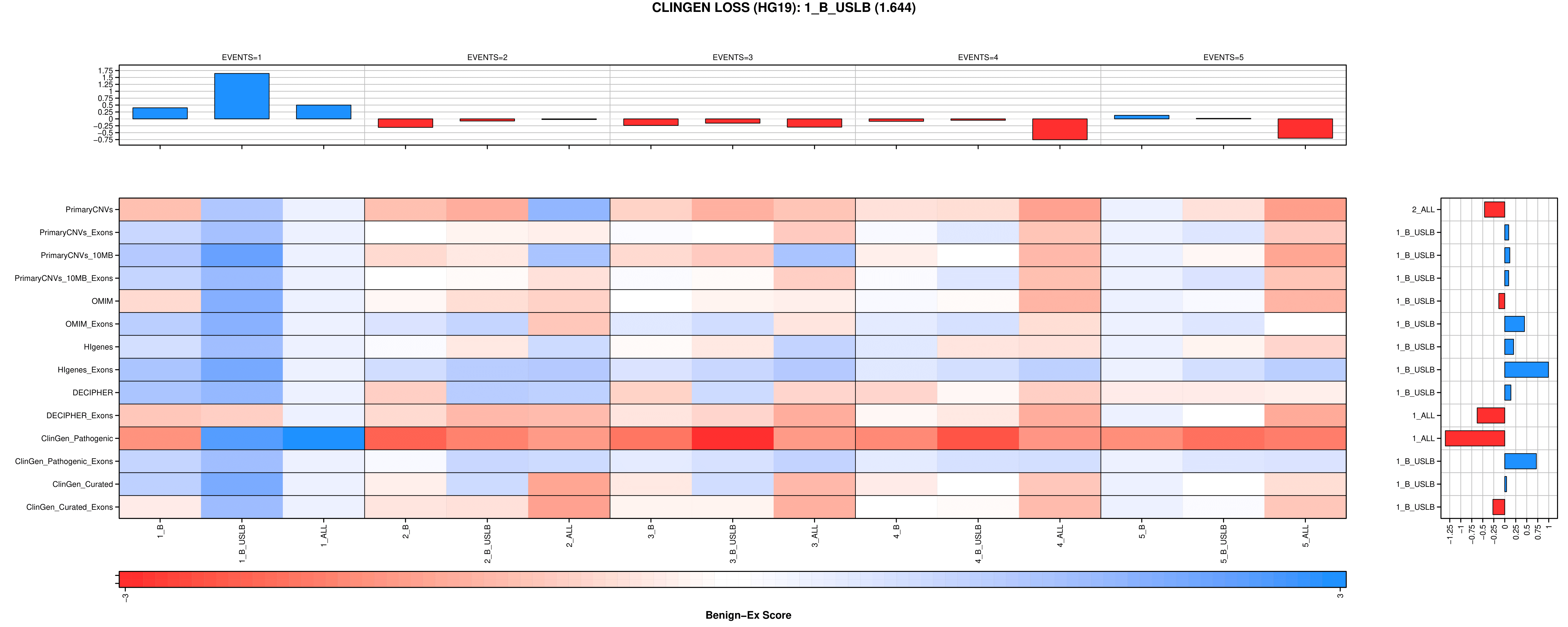

B

**Supplemental Figure 3: Size Distribution of Benign Regions**

**.** The size distribution of benign regions called by Benign-Ex for gains (blue), losses (red), and merged gains and losses (grey) are shown for (A) DGV, (B) ClinGen, and (C) merged DGV+ClinGen. Vertical black lines represent the median benign region length. (D) The relative contribution of benign regions from the DGV (blue, red, grey) and ClinGen (black) databases to the final merged benign regions.

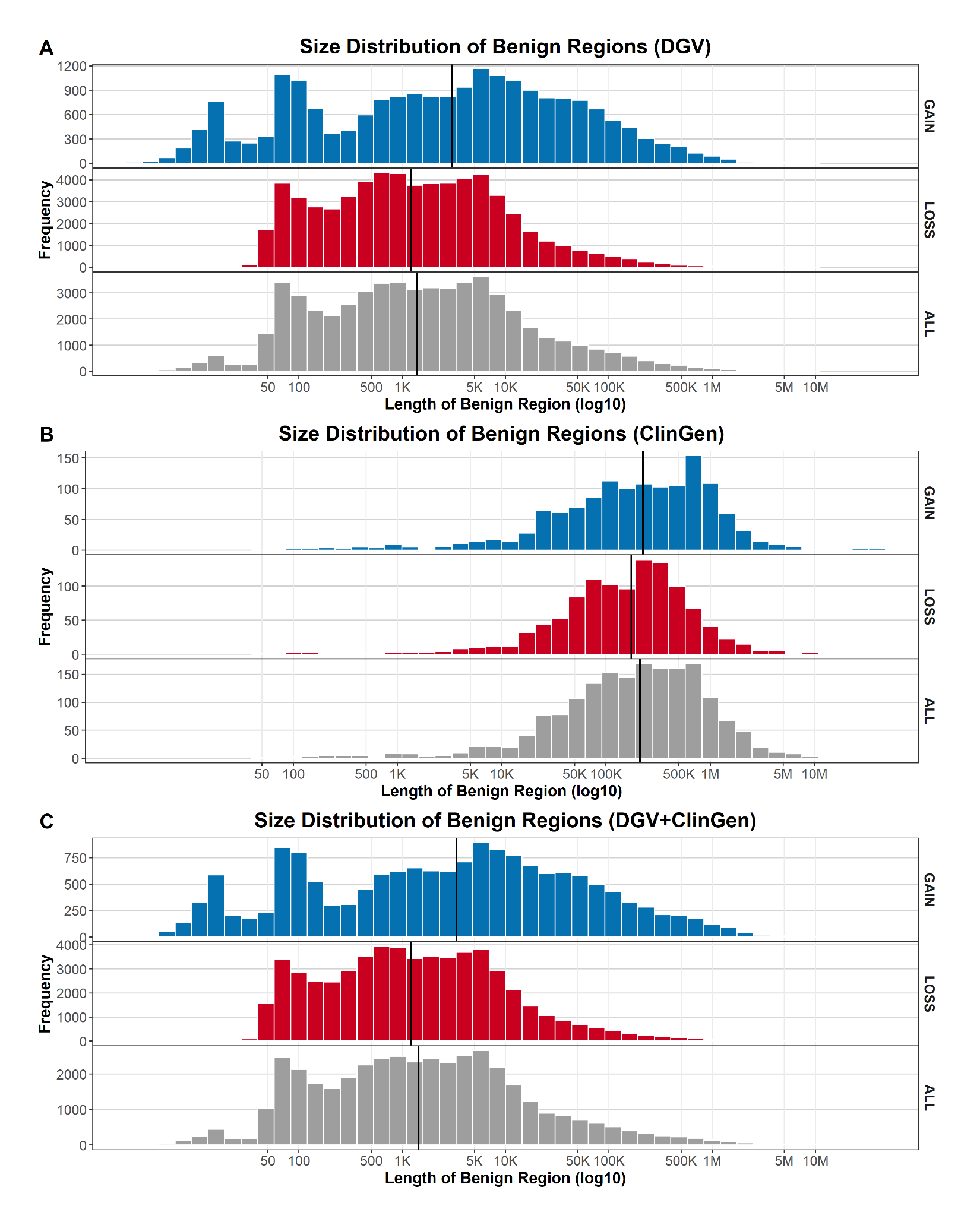

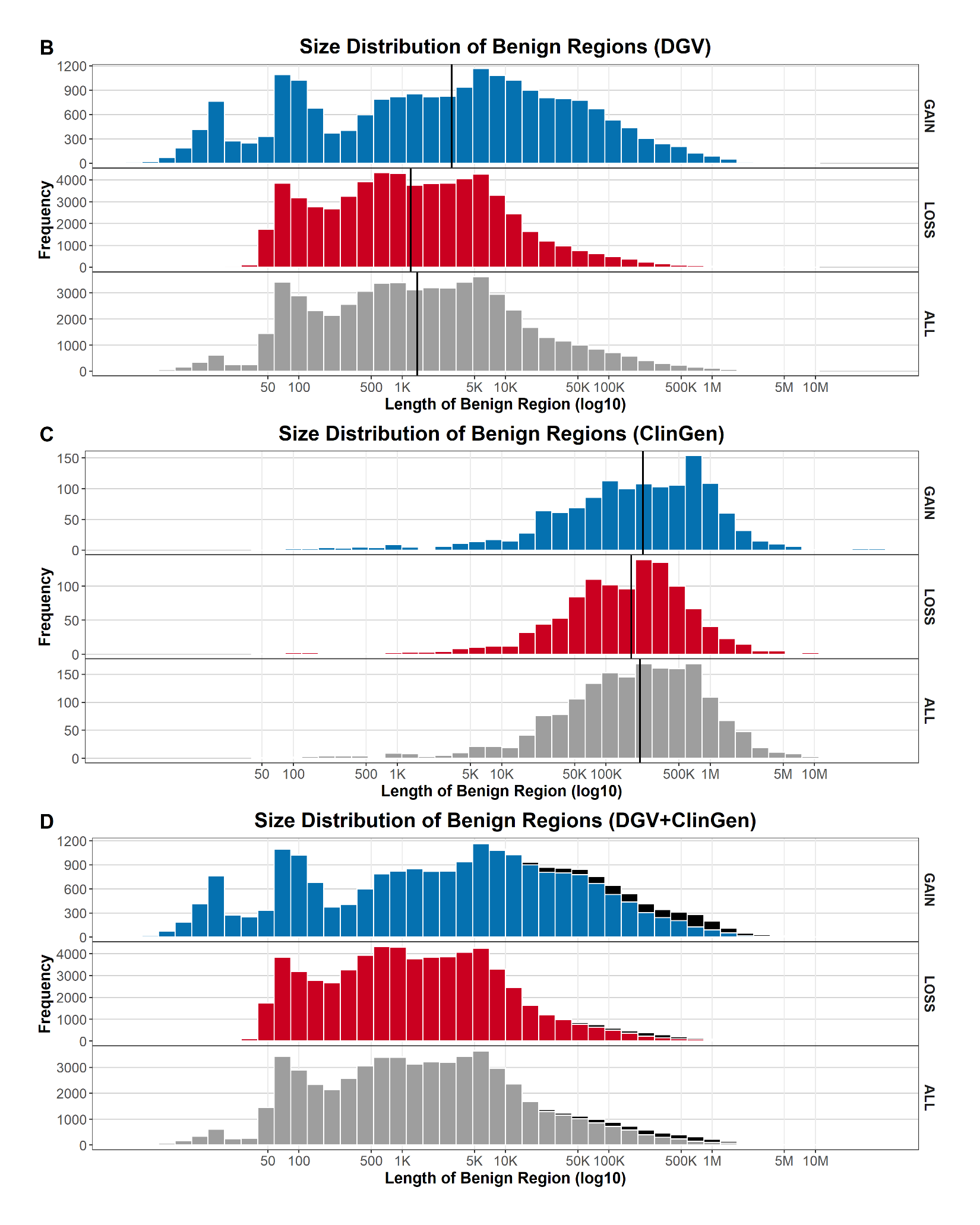

**Supplemental Figure 4: Spatial Correlation of Benign Regions: Benign-Ex’s DGV vs ClinGen**

**.** Spline graph depicting the spatial correlation between Benign-Ex’s DGV and ClinGen benign regions as assessed by the Bedtools reldist function for (A) gains and (B) losses. For each graph, both reciprocal comparisons are plotted.

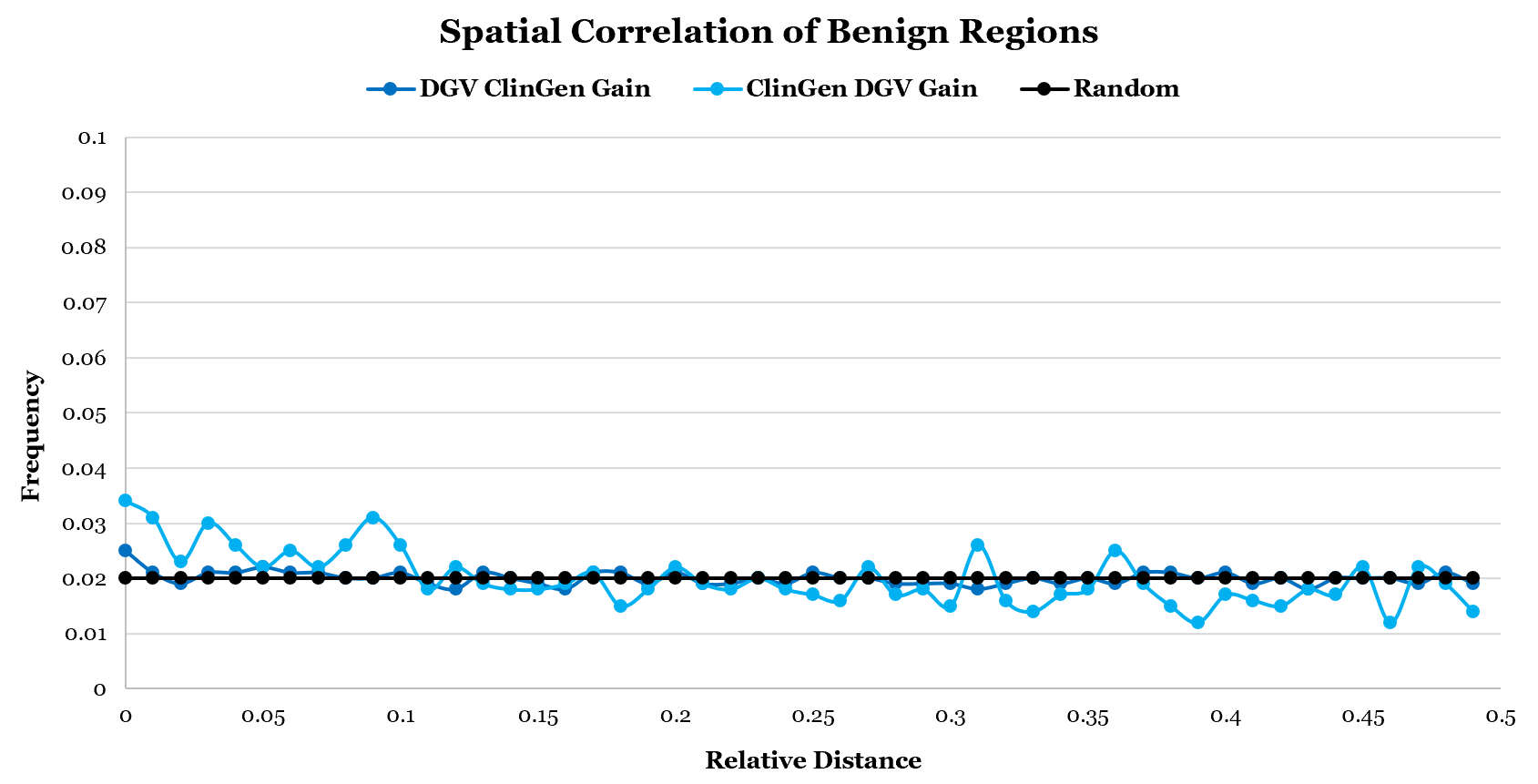

A

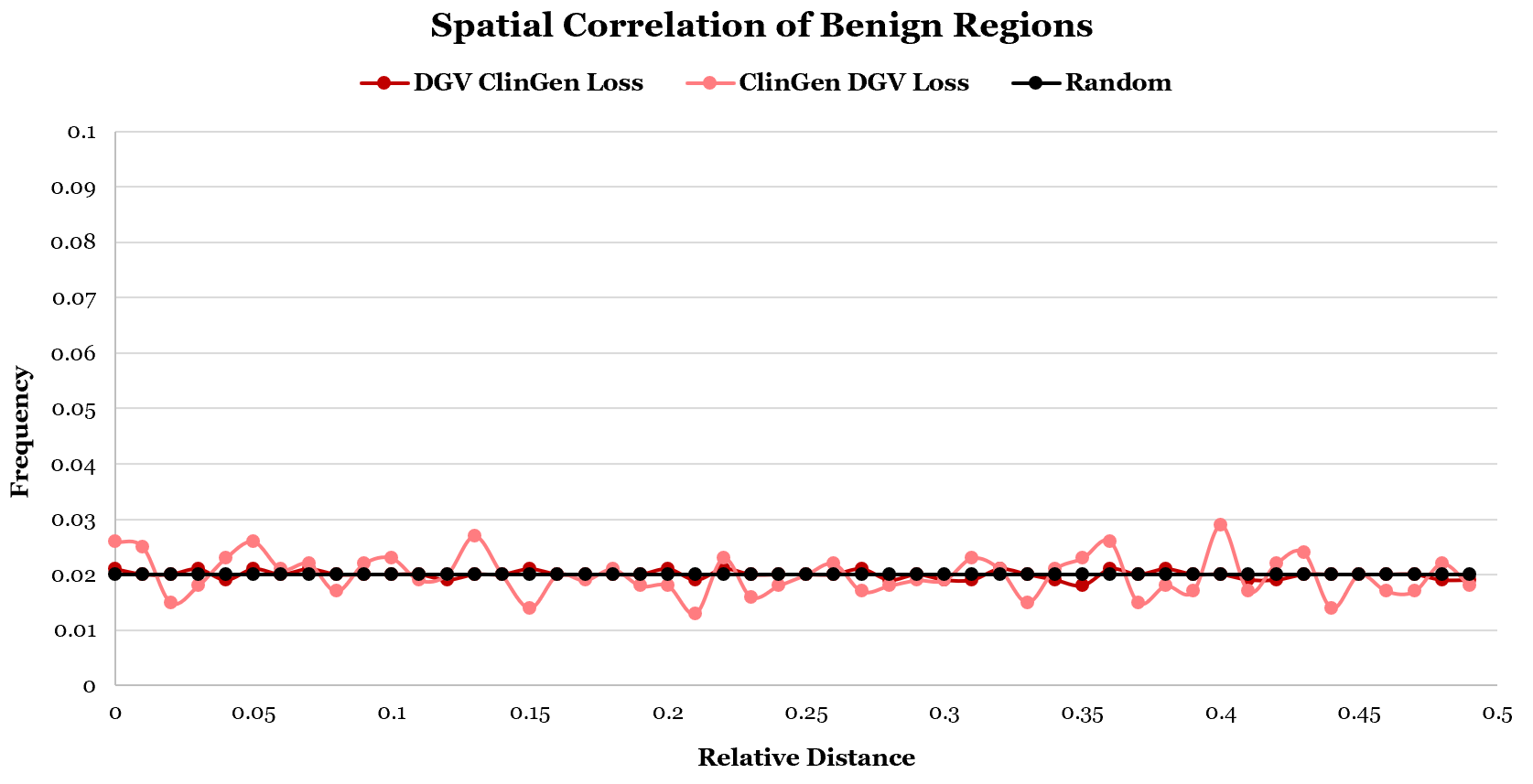

B

**Supplemental Figure 5: Percent Coverage by Chromosome for the Optimal Benign-Ex Parameter Settings**

**.** The percentage of each chromosome called benign for the optimal gain and loss parameter setting by Benign-Ex was tracked for the HG19 dataset. The genome coverage average for gain and loss is indicated by the black dashed lines. (A) DGV (B) ClinGen.

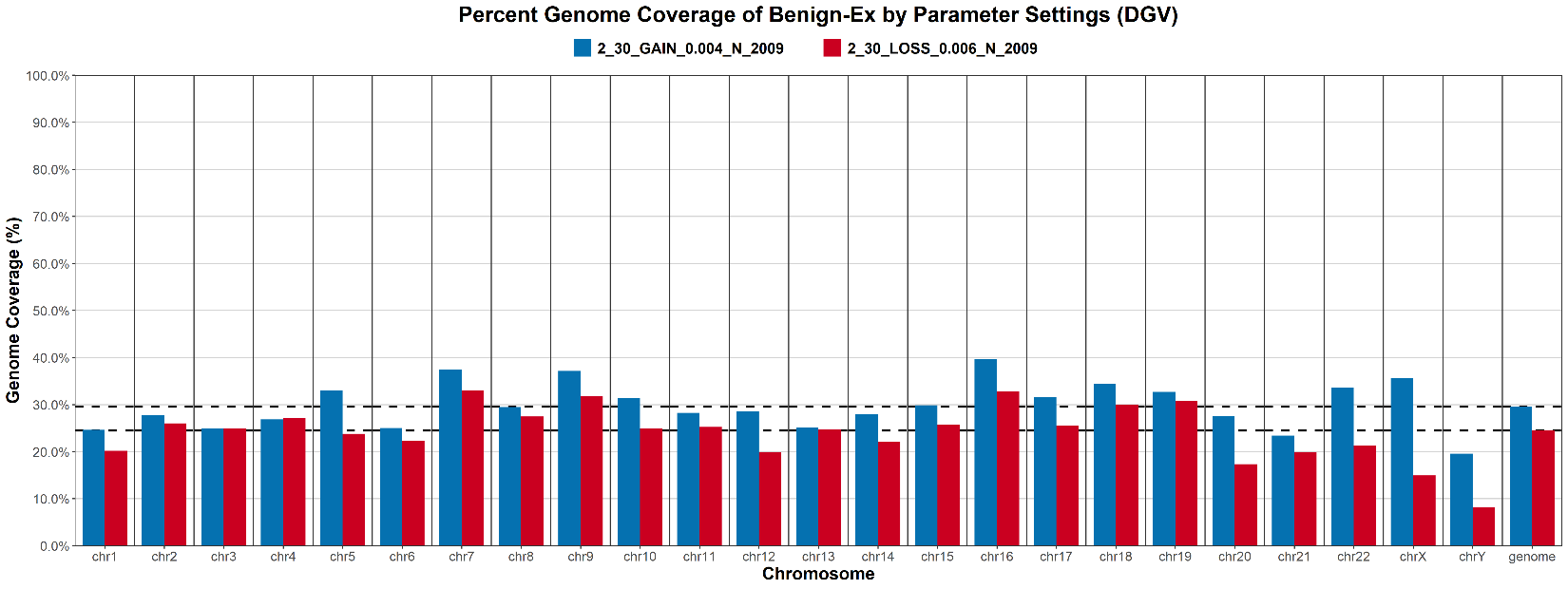

A

B

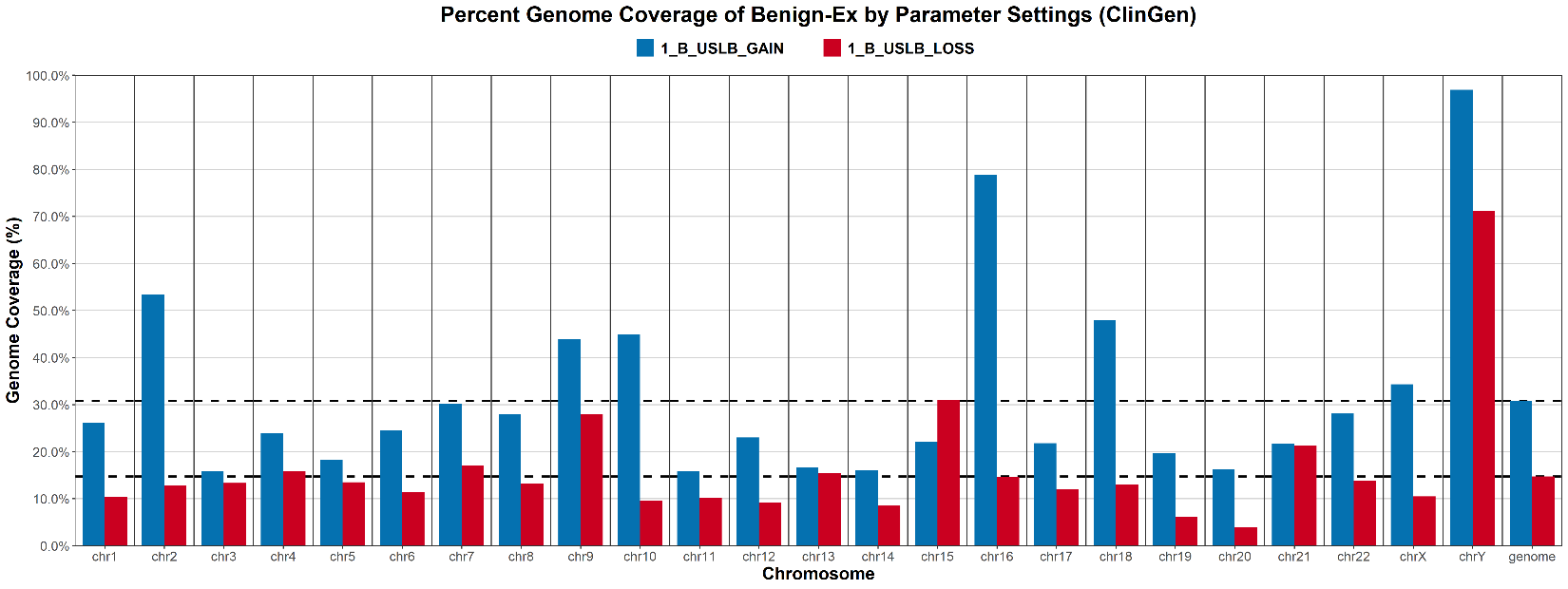

**Supplemental Figure 6: Spatial Correlation of Benign Regions: CNV Map**

**.** Spline graph depicting the spatial correlation between Zarrei *et al.*’s Stringent and Inclusive CNV Maps^24^ as assessed by the Bedtools reldist function for (A) gains and (B) losses.

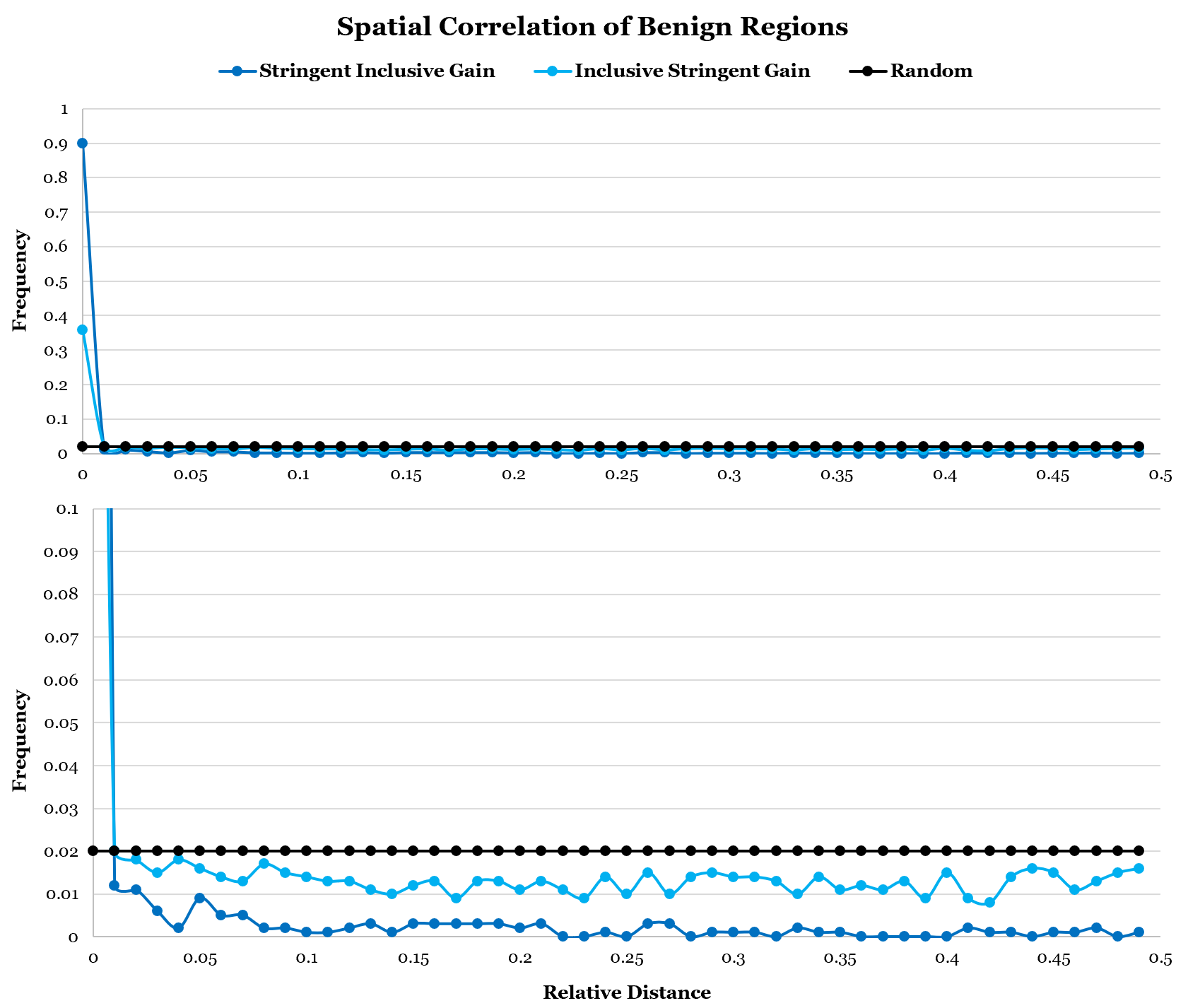

A

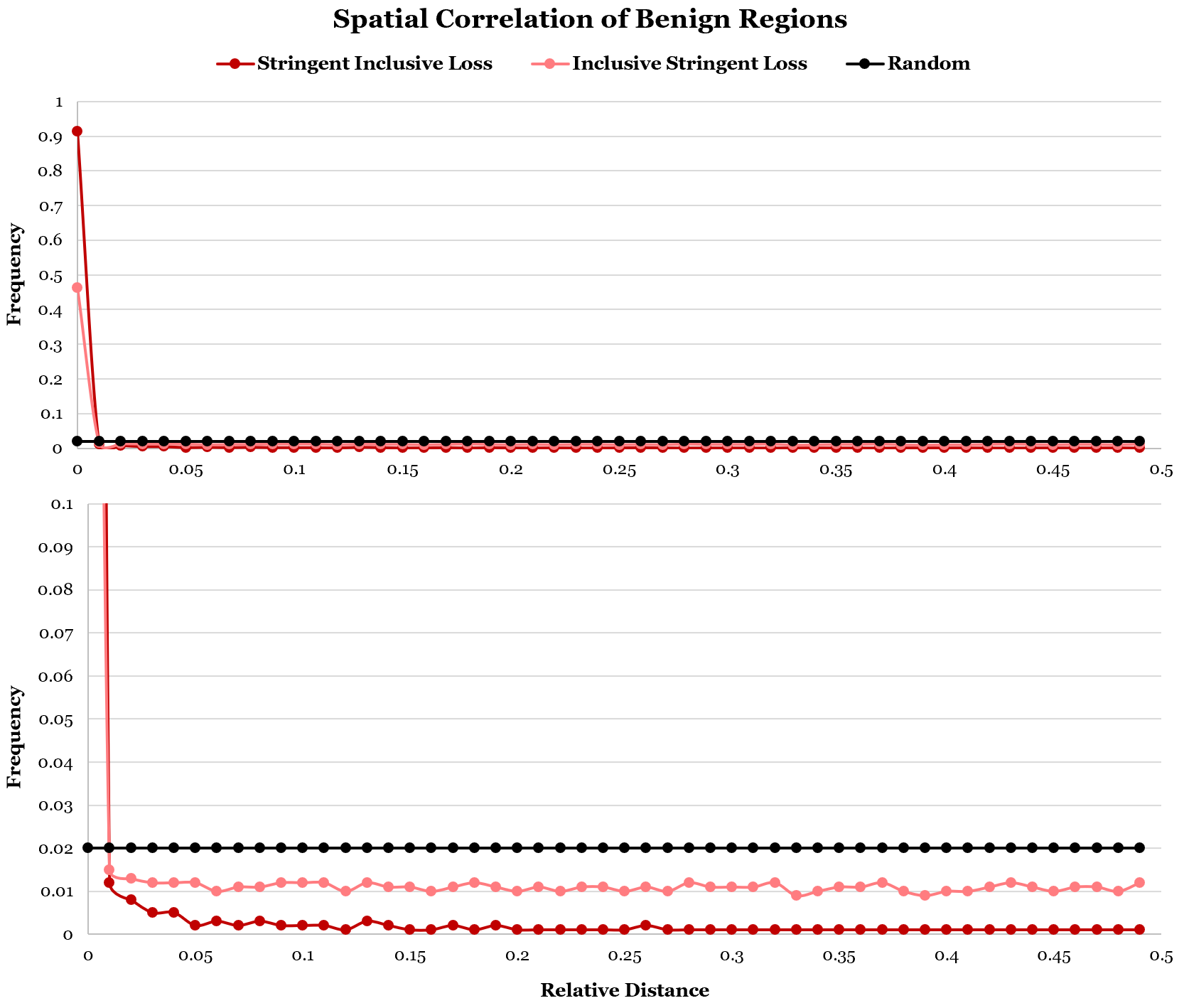

B

**Supplemental Figure 7: Spatial Correlation of Benign Regions: Benign-Ex DGV vs CNV Map**

**.** Spline graph depicting the spatial correlation between Benign-Ex’s DGV and ClinGen benign regions as assessed by the Bedtools reldist function for (A) DGV vs Stringent Gain; (B) DGV vs Stringent Loss; (C) DGV vs Inclusive Gain; and (D) DGV vs Inclusive Loss.

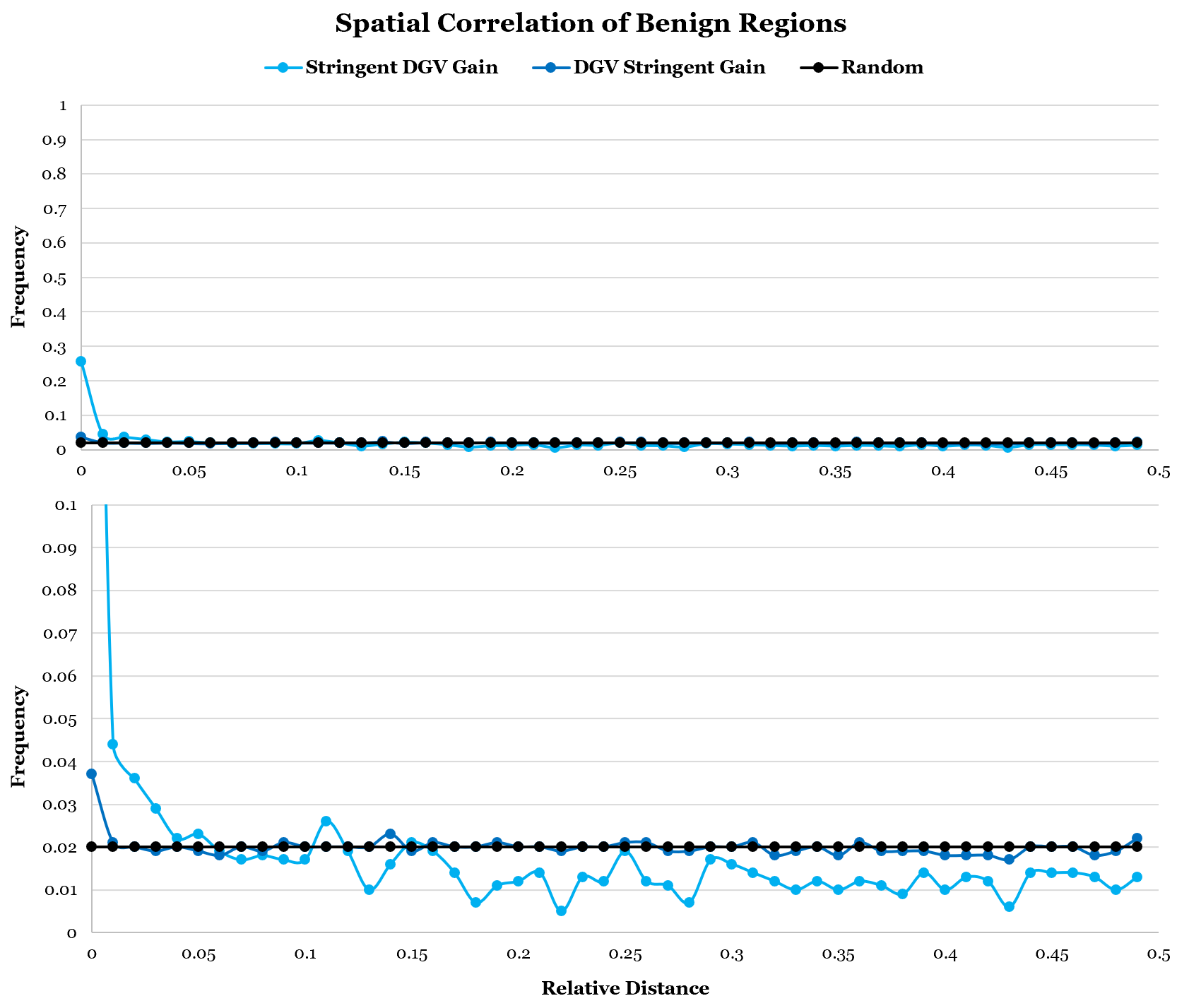

A

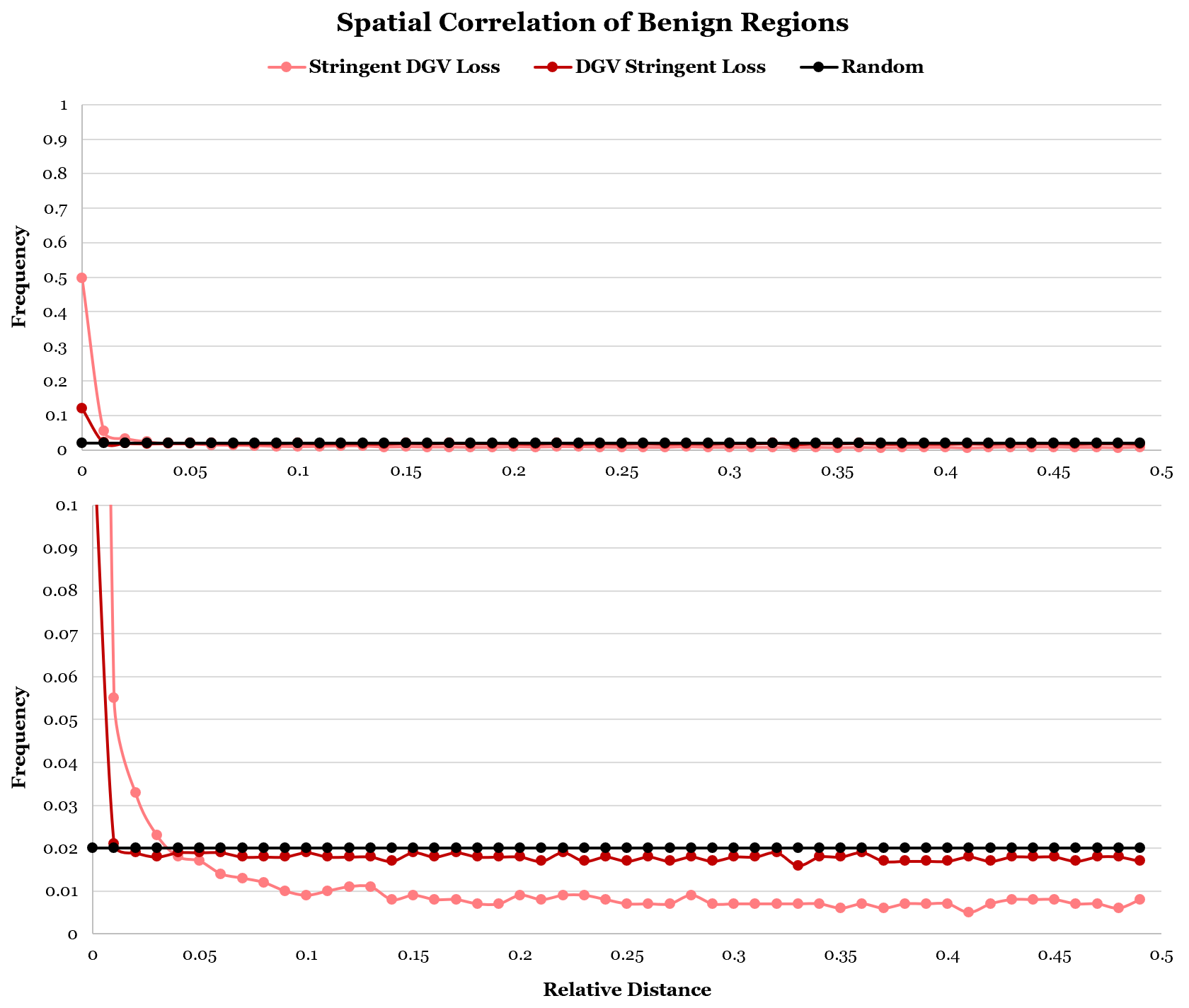

B

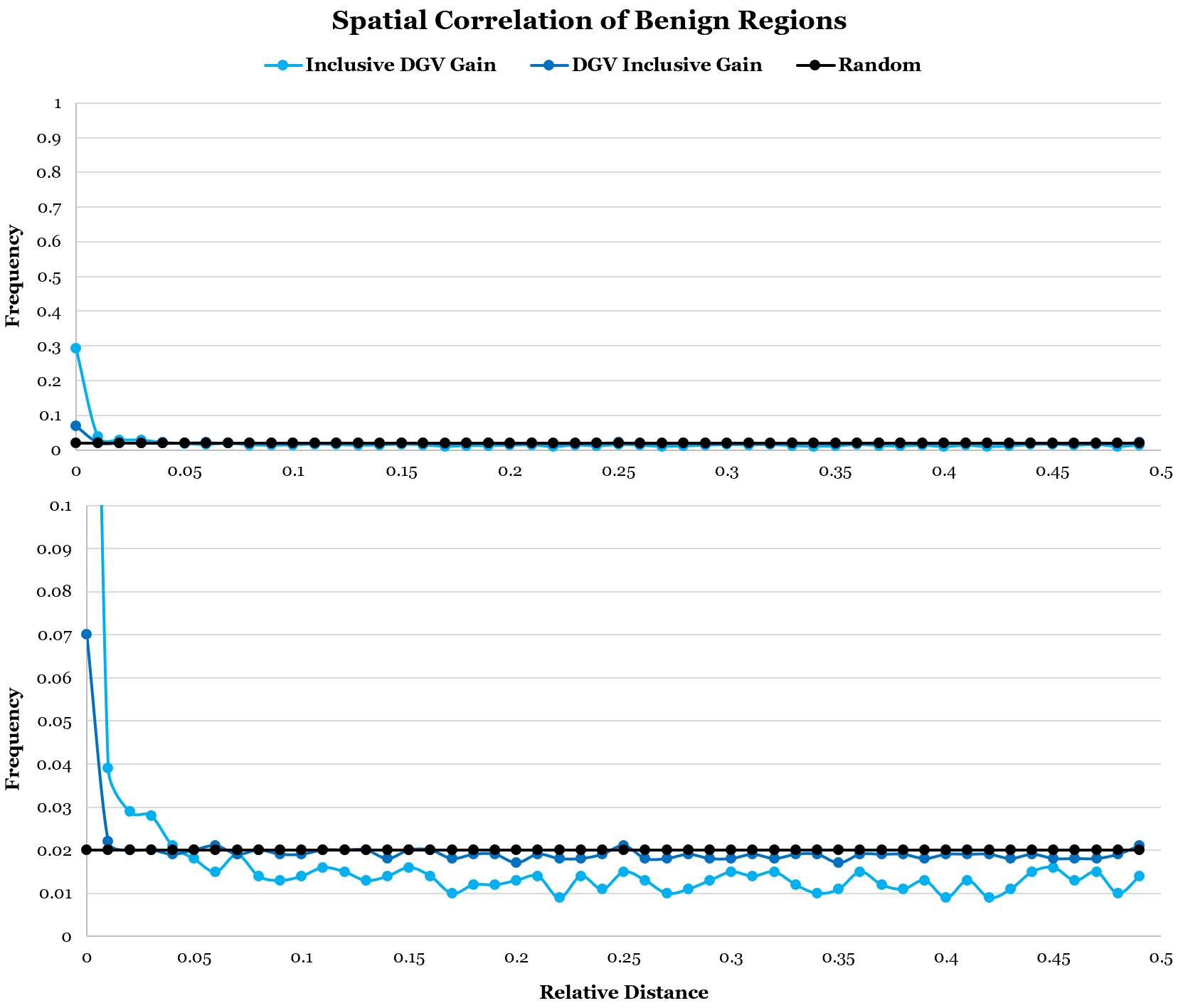

C

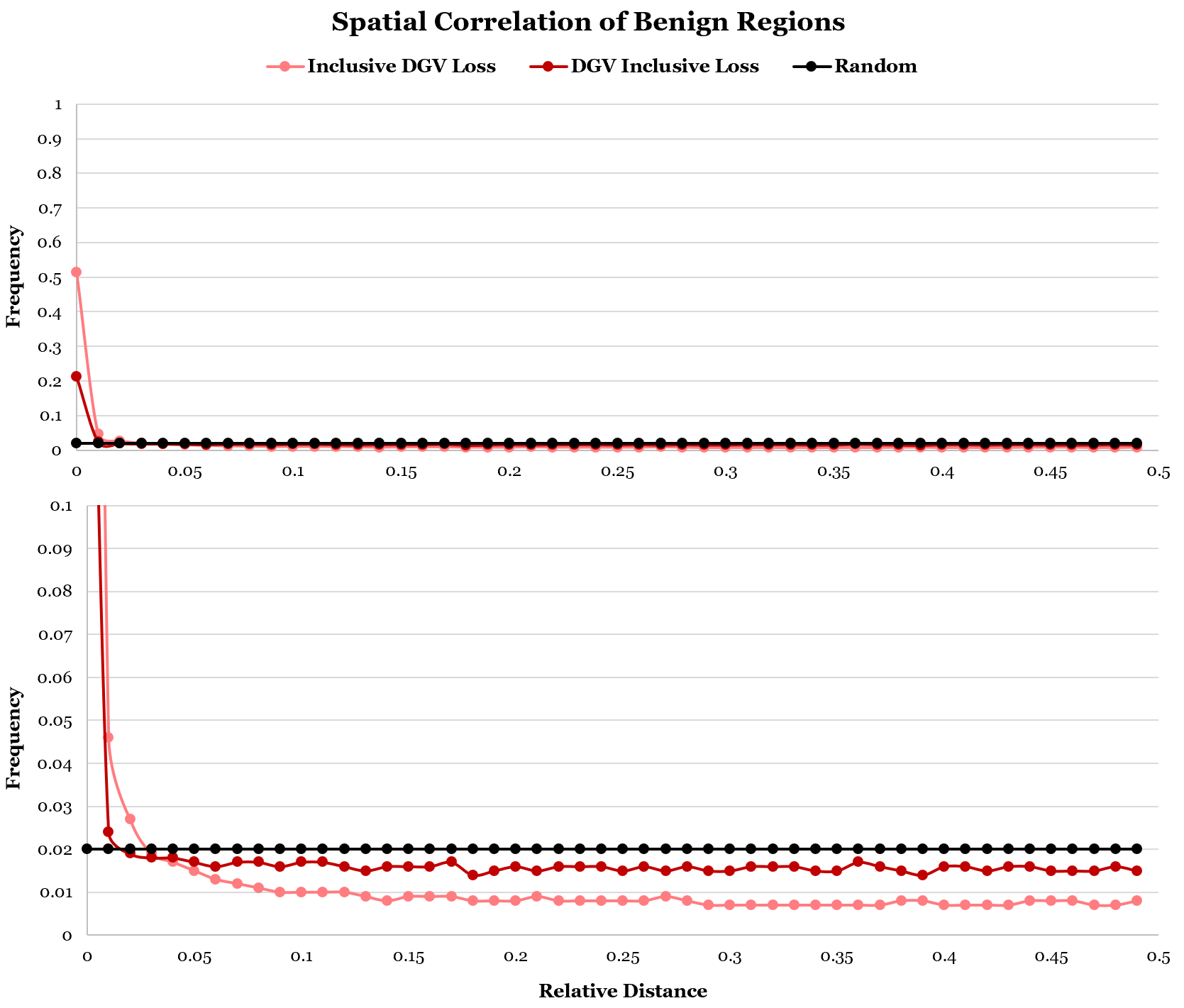

D

**Supplemental Figure 8: Comparative Size Distribution of Benign Regions**

**.** The size distribution of (A) gain and (B) loss benign regions called by Benign-Ex (top), and Zarrei *et al.’s* Inclusive (middle) and Stringent (bottom) CNV Maps ^24^. Vertical black lines represent the median benign region length.

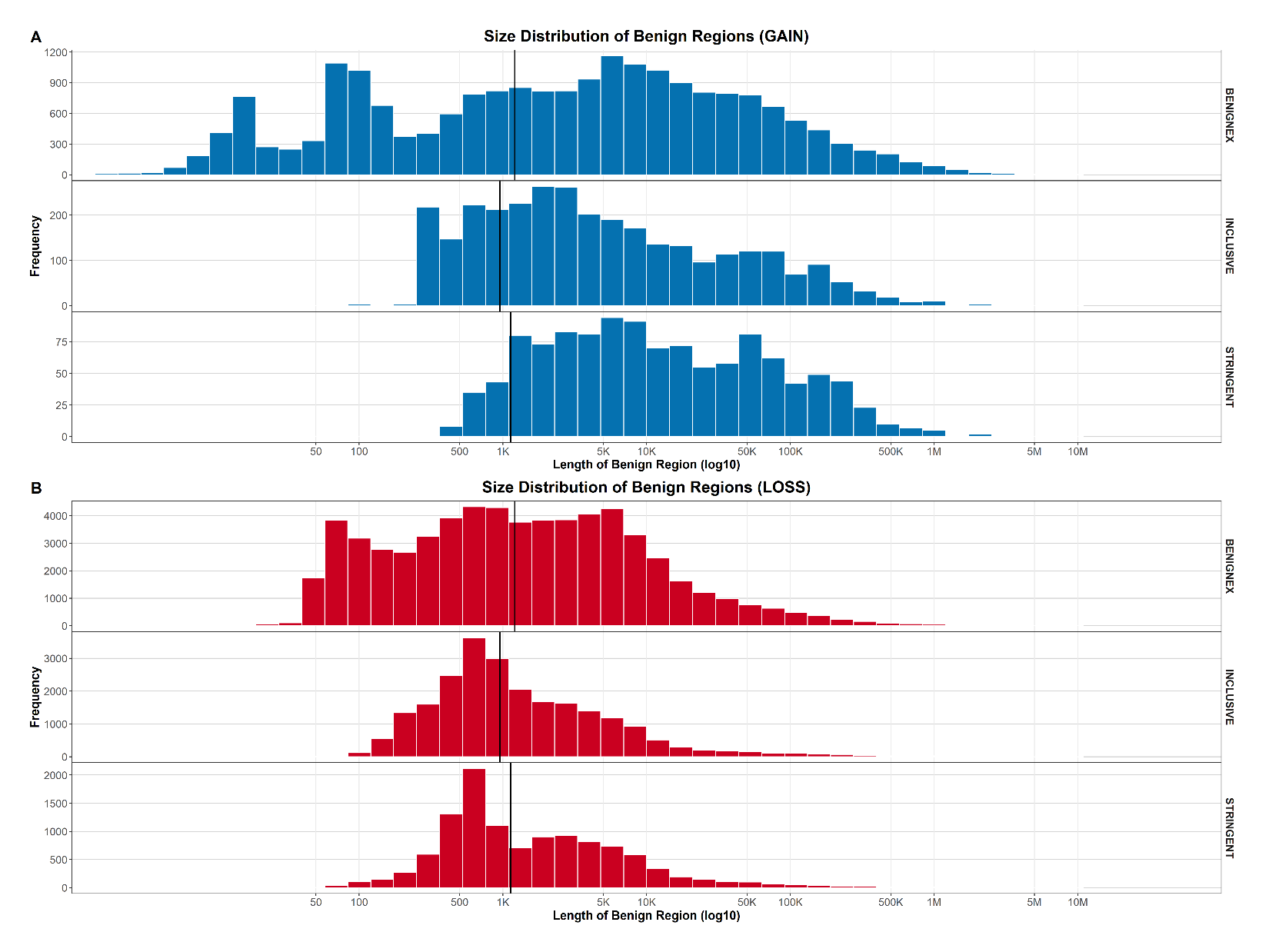

**Supplemental Figure 9: Percent Genome Called Benign by Benign-Ex by Parameter Setting (Final Simulation Set)**

**.** The percentage of the genome called benign for each parameter setting by Benign-Ex was tracked for the HG19 dataset for DGV and ClinGen. (A) The minimum number of events is indicated on the top, the allele frequency on the bottom, and the method and year filters are shown on the right. The top left box displays the percentage of the genome called benign for each allele frequency value with a minimum of 2 events and no method or year filter applied. (B) The minimum number of events is indicated on the bottom and the classification(s) on the top. The left box displays the percentage of the genome called benign for each minimum number of events wherein only benign variants were utilized.

A

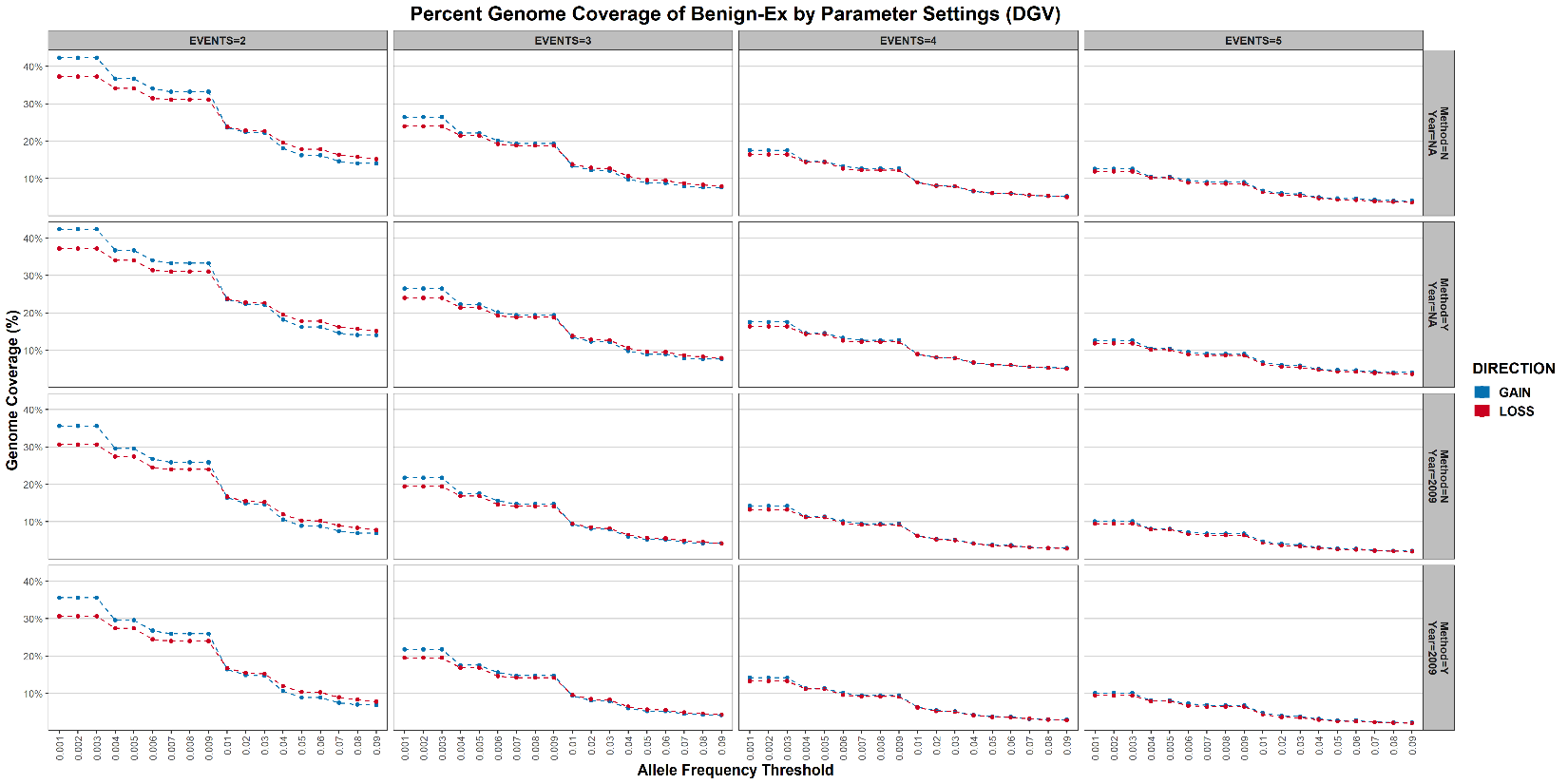

B

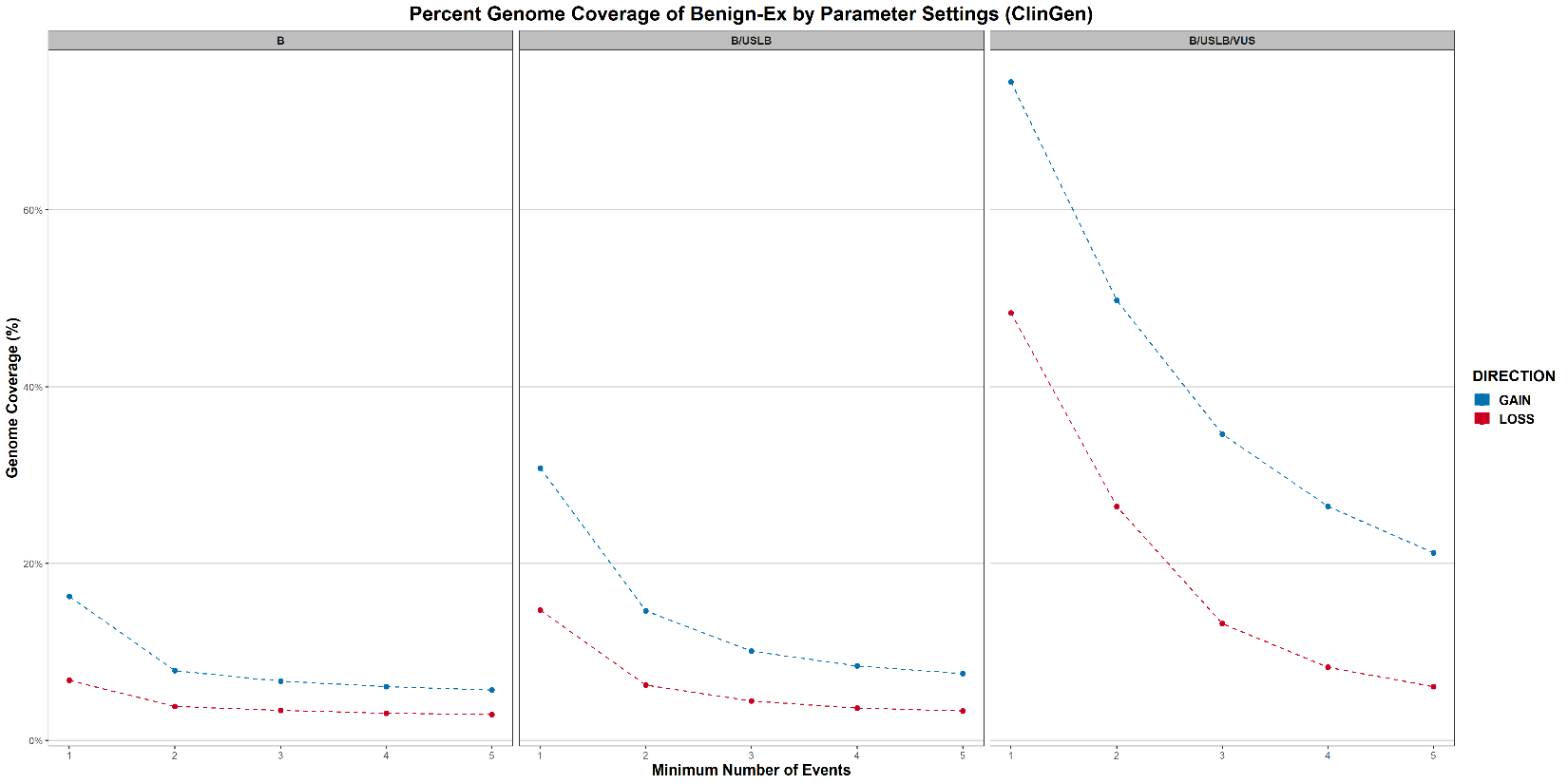

**Supplemental Figure 10: Benign-Ex Code Structure**

**.** Benign-Ex reads the AUTOMATION file, which determines which dataset(s) are being processed (DGV and/or CLINGEN). The corresponding ALGORITHM file is run for each chromosome * parameter combination specified by the PARAMETERS file. After all individual chromosome files have been created, the genome-wide parameter file is created, and the ONE_OVERLAP and HEATMAP_DATA codes are run. If the individual chromosome files do not need to be created (dashed line), the code will skip to ONE_OVERLAP and HEATMAP_DATA.

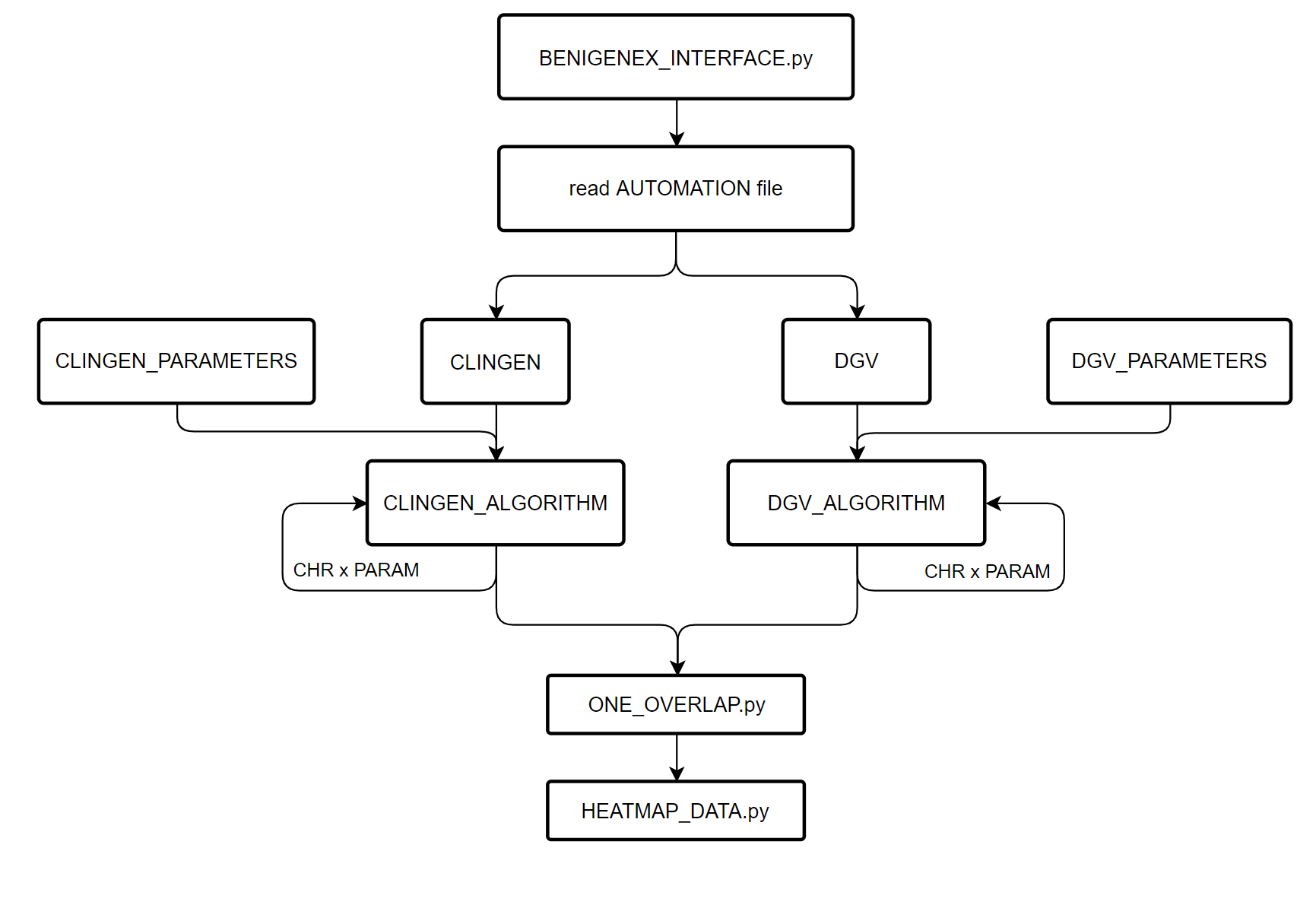

**Supplemental Figure 11: Distribution of Allele Frequencies within DGV**

**.** Frequency distribution of the number of gain (blue) and loss (red) variants in DGV at each allele frequency threshold bin. The cumulative percentage of variants accounted for at each allele frequency threshold is also shown.

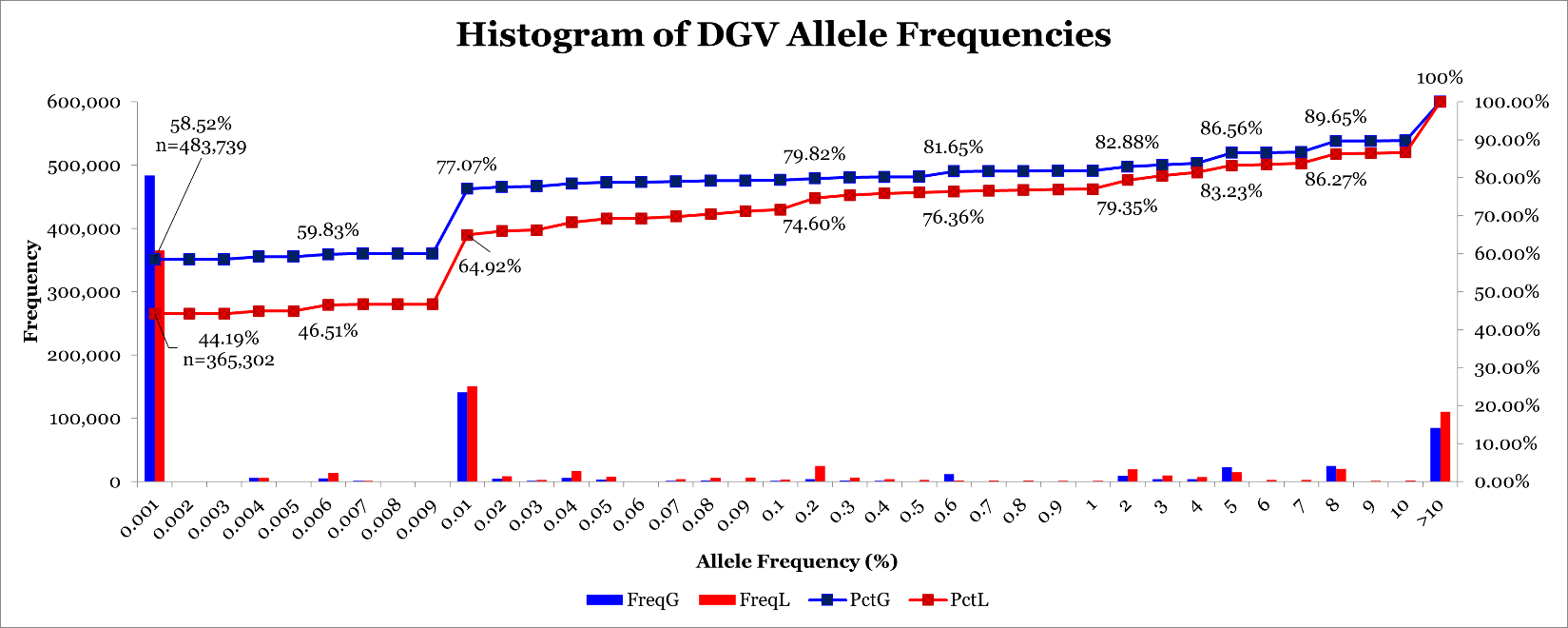

**Supplemental Figure 12: Preliminary HG19 Benign-Ex Heatmaps for DGV**

**.** To visually demonstrate how different Benign-Ex parameter sets perform across multiple pathogenic lists, Benign-Ex produces the following heatmap. The histogram on the top represents the average performance of that parameter set across all pathogenic lists. The maximum value is the optimal parameter setting across all pathogenic lists. The histogram on the right represents the average performance of all Benign-Ex parameter sets across that pathogenic list. The highest performing parameter set is indicated for each pathogenic list. The heatmap in the middle plots the final summary statistic for each pathogenic list + Benign-Ex parameter set combination. The Benign-Ex parameter simulations are sorted first by the minimum number of events, then left to right by the combination of the year and methodology filter applications: (N/2009, N/NA, Y/2009, Y/NA). Finally, the simulations are sorted by allele frequency (AF) from smallest to largest. Red indicates negative scores, Blue indicates positive scores, and White indicates a ‘0’ score. (A) All 1280 DGV Gain Simulations, (B) DGV Gain Simulations for Method=N; Year=NA (C) DGV Gain Simulations for Method=N; Year=2009, (D) DGV Gain Simulations for Method=Y; Year=NA; (E) DGV Gain Simulations for Method=Y; Year=2009; (F) All 1280 DGV Loss Simulations, (G) DGV Loss Simulations for Method=N; Year=NA (H) DGV Loss Simulations for Method=N; Year=2009, (I) DGV Loss Simulations for Method=Y; Year=NA; (J) DGV Loss Simulations for Method=Y; Year=2009.

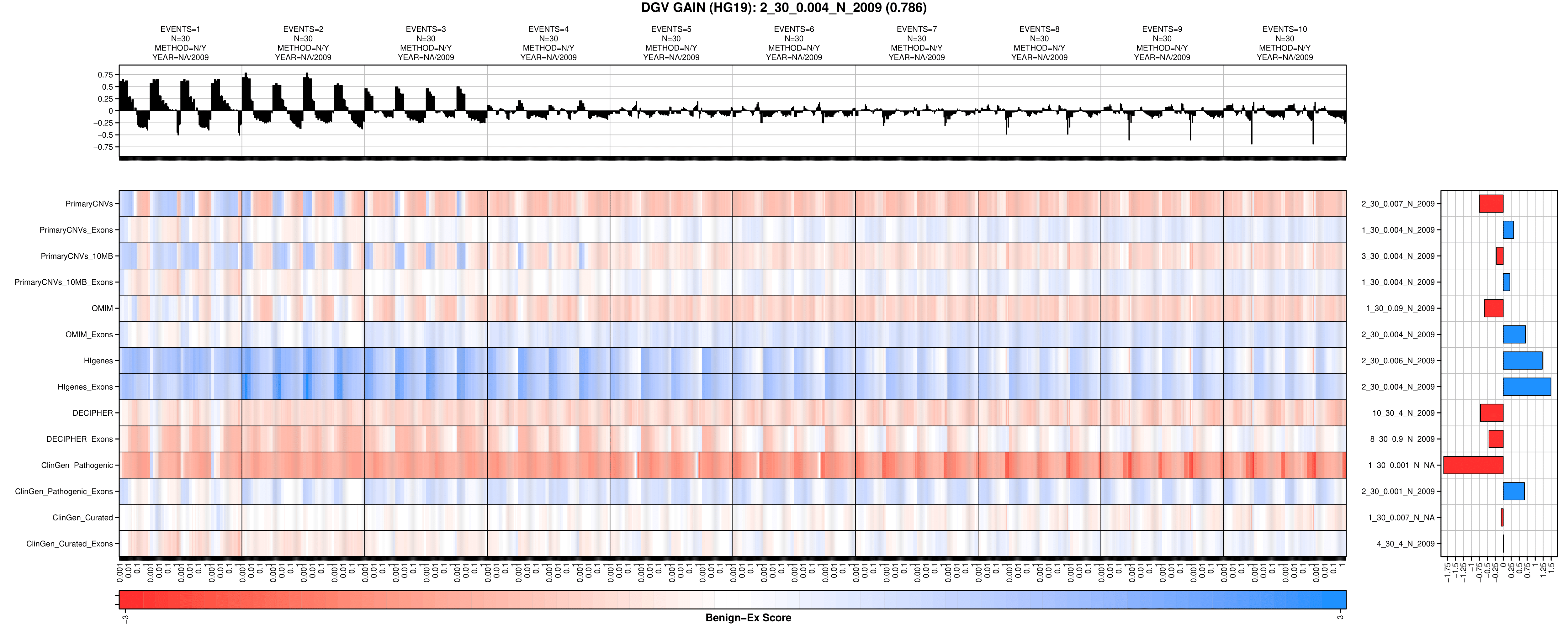

A

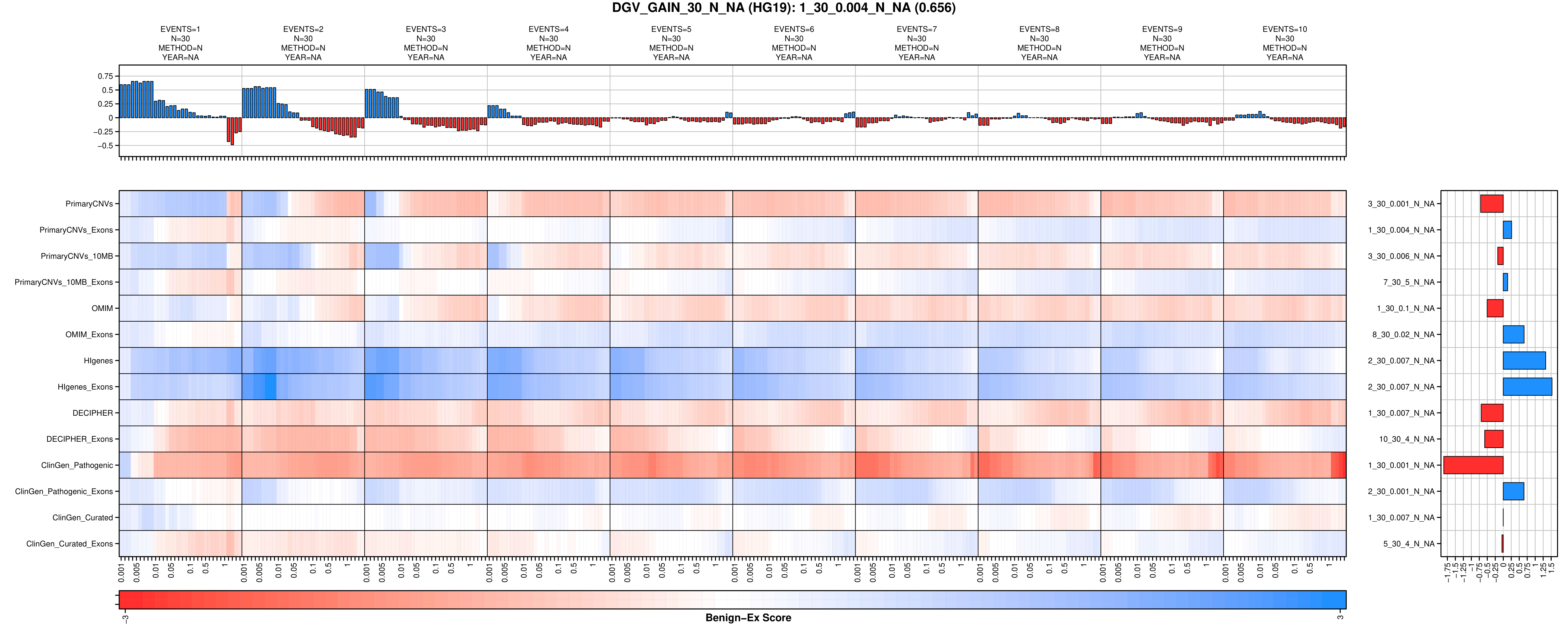

B

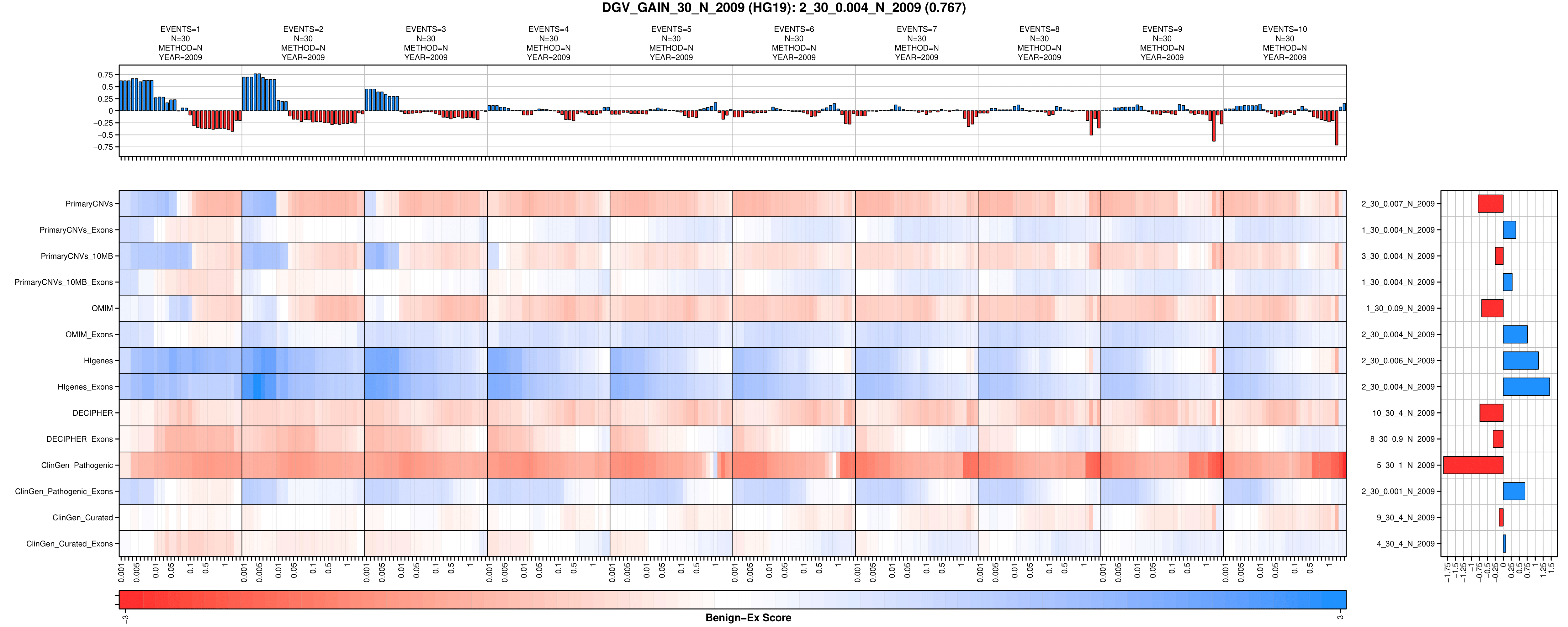

C

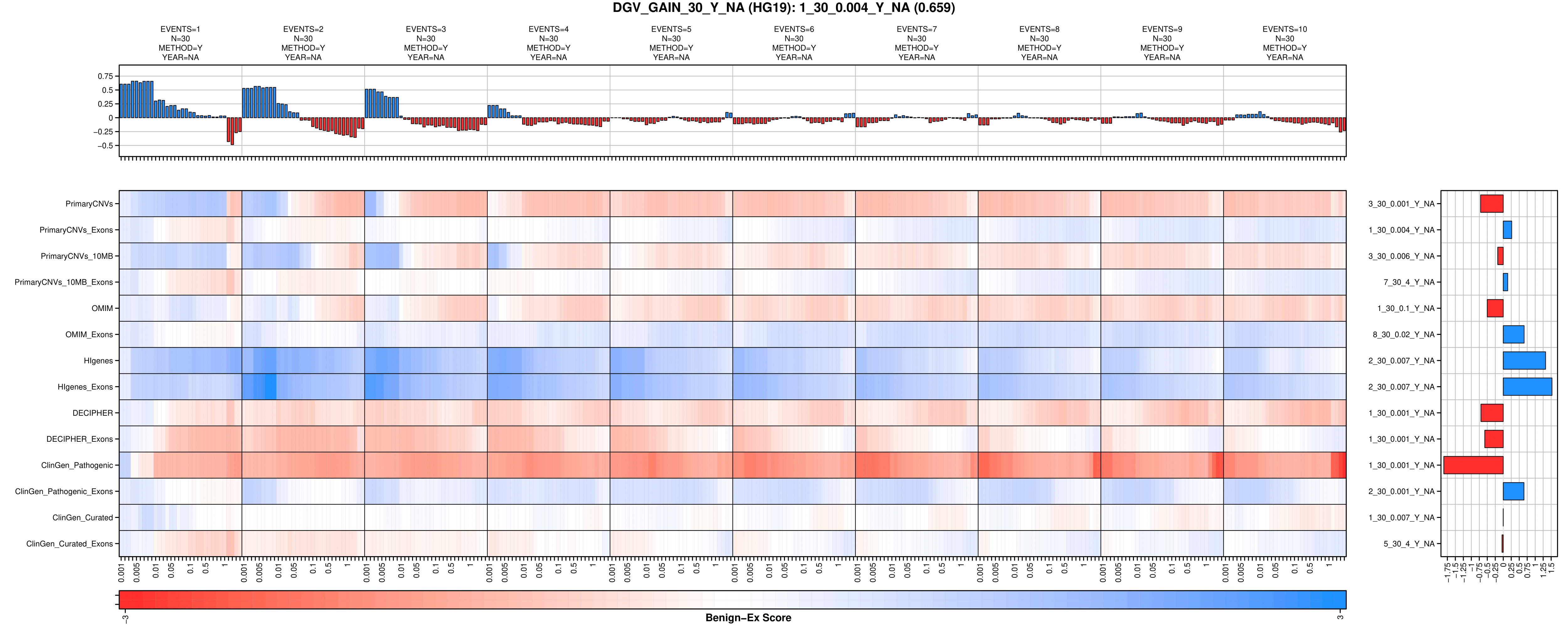

D

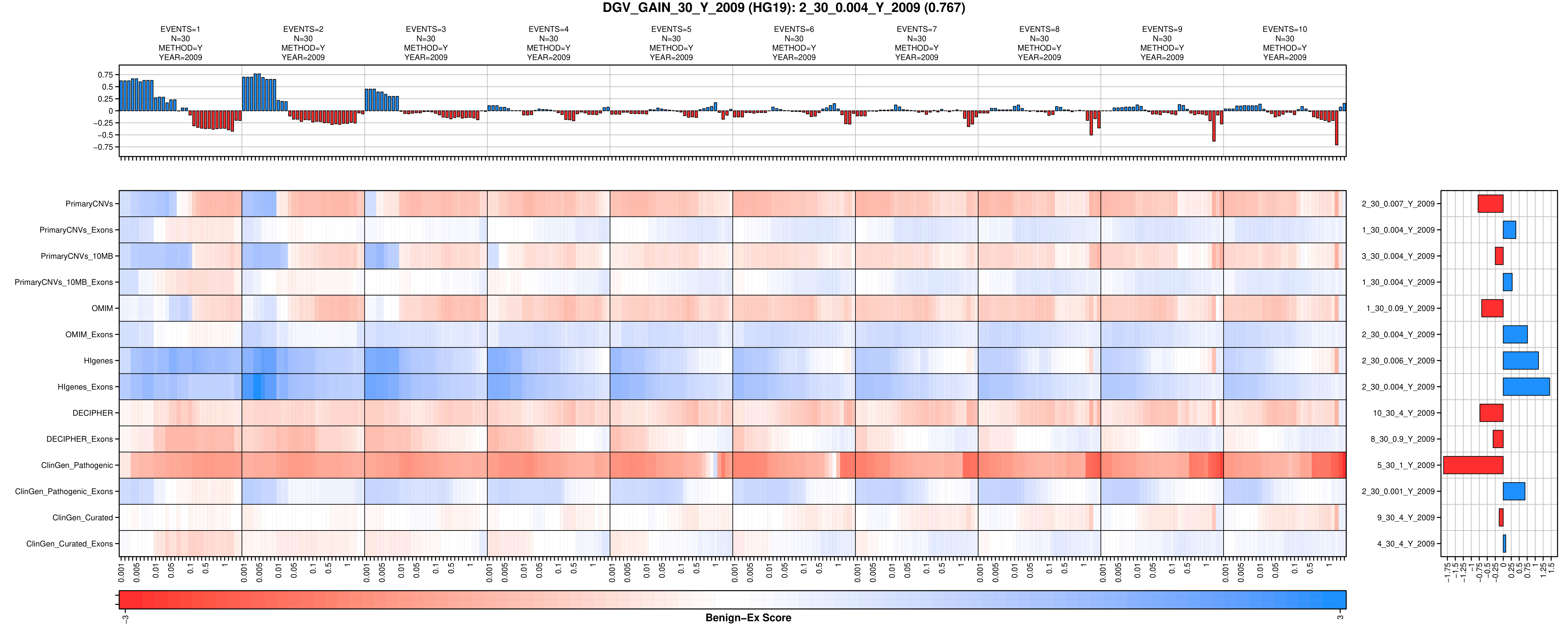

E

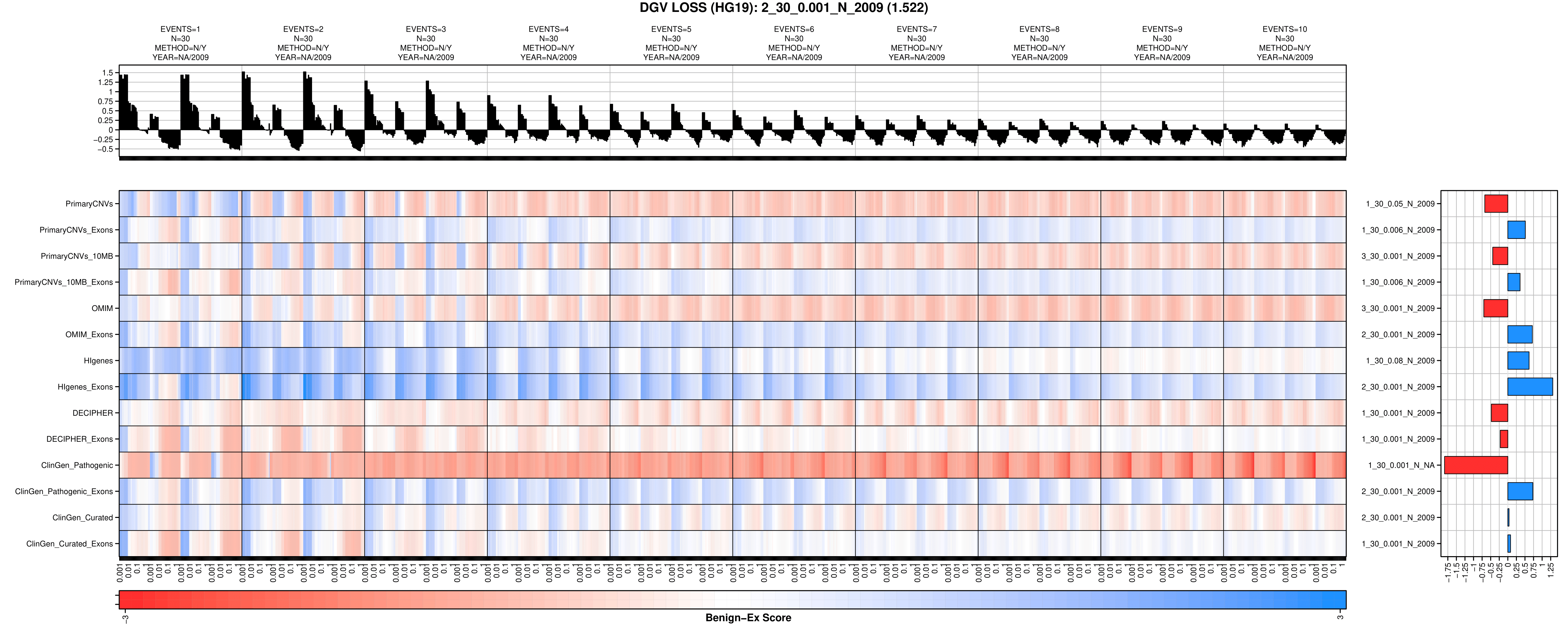

F

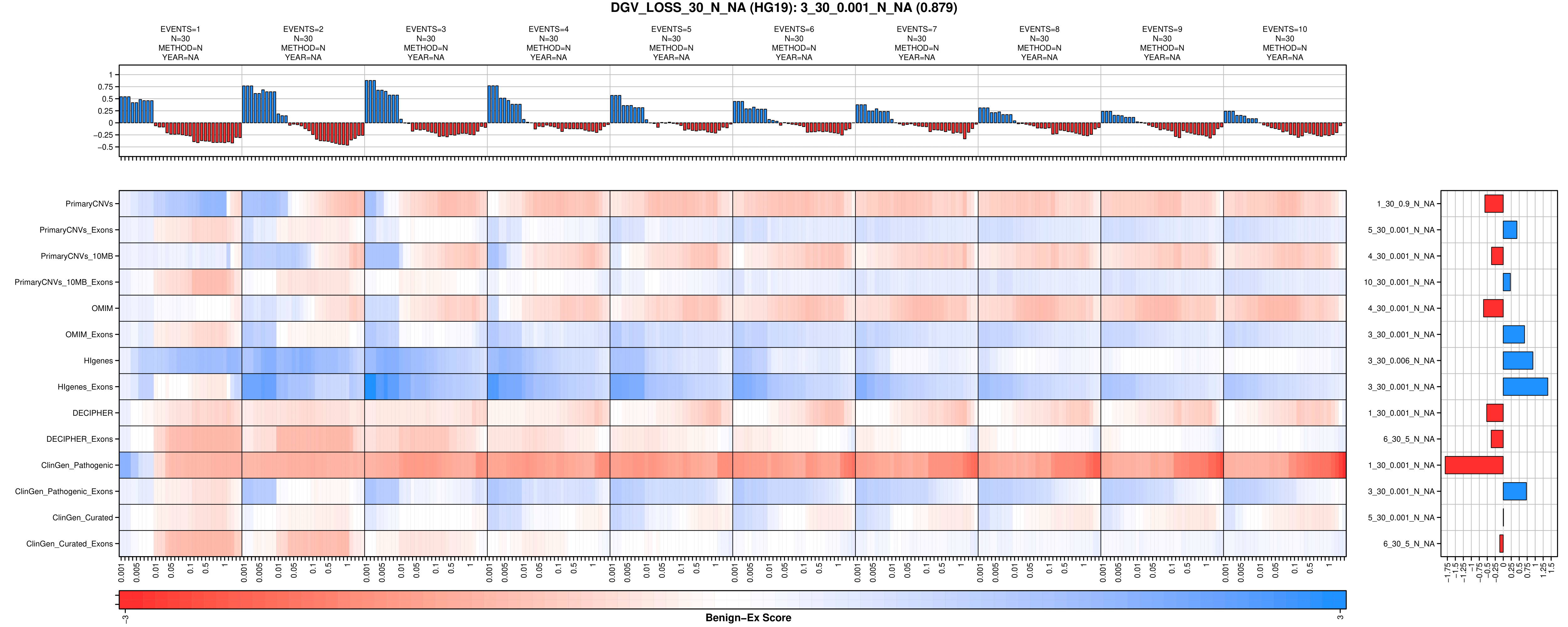

G

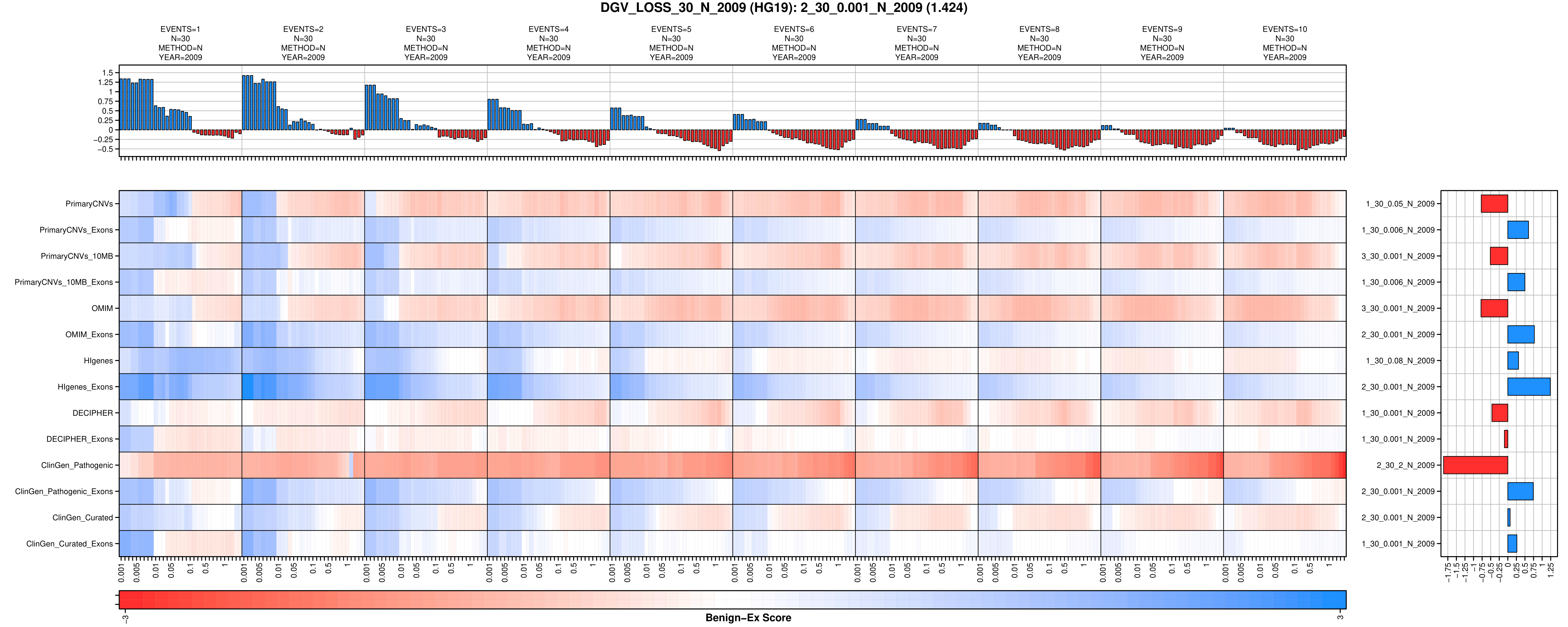

H

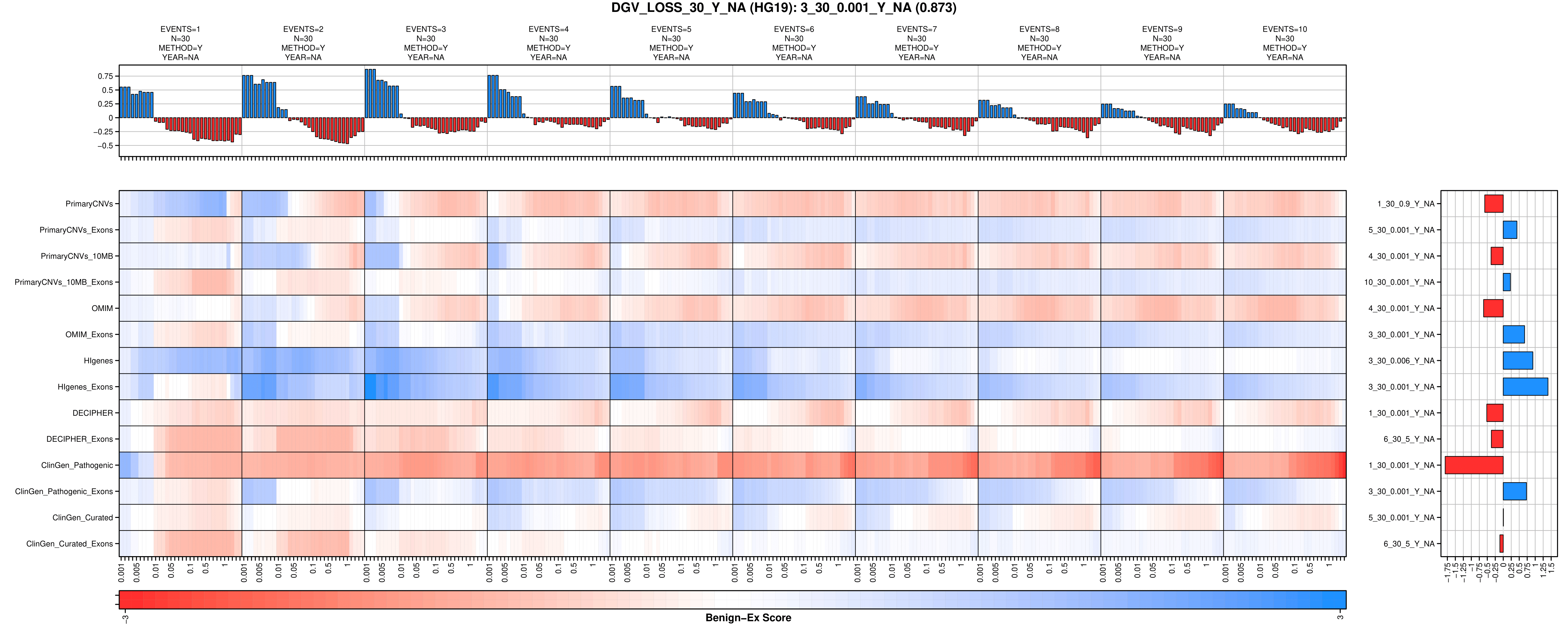

I

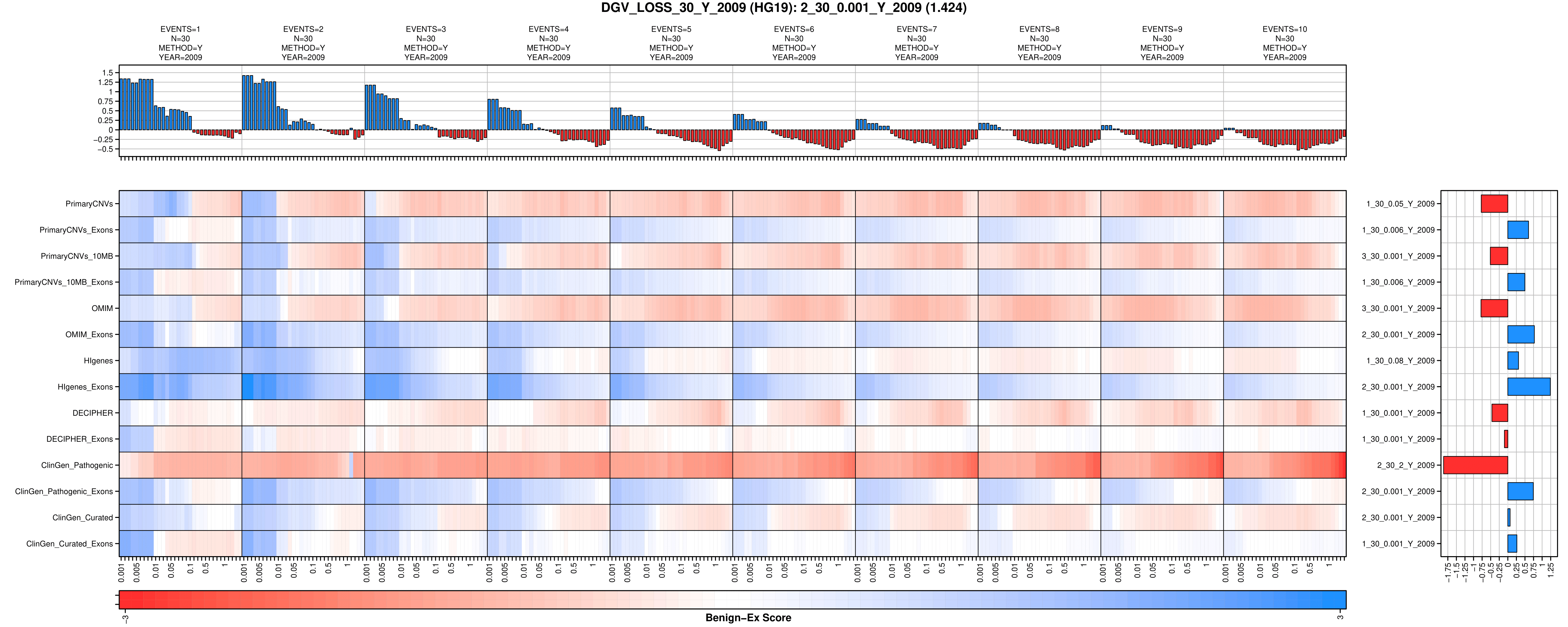

J

**Supplemental Figure 13: Preliminary HG19 Benign-Ex Heatmaps for ClinGen**

**.** To visually demonstrate how different Benign-Ex parameter sets perform across multiple pathogenic lists, Benign-Ex produces the following heatmap. The histogram on the top represents the average performance of that parameter set across all pathogenic lists. The maximum value is the optimal parameter setting across all pathogenic lists. The histogram on the right represents the average performance of all Benign-Ex parameter sets across that pathogenic list. The highest performing parameter set is indicated for each pathogenic list. The heatmap in the middle plots the final summary statistic for each pathogenic list + Benign-Ex parameter set combination. The Benign-Ex parameter simulations are sorted by the acceptable classifications (‘B’, ‘B/USLB’, ‘B/USLB/VUS’). Red indicates negative scores, Blue indicates positive scores, and White indicates a ‘0’ score. (A) All 60 ClinGen Gain Simulations, (B) All 60 ClinGen Loss Gain Simulations.

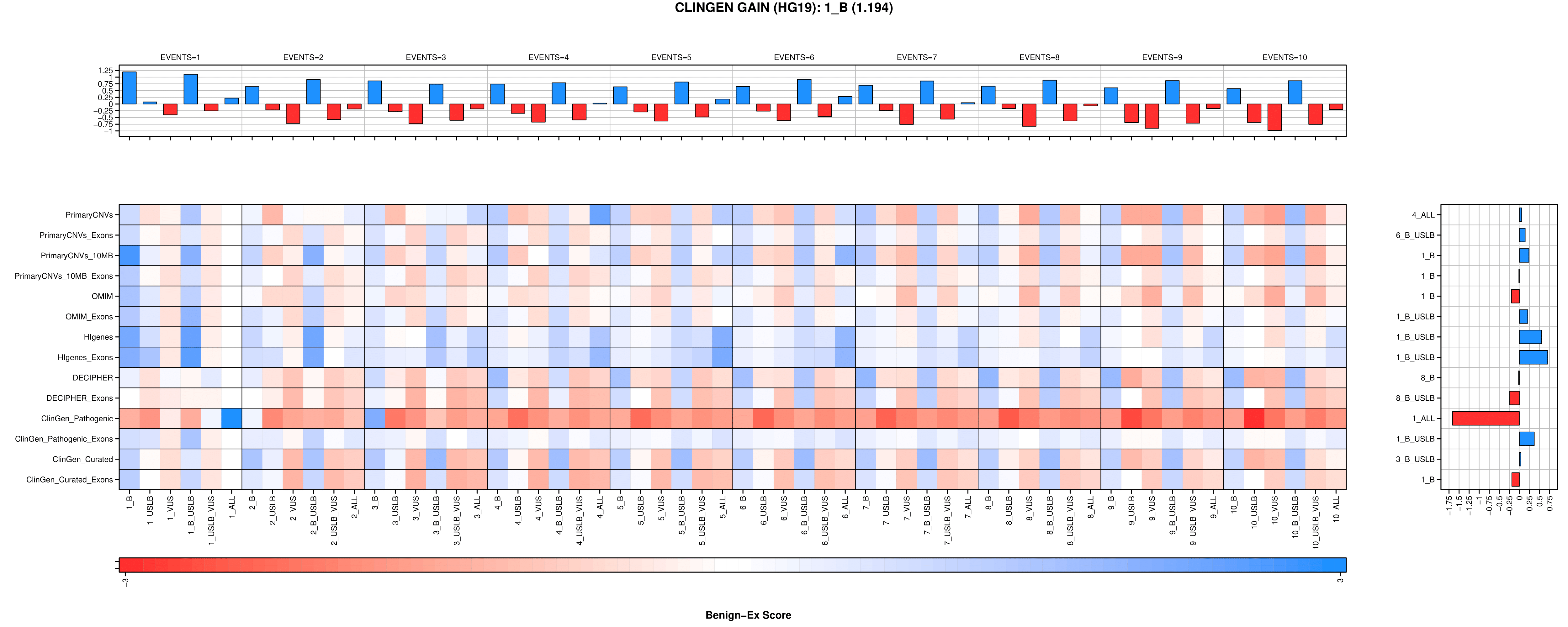

A

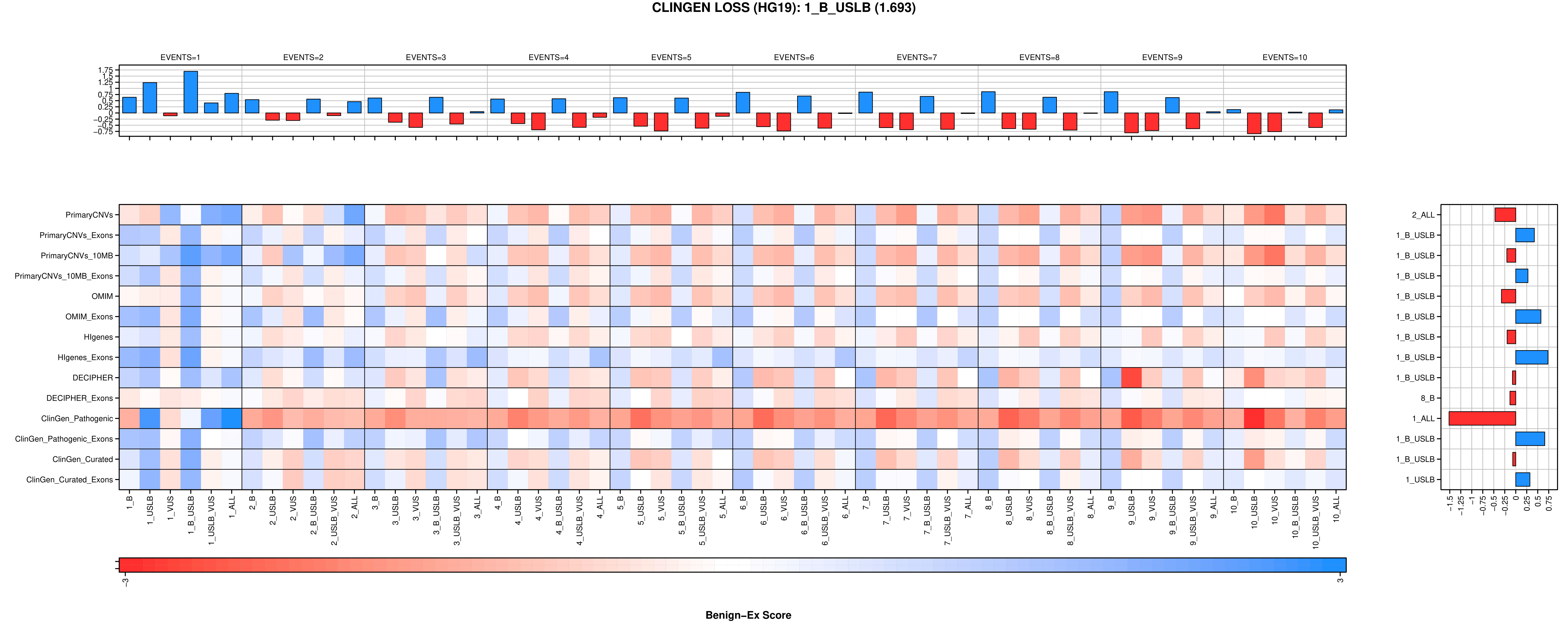

B

**Supplemental Figure 14: Distribution of DGV Events in Benign-Ex Called Benign Regions**

**.** Frequency distribution of the number of gain (blue) or loss (red) DGV entries which overlap each benign region identified by Benign-Ex. The percentage of benign regions retained at each threshold value is shown (*e.g.* 24.3% of the identified benign LOSS regions are supported by five or more DGV variants).

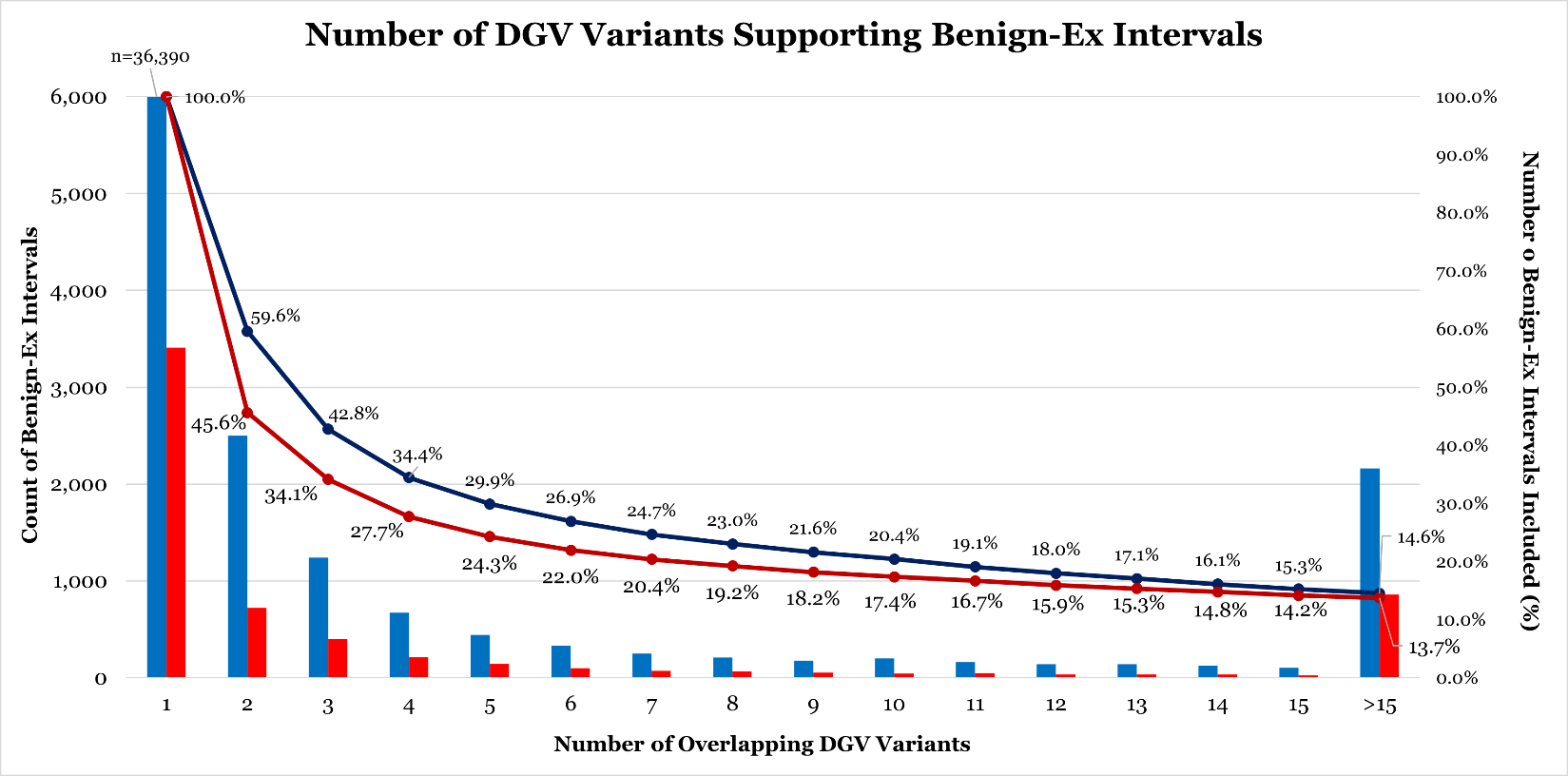

**Supplemental Figure 15: Distribution of Overlapping DGV Events: Tiled 500-bp Windows**

**.** Frequency distribution of the number of gain (blue) or loss (red) DGV entries which overlap a single 500-bp window. The percentage of 500-bp windows which are retained at each threshold is shown.

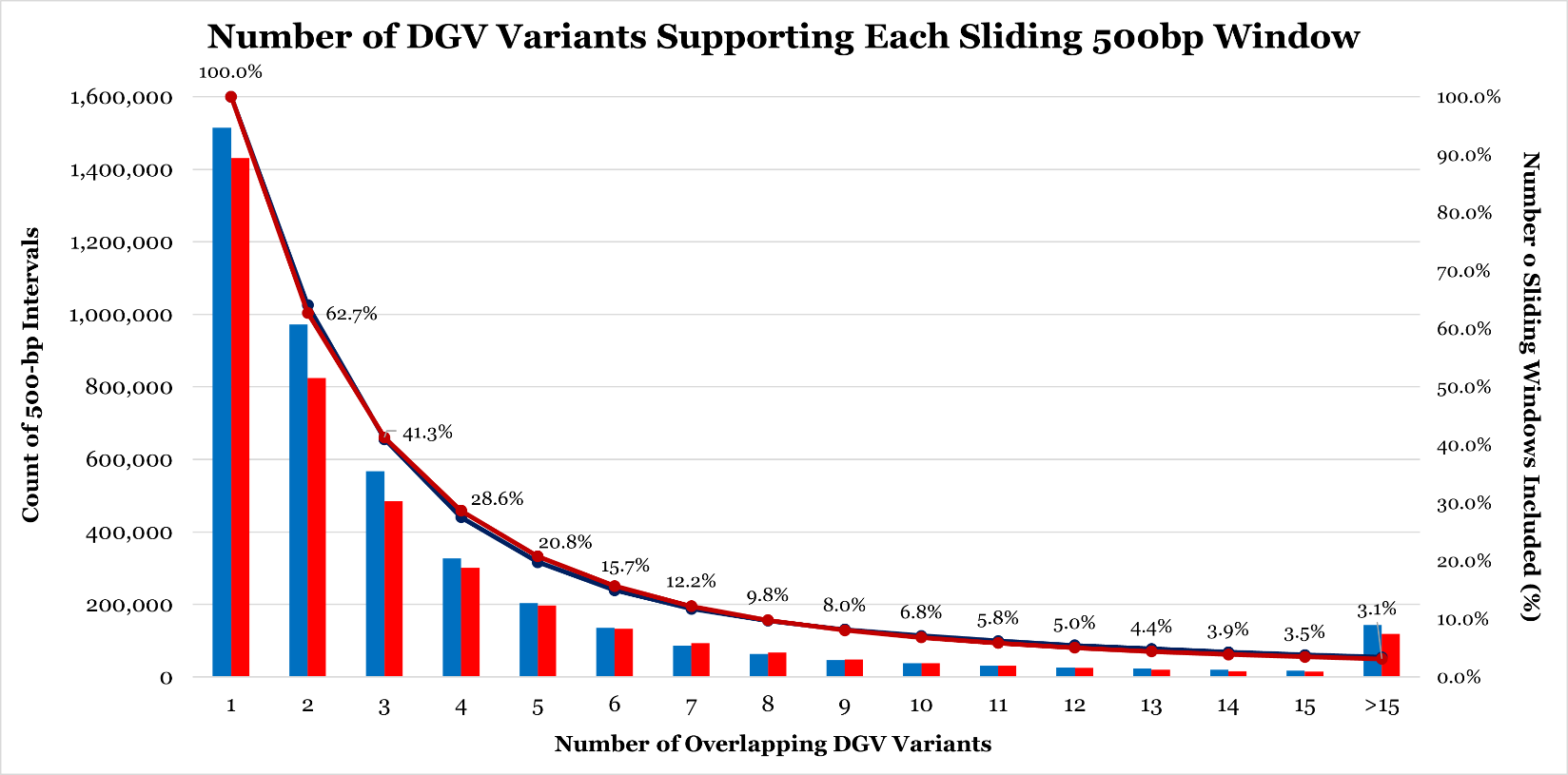

**Supplemental Figure 16: Distribution of ClinGen Events in Benign-Ex Called Benign Regions**

**.** Frequency distribution of the number of gain (blue) or loss (red) DGV entries which overlap each benign region identified by Benign-Ex. The percentage of benign regions retained at each threshold value is shown (*e.g.* 24.3% of the identified benign LOSS regions are supported by five or more DGV variants).

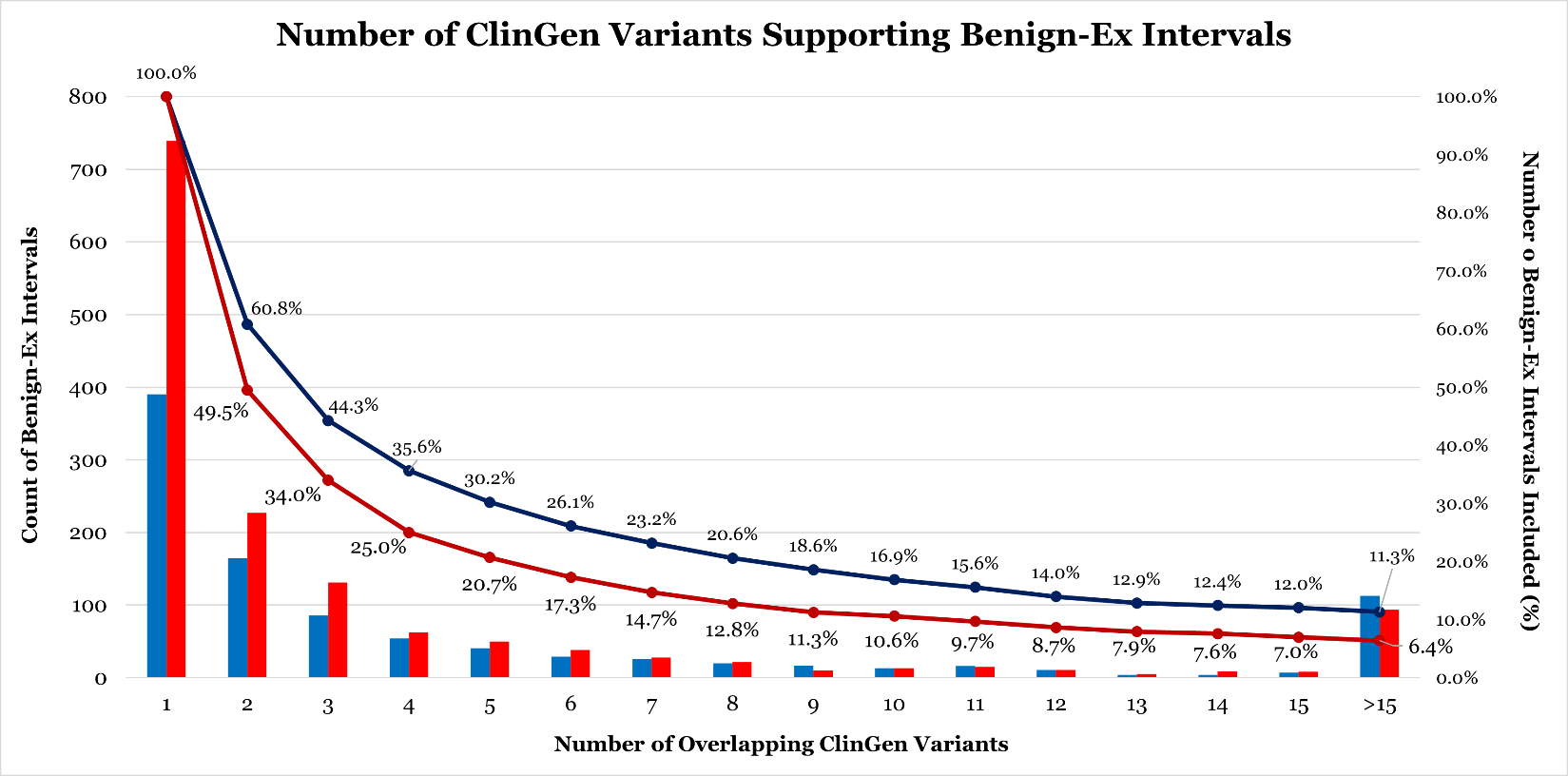

**Supplemental Figure 17: Distribution of Overlapping ClinGen Events: Tiled 500-bp Windows**

**.** Frequency distribution of the number of gain (blue) or loss (red) ClinGen variants entries that overlap a single 500-bp window. The percentage of 500-bp windows which are retained at each threshold is shown.

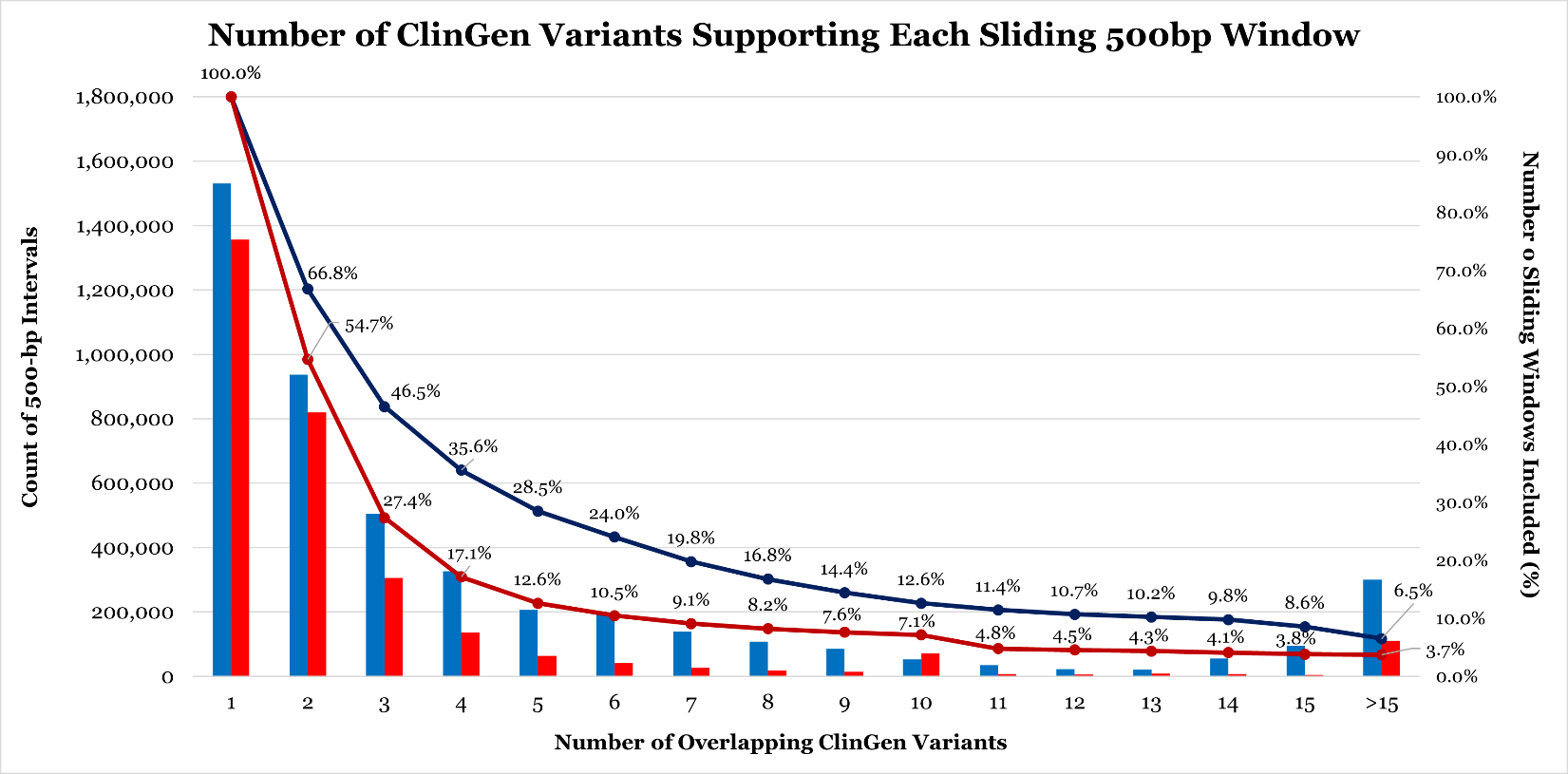

**Supplemental Figure 18: Time Taken to Generate ClinGen Benign Intervals by Parameter Setting and Chromosome Combination**

**.** The time taken to generate the individual chromosome file for each ClinGen parameter setting was tracked for the HG19 dataset. Individual chromosomes are indicated on the top, the corresponding parameter setting group is shown on the right, and the minimum number of events is shown on the bottom. Therefore, the top left box displays the amount of time it took for Benign-Ex to generate the chromosome 1 benign interval file for ‘ALL’ events in the ‘GAIN’ direction for different thresholds of the minimum number of events.

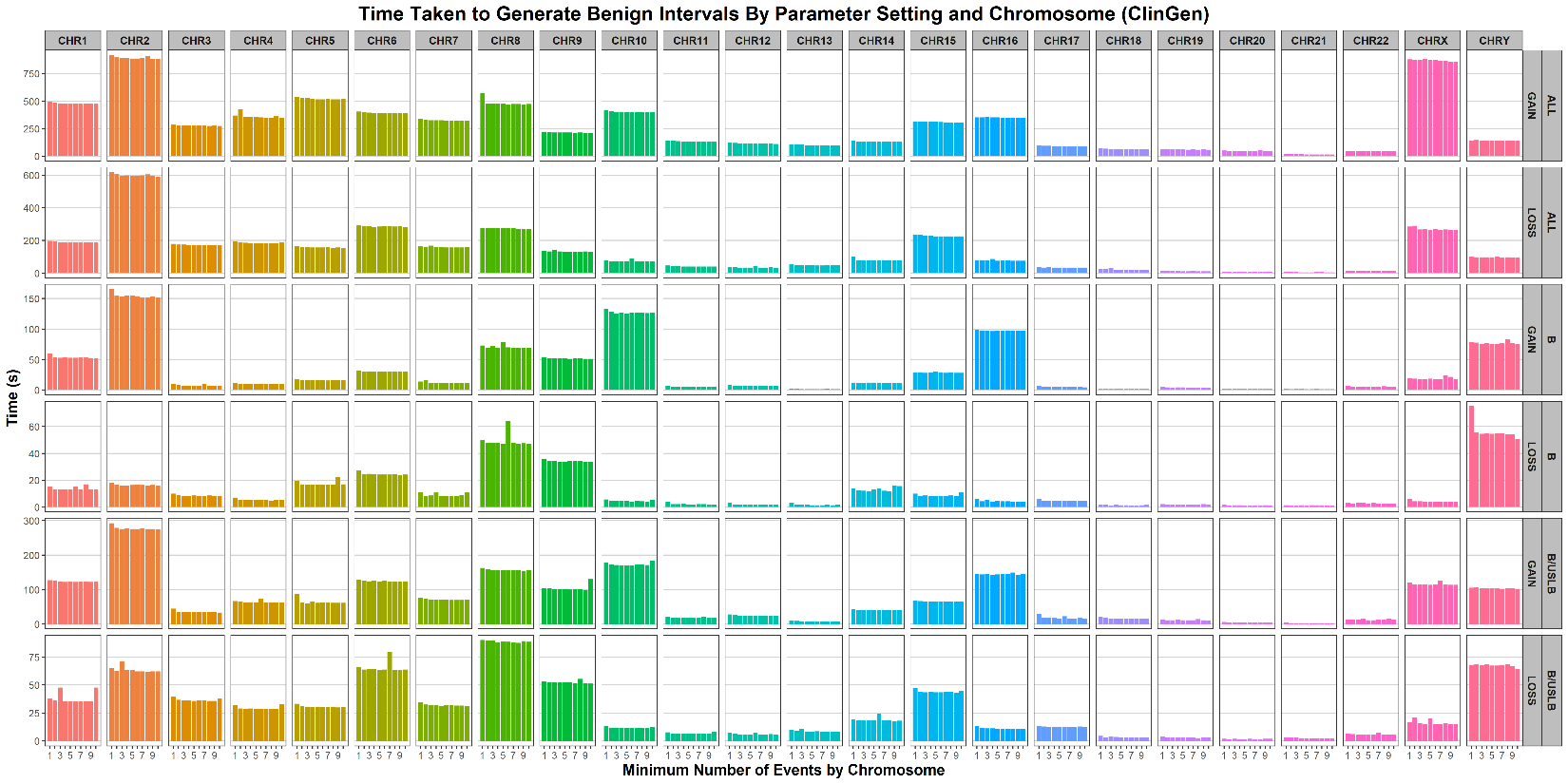

**Supplemental Figure 19: Time Taken to Generate DGV Benign Intervals by Parameter Setting and Chromosome Combination**

**.** The total time to generate the individual chromosome file for each DGV parameter setting was tracked for the HG19 dataset. Individual chromosomes are indicated on the top, the corresponding parameter setting group is indicated on the right, and the minimum number of events is indicated on the bottom. Therefore, the top left box displays the amount of time it took for Benign-Ex to generate the chromosome 1 benign interval file for ‘GAIN’ events identified from studies published in or after ‘2009’ for different thresholds of the minimum number of events.

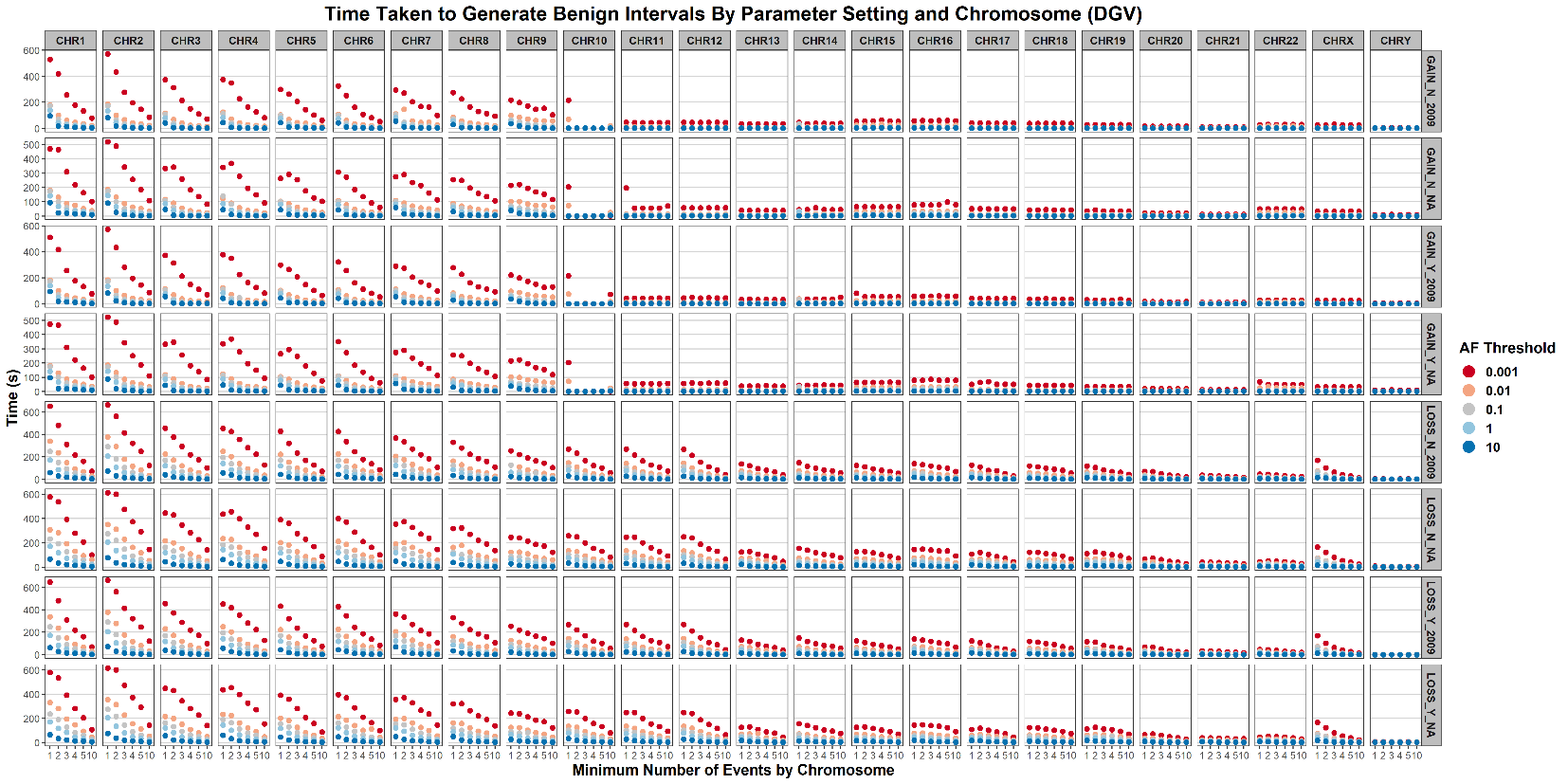

**Supplemental Figure 20: Time Taken to Generate DGV Benign Intervals by Parameter Setting and Chromosome Combination for Gain Events**

**.** The total time taken to generate the individual chromosome file for each DGV parameter setting with the following constraints (Direction = “Gain”; Minimum Number of Events = 1) was tracked for the HG19 dataset. Individual chromosomes are indicated on the top, the minimum allele frequency is indicated on the right, and the parameter setting grouping is indicated on the bottom. Therefore, the top left box displays the amount of time it took for Benign-Ex to generate the chromosome 1 benign interval file for ‘GAIN’ events, with a 0.001 minimum allele frequency, for each of the four parameter setting groupings.

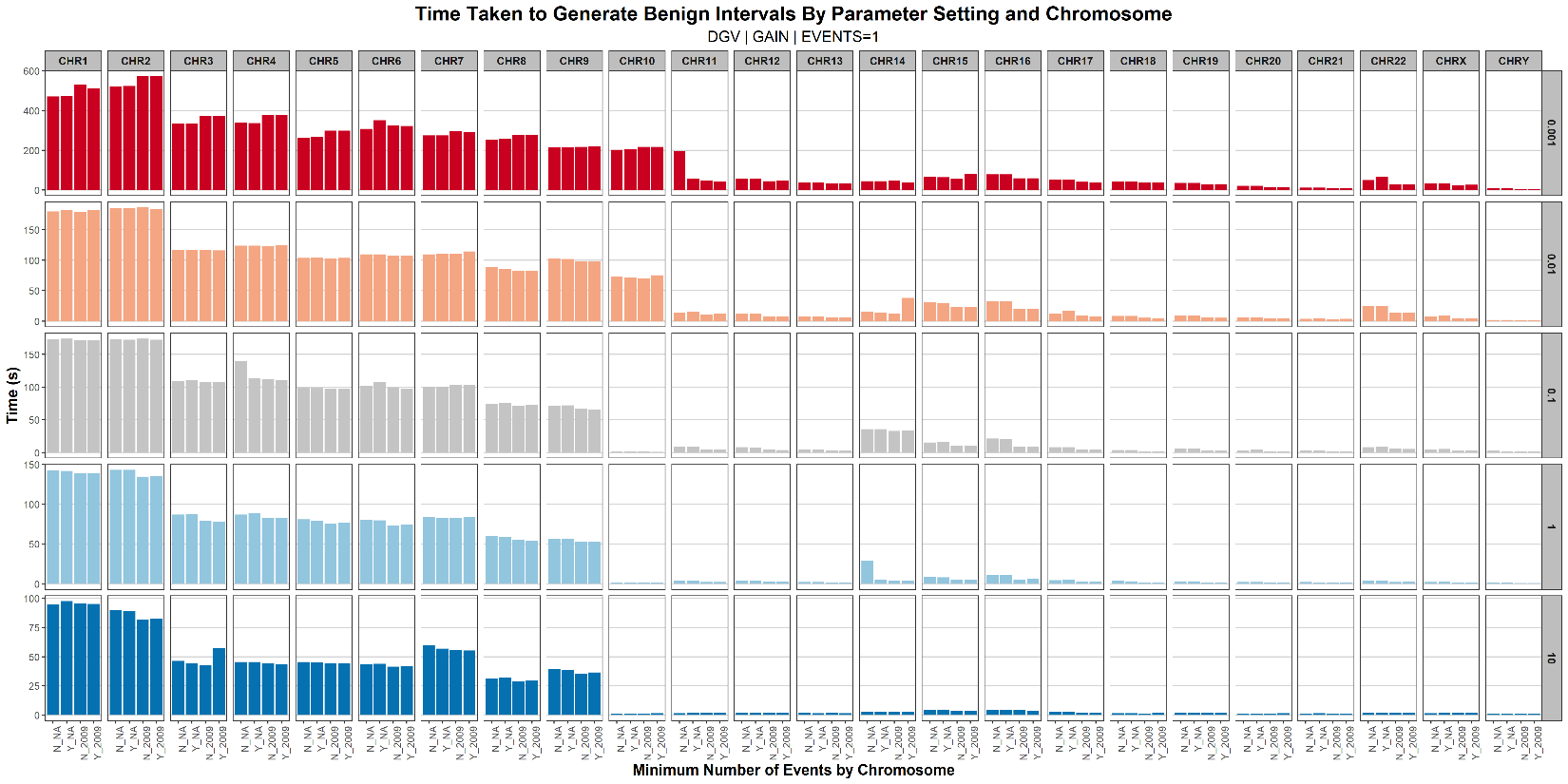

**Supplemental Figure 21: Number of Intervals to be Evaluated by Benign-Ex by Chromosome and Parameter Setting**

**.** The total number of GAIN intervals evaluated by Benign-Ex was tracked for the HG19 dataset. Individual chromosomes are indicated on the top, the minimum number of events is indicated on the right, and the allele frequency threshold is indicated on the bottom. The top left box displays the total number of intervals evaluated for all DGV variants on chromosome 1 that meet the allele frequency threshold.

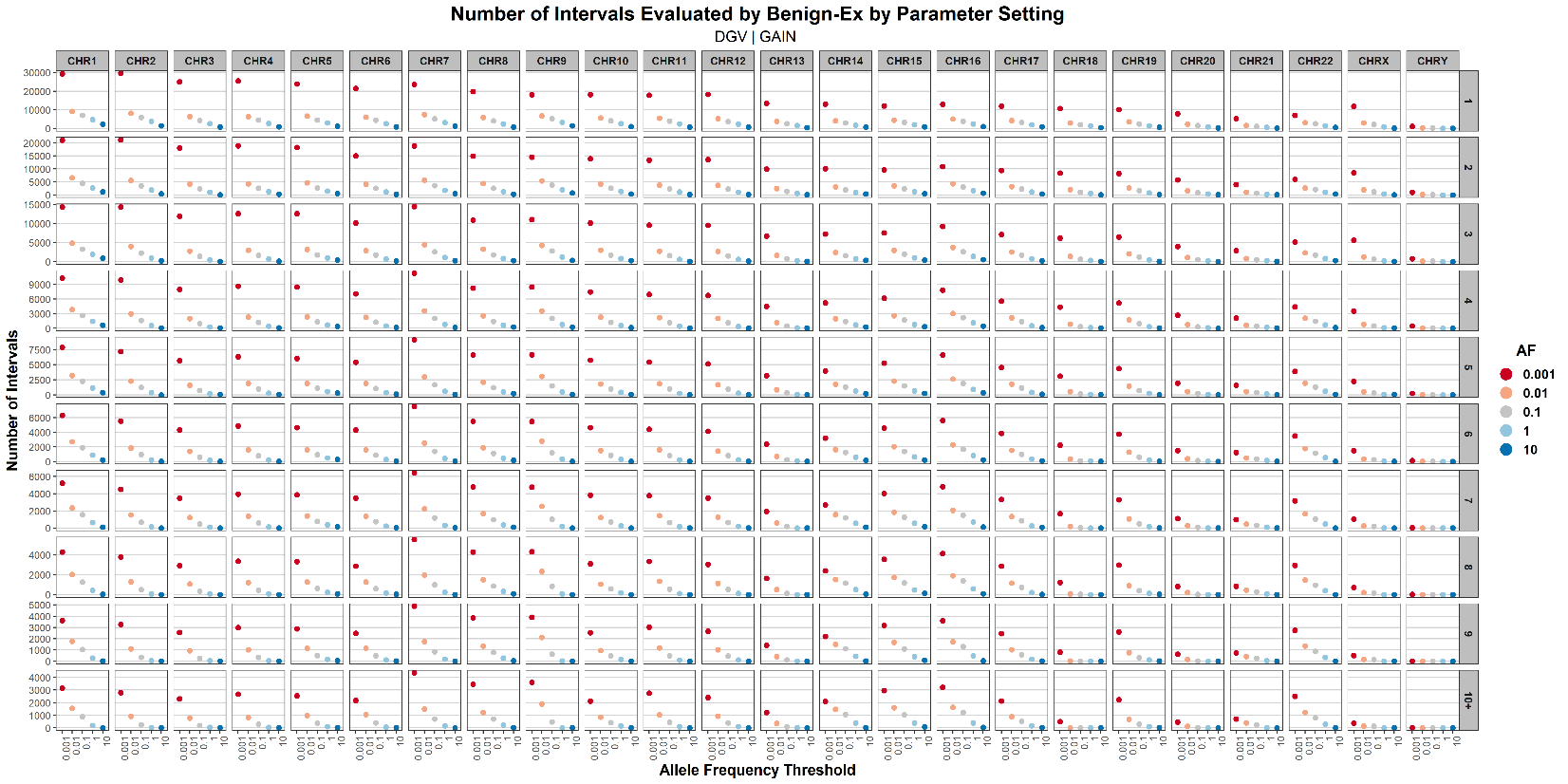

**Supplemental Figure 22: Percent Genome Called Benign by Benign-Ex by Parameter Setting for DGV**

**.** The percentage of the genome called benign for each parameter setting by Benign-Ex was tracked for the HG19 dataset. The minimum number of events is indicated on the top, the allele frequency on the bottom, and the method and year filters on the right. The top left box displays the percentage of the genome called benign for each allele frequency value with a minimum of 1 event and no method or year filter applied.

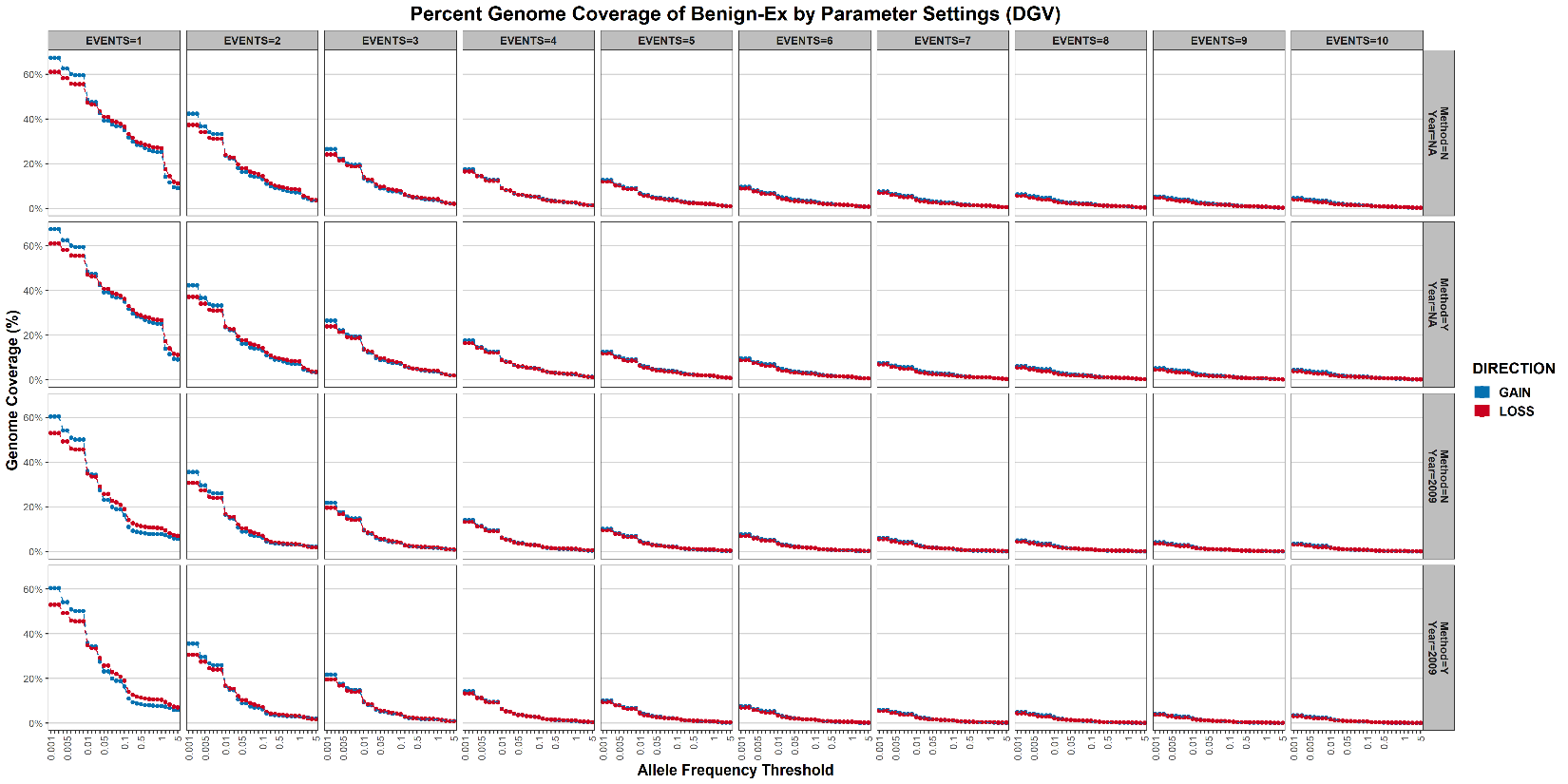

**Supplemental Figure 23: Percent Genome Called Benign by Benign-Ex by Parameter Setting for ClinGen**

**.** The percentage of the genome called benign for each parameter setting by Benign-Ex was tracked for the HG19 dataset. The minimum number of events is indicated on the bottom and the classification(s) on the top. The left box displays the percentage of the genome called benign for each minimum number of events wherein only benign variants were utilized.

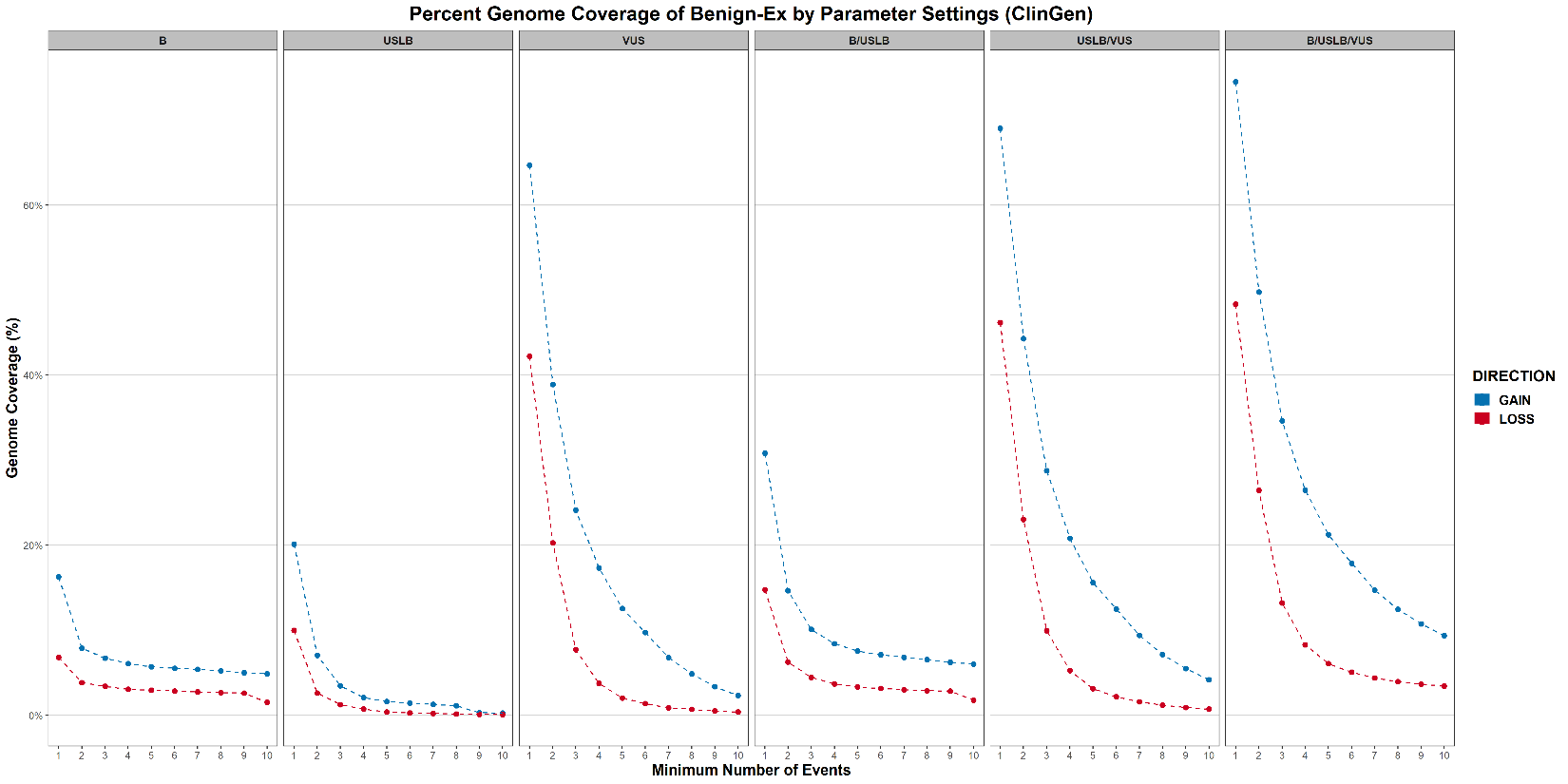

**Supplemental Figure 24: Distribution of Allele Frequencies within DGV (GRCh38)**

**.** Frequency distribution of the number of gain (blue) and loss (red) variants in DGV at each allele frequency threshold bin. The cumulative percentage of variants accounted for at each allele frequency threshold is also shown.

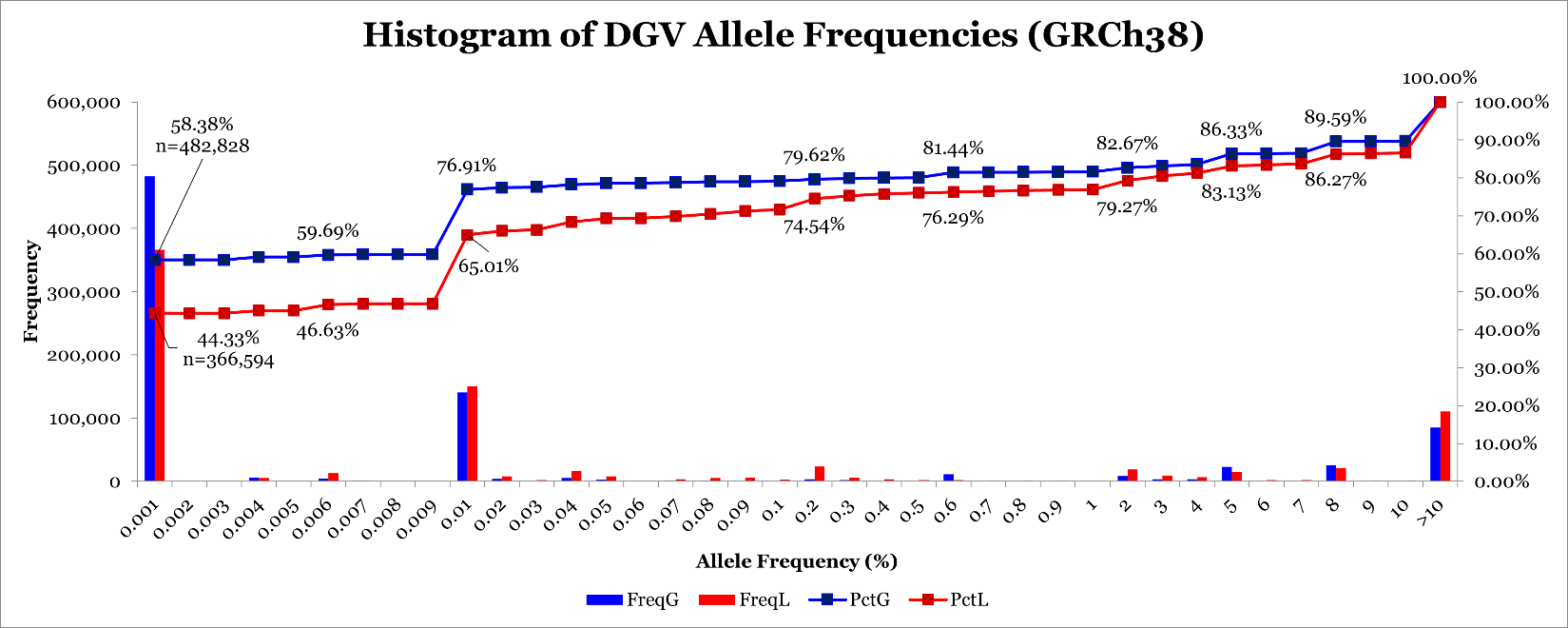

**Supplemental Figure 25: Distribution of Overlapping DGV Events: Tiled 500-bp Windows (GRCh38)**

**.** Frequency distribution of the number of gain (blue) or loss (red) DGV entries which overlap a single 500-bp window. The percentage of 500-bp windows which are retained at each threshold is shown.

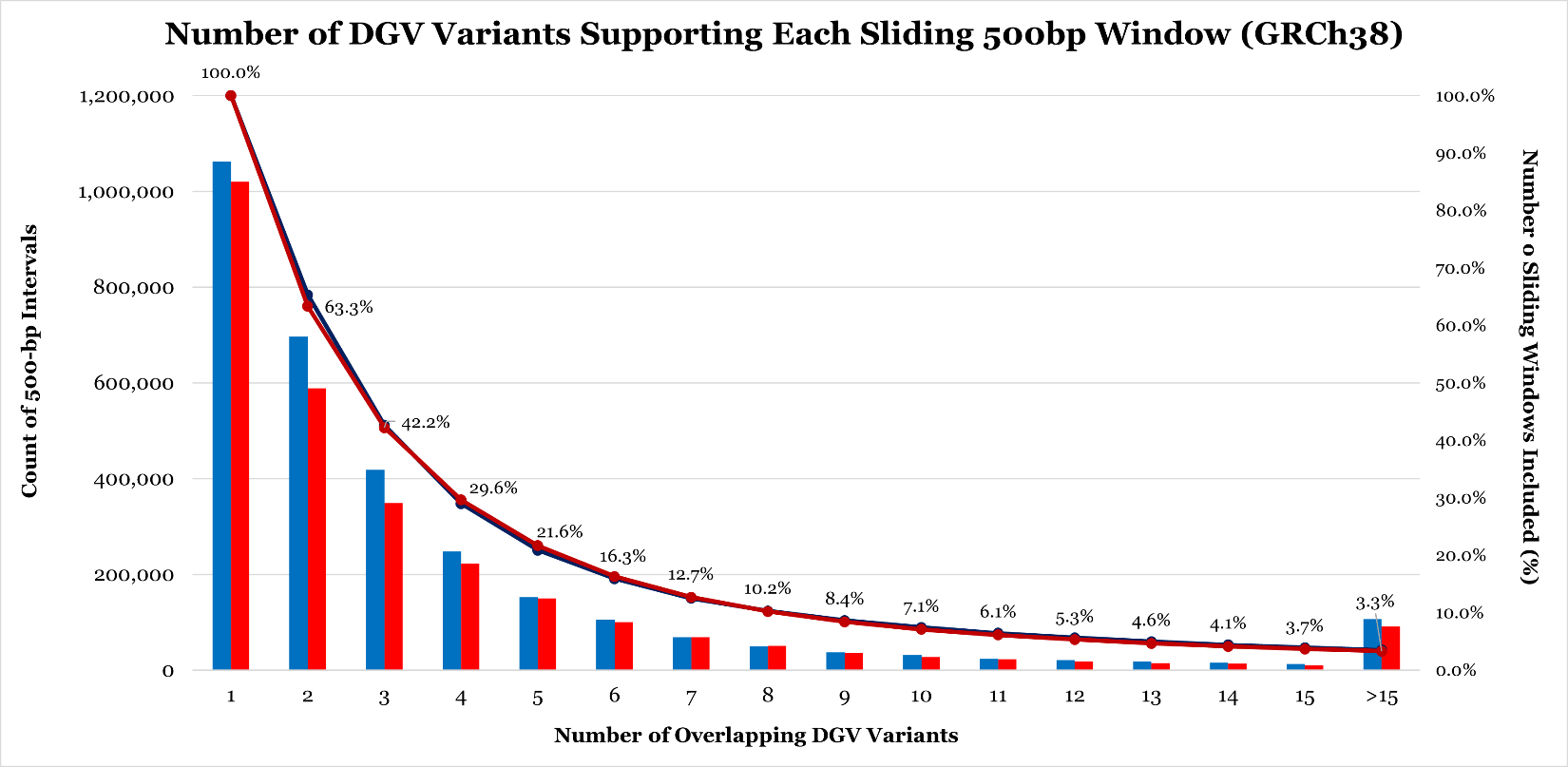

**Supplemental Figure 26: Final HG38 Benign-Ex Heatmaps for DGV**

**.** To visually demonstrate how different Benign-Ex parameter sets perform across multiple pathogenic lists, Benign-Ex produces the following heatmap. The histogram on the top represents the average performance of that parameter set across all pathogenic lists. The maximum value is the optimal parameter setting across all pathogenic lists. The histogram on the right represents the average performance of all Benign-Ex parameter sets across that pathogenic list. The highest performing parameter set is indicated for each pathogenic list. The heatmap in the middle plots the final summary statistic for each pathogenic list + Benign-Ex parameter set combination. The Benign-Ex parameter simulations are sorted first by the minimum number of events, then left to right by the combination of the year and methodology filter applications: (N/2009, N/NA, Y/2009, Y/NA). Finally, the simulations are sorted by allele frequency (AF) from smallest to largest. Red indicates negative scores, Blue indicates positive scores, and White indicates a ‘0’ score. (A) Final 576 DGV Gain Simulations, (B) DGV Gain Simulations for Method=N; Year=NA (C) DGV Gain Simulations for Method=N; Year=2009, (D) DGV Gain Simulations for Method=Y; Year=NA; (E) DGV Gain Simulations for Method=Y; Year=2009; (F) Final 576 DGV Loss Simulations, (G) DGV Loss Simulations for Method=N; Year=NA (H) DGV Loss Simulations for Method=N; Year=2009, (I) DGV Loss Simulations for Method=Y; Year=NA; (J) DGV Loss Simulations for Method=Y; Year=2009.

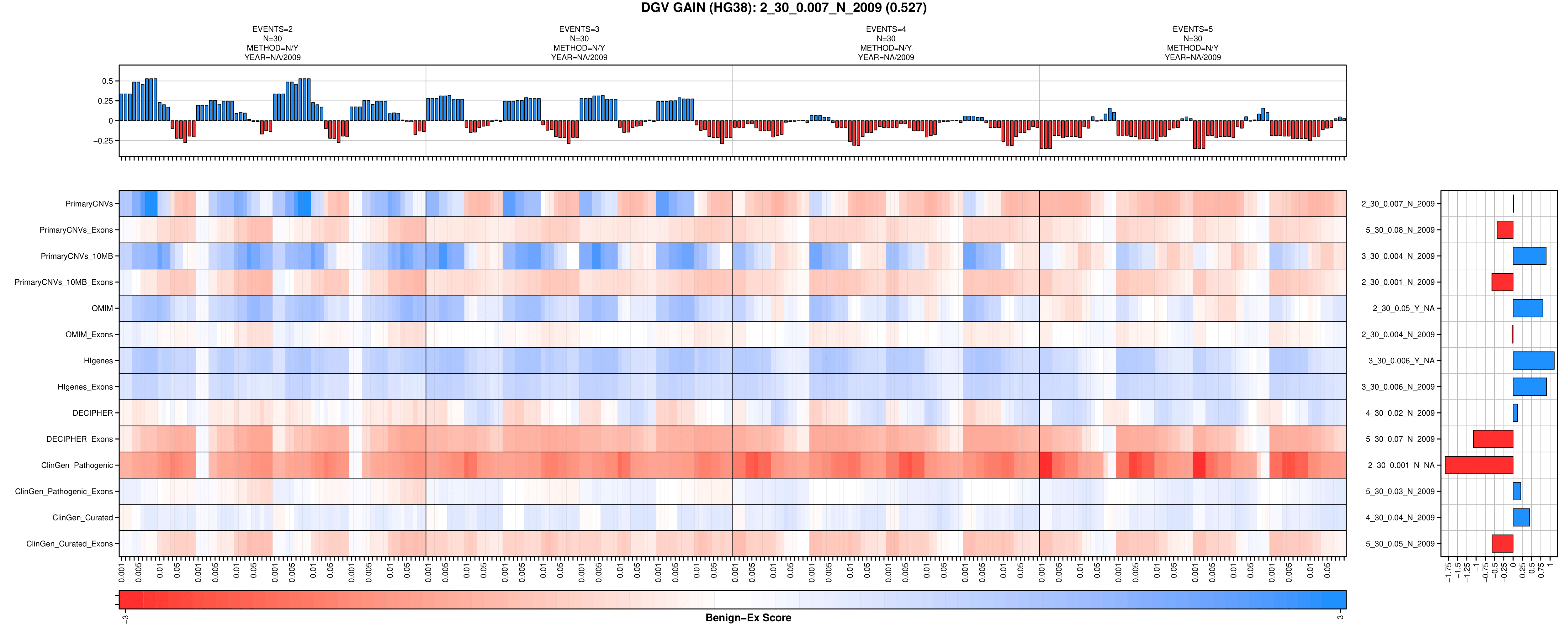

A

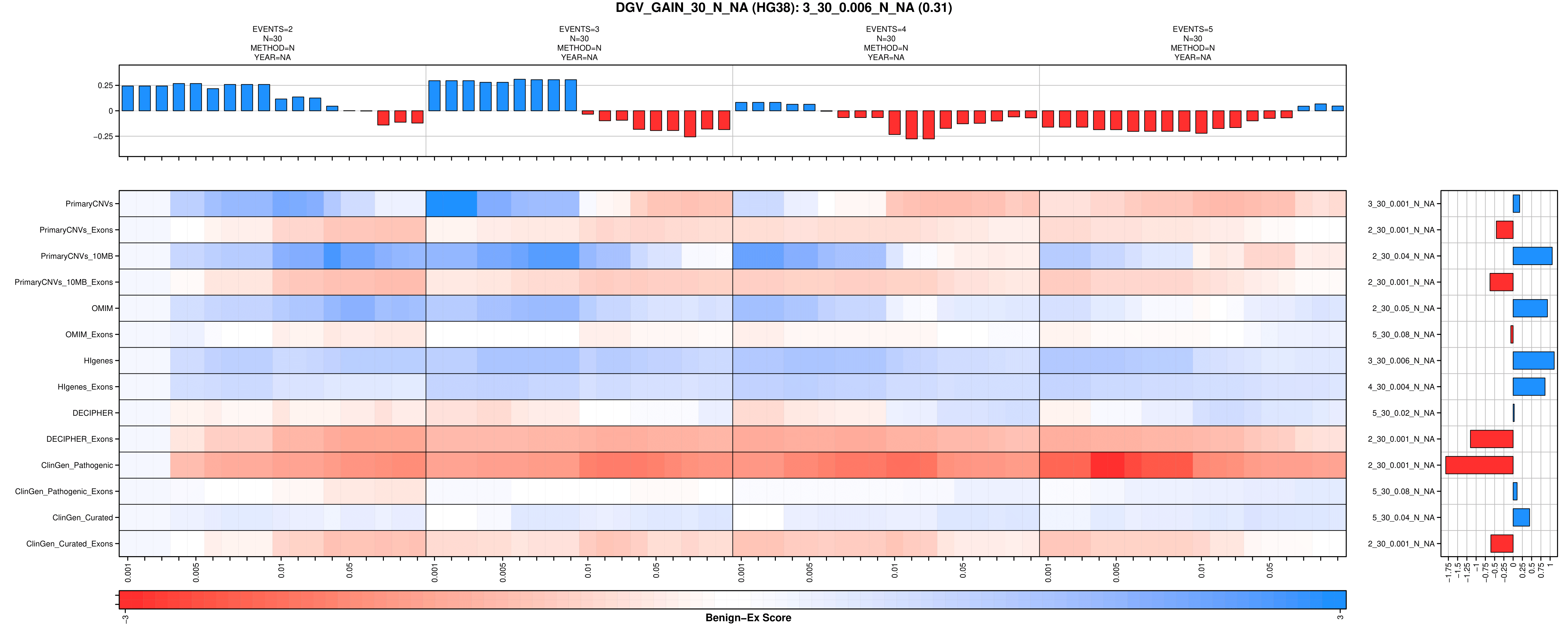

B

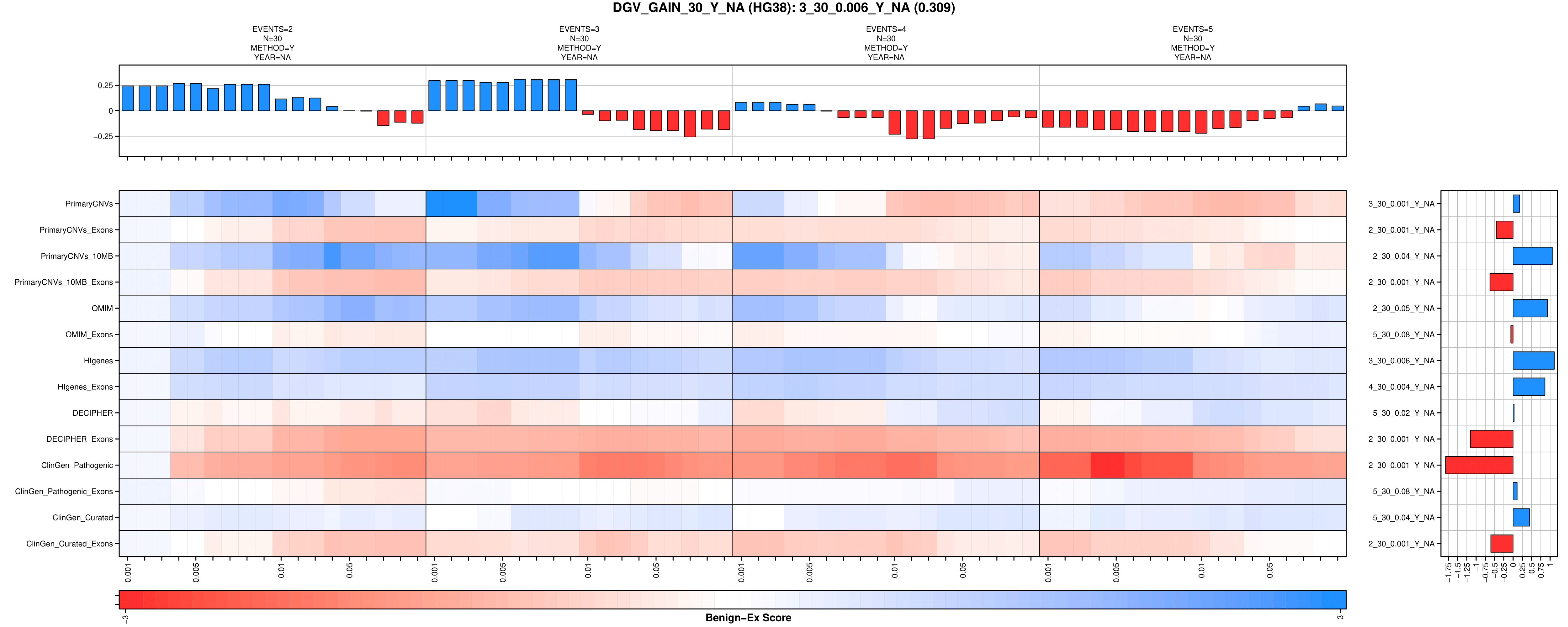

C

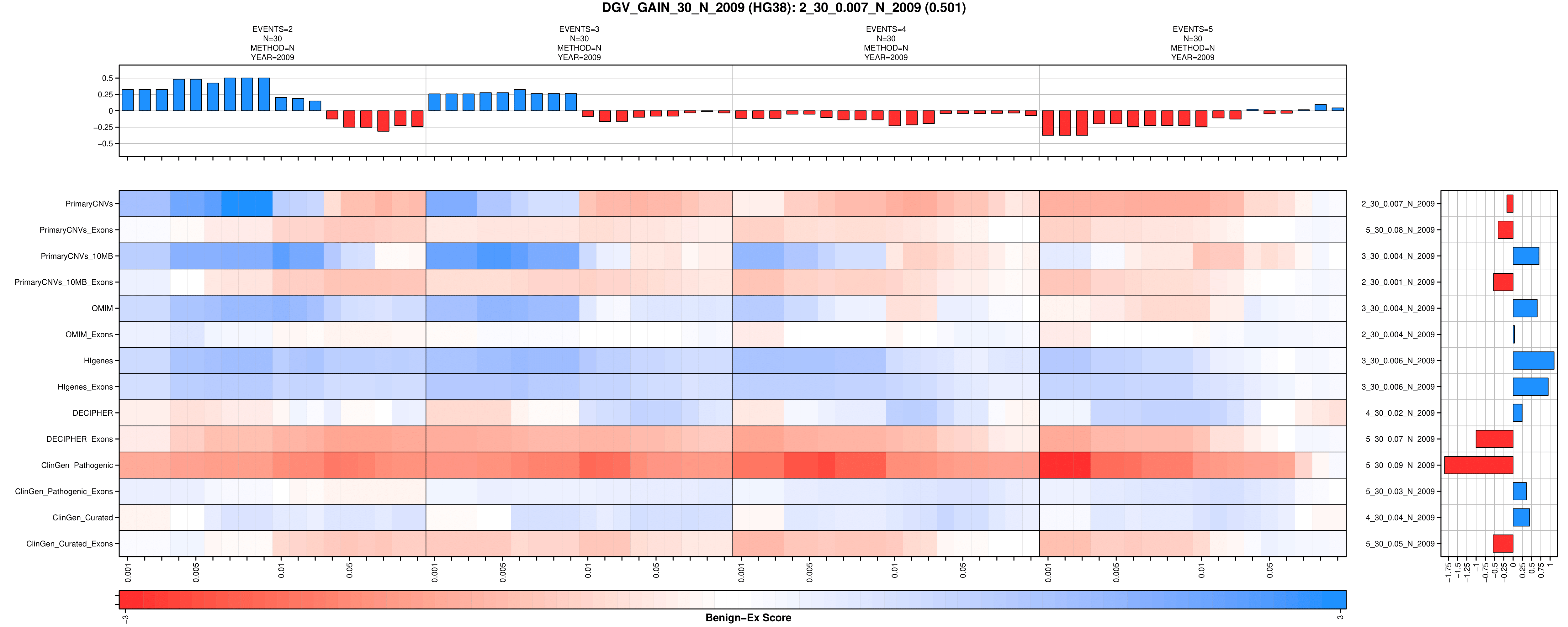

D

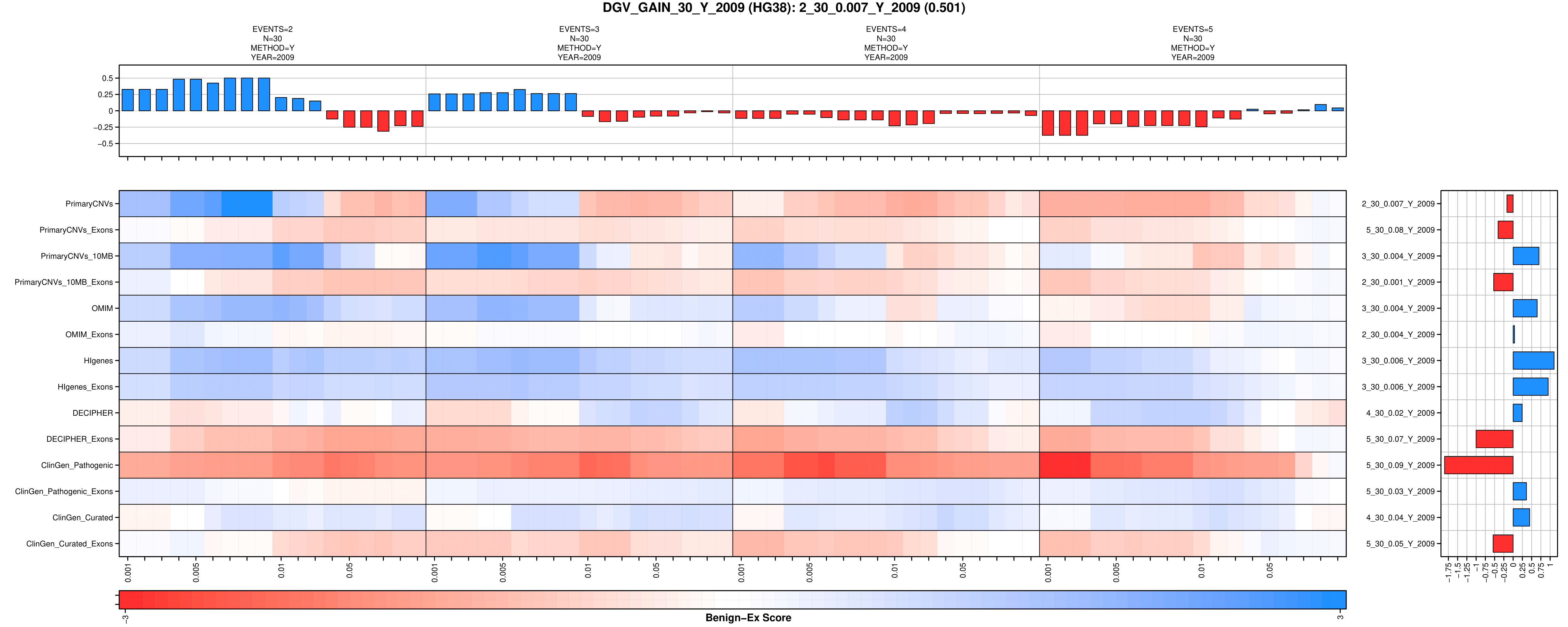

E

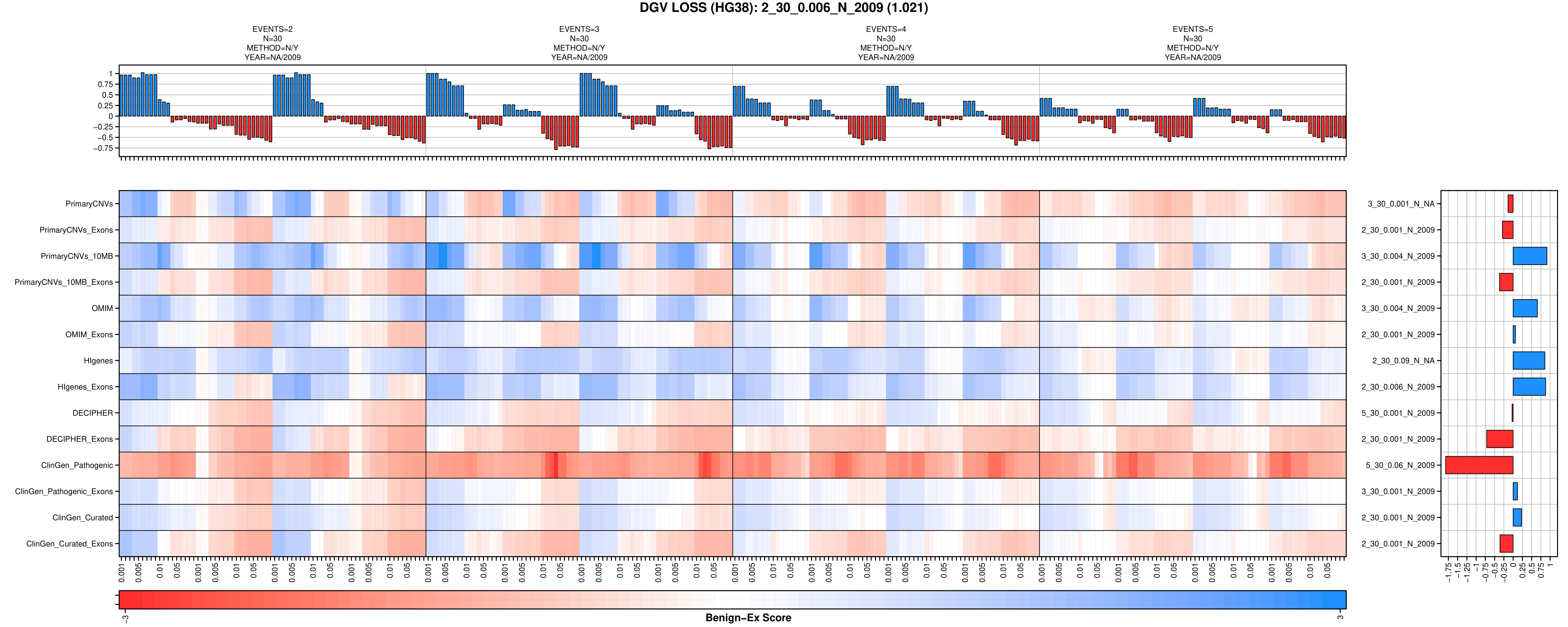

F

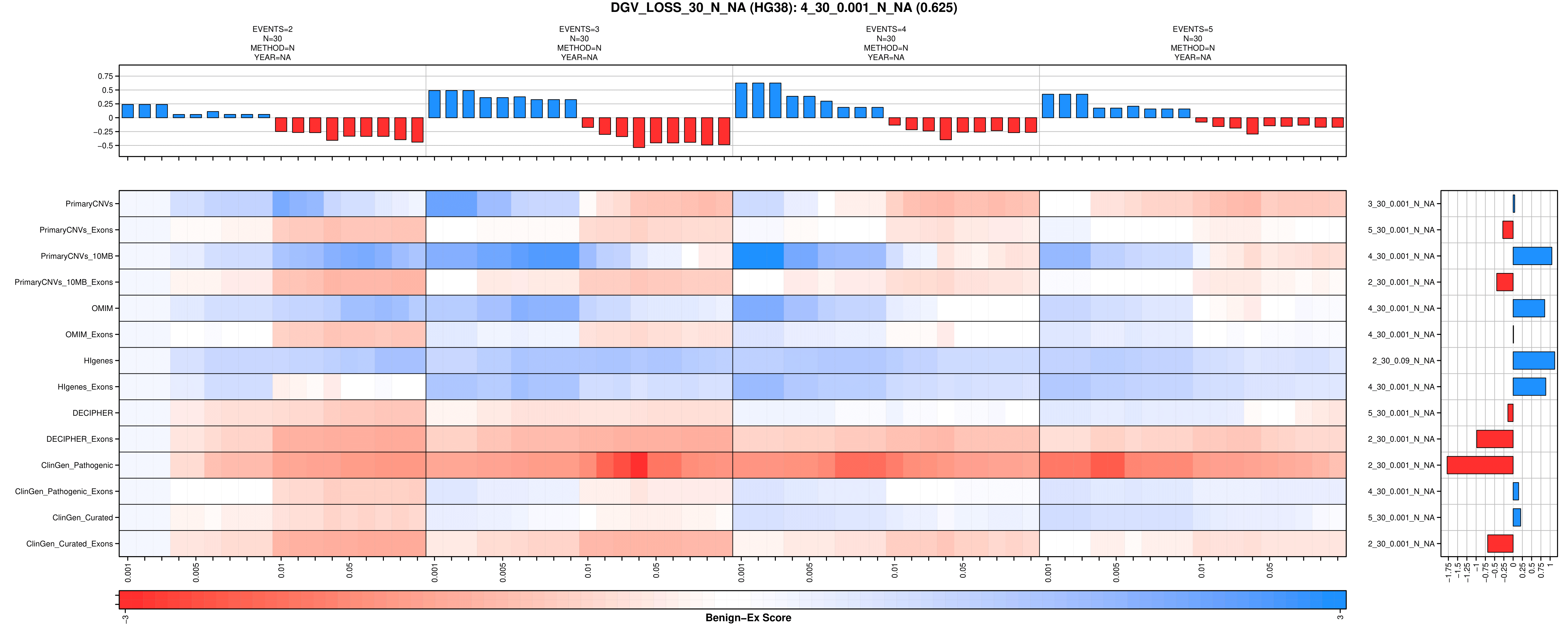

G

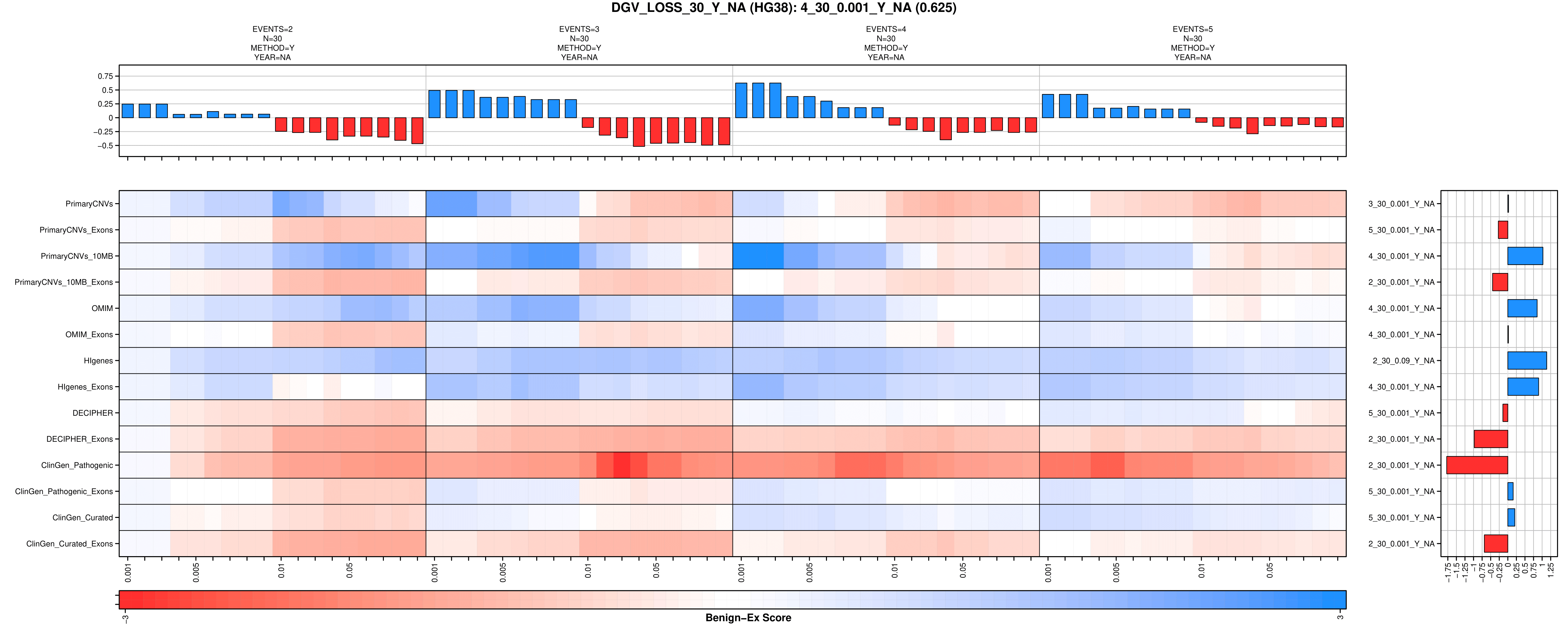

H

I

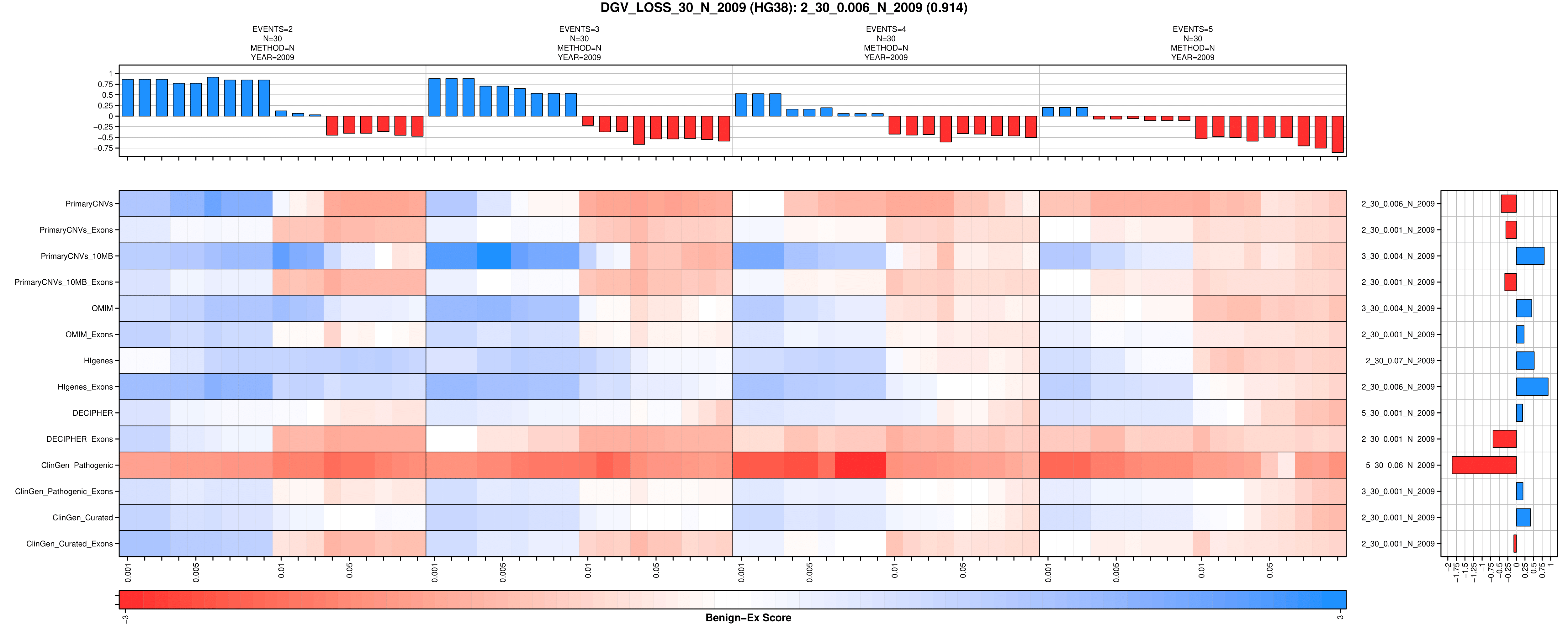

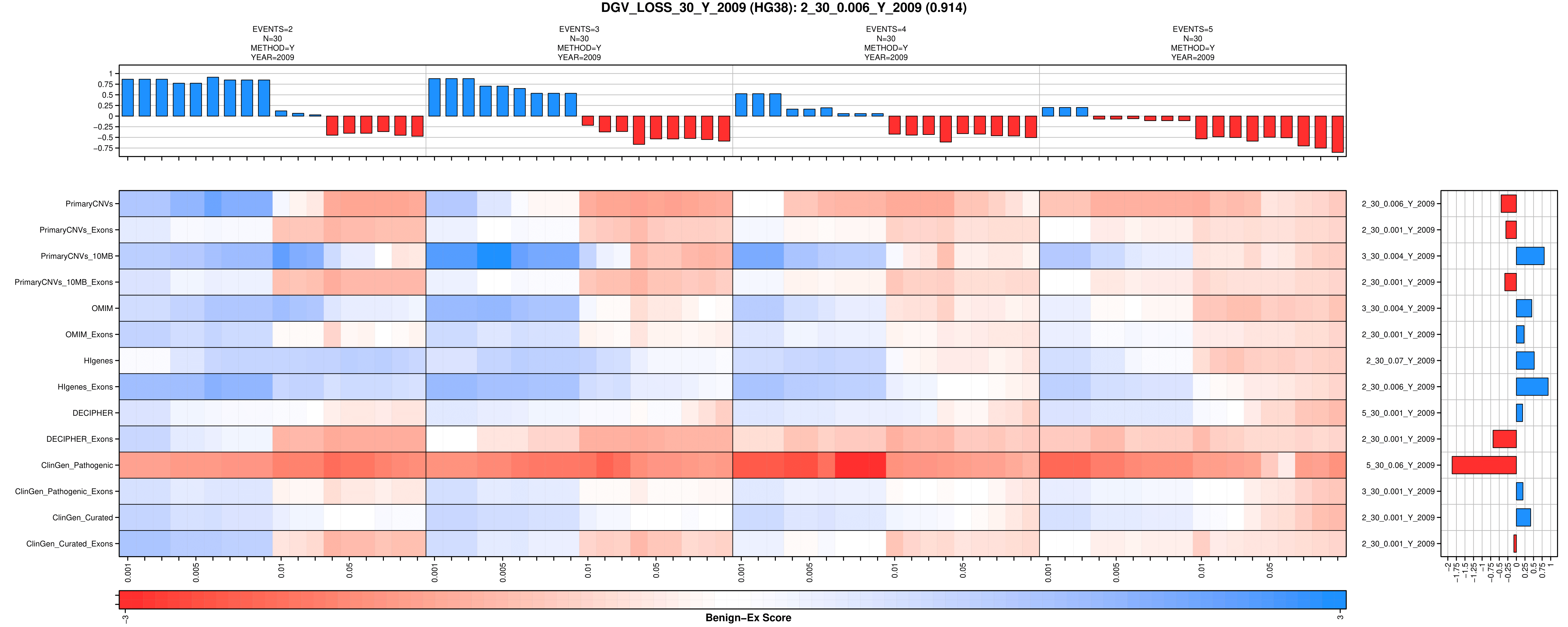

J

**Supplemental Figure 27: Final HG38 Benign-Ex Heatmaps for ClinGen**

**.** To visually demonstrate how different Benign-Ex parameter sets perform across multiple pathogenic lists, Benign-Ex produces the following heatmap. The histogram on the top represents the average performance of that parameter set across all pathogenic lists. The maximum value is the optimal parameter setting across all pathogenic lists. The histogram on the right represents the average performance of all Benign-Ex parameter sets across that pathogenic list. The highest performing parameter set is indicated for each pathogenic list. The heatmap in the middle plots the final summary statistic for each pathogenic list + Benign-Ex parameter set combination. The Benign-Ex parameter simulations are sorted by the acceptable classifications (‘B’, ‘B/USLB’, ‘B/USLB/VUS’). Red indicates negative scores, Blue indicates positive scores, and White indicates a ‘0’ score. (A) Final 30 ClinGen Gain Simulations, (B) All 30 ClinGen Loss Gain Simulations.

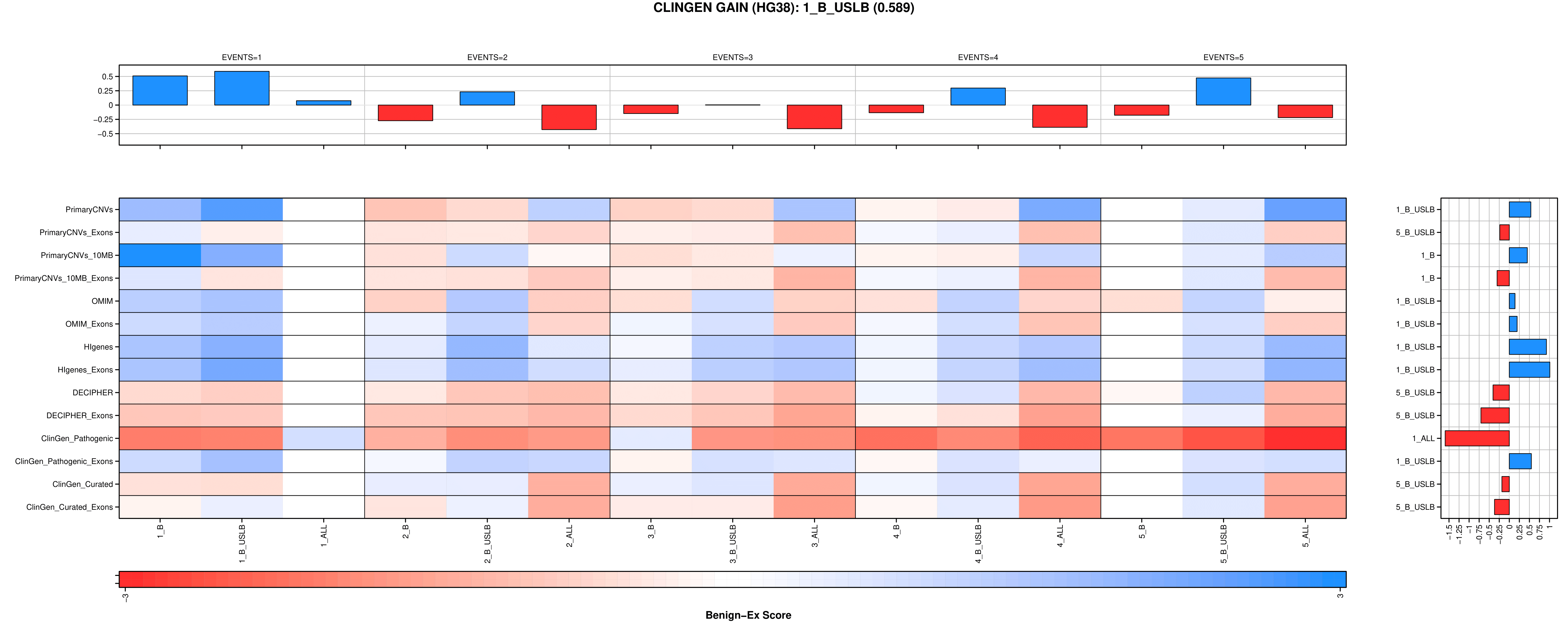

A

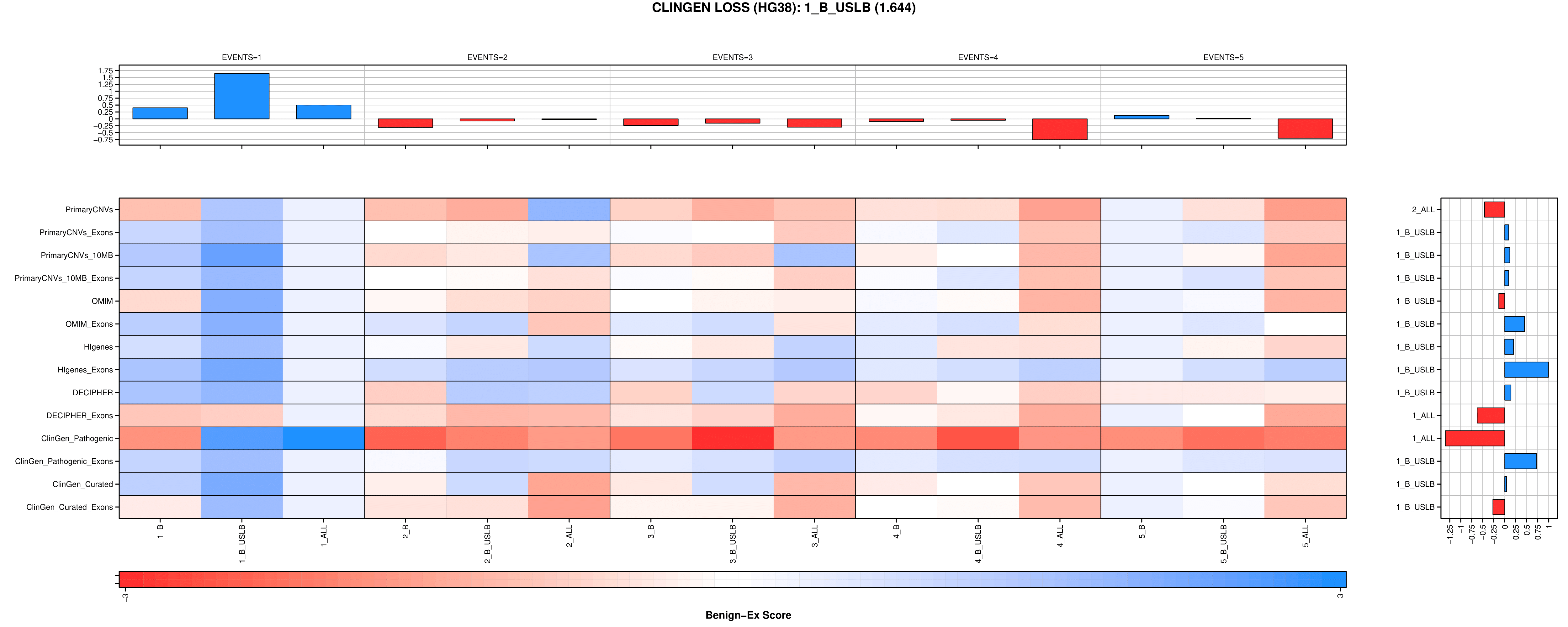

B

**Supplemental Figure 28: Distribution of Reported Ethnicities by Continent**

**.**

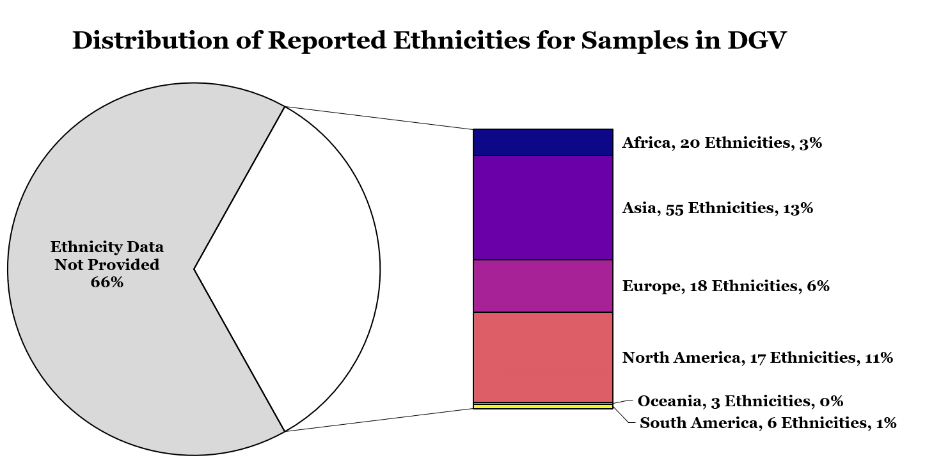

**Supplemental Figure 29: Map of Reported Ethnicities in DGV**

**.** World map depicts each of the 114 different ethnicities sampled from 54 different countries. Countries are color-coded by continent.
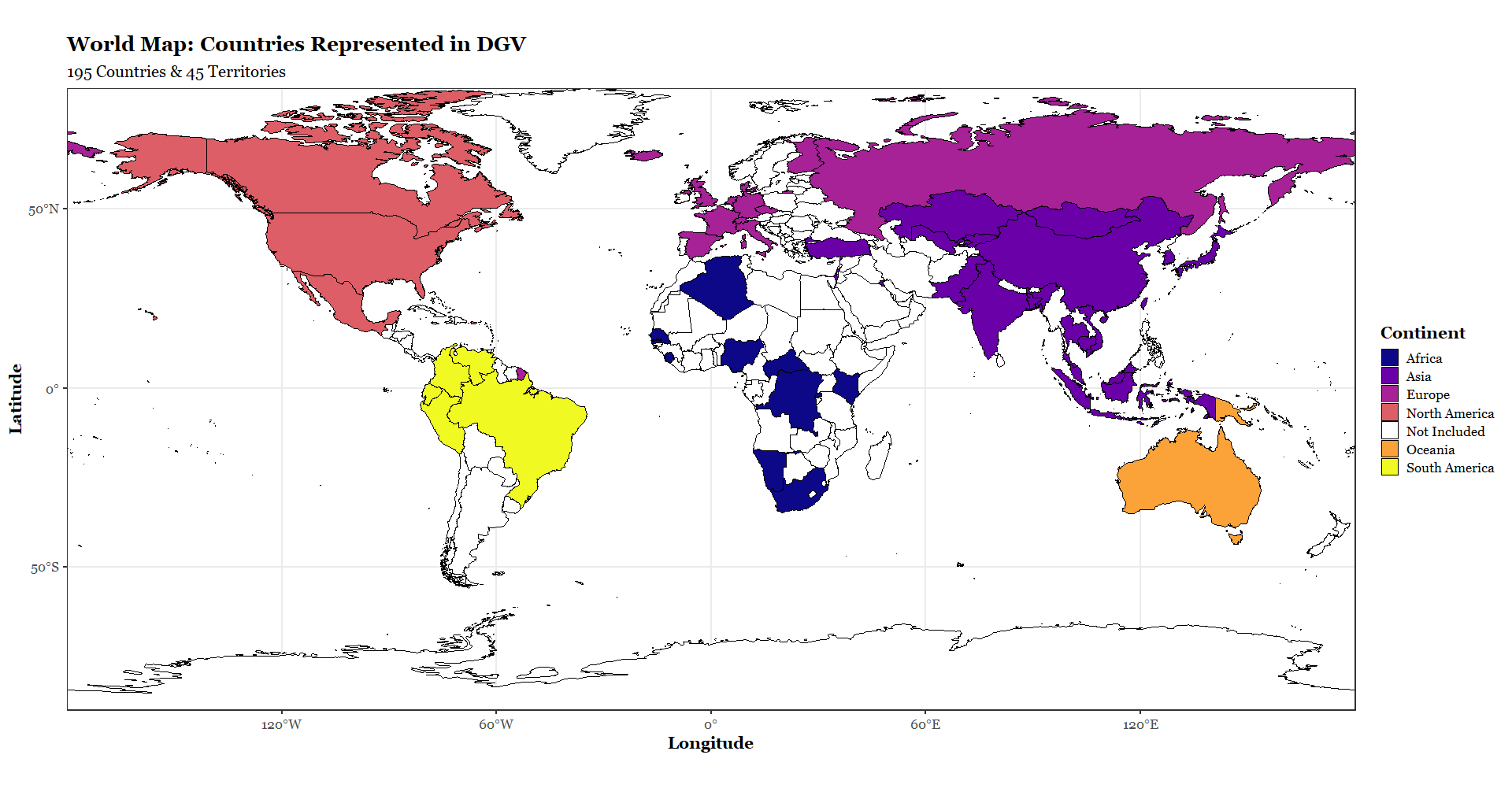

**North America:**

Admixed, Africa, African American, African Caribbean, Ashkenazi Jewish, Asian, Asian American, Caucasian, Chinese, French, Gujarati Indian, Han Chinese, Mayan, Mexican, Pima, Puerto Rican, Unspecified Native American

**South America**

Colombian, Huaorani, Karitiana, Peruvian, Surui, Venezuelan

**Africa:**

African, Bantu, Biaka Pygmies, Esan, Gambian, Herero, Luhya, Maasai, Mandenka, Mbuti Pygmy, Mende, Mozabite, Ovambo, Pedi, Rwandese, San, Sotho, Tswana, Yoruba, Zulu

**Europe:**

Basque, British, Caucasian, Czech, Danish, Dutch, European, Finnish, French, Iberian, Icelandic, Indian Telugu, Italian, Orcadian, Sardinian, Sri Lankan Tamil, Swiss, Toscani, Tuscan

**China:**

Chinese, Dai, Daur, Han, Hezhen, Hui, Lahu, Miao, Mongola, Naxi, Oroqen, She, Southern Han, Tibetan, Tu, Tujia, Uyghur, Xibe, Xibo, Yizu

**Asia:**

Ami, Atayal, Balochi, Bedouin, Bengali, Brahui, Burusho, Cambodian, Druze, Hazara, Indian, Japanese, Kalash, Kazakh, Khirghiz, Kinh, Korean, Makrani, Malay, Mongolian, Negev, Negrito, Palestinian, Pathan, Persian,Punjabi, Saudi, Sindhi, Thai, Turkish, Uzbek

**Oceania:**

Caucasian, Melanesian, Papuan

**Russia:**

Adygei, Russian, Tatar, Yakut

.

Literature Cited

1.	Rehm, H.L. *et al.* ClinGen--the Clinical Genome Resource. *N Engl J Med* **372**, 2235-42 (2015).

2.	Karolchik, D. *et al.* The UCSC Table Browser data retrieval tool. *Nucleic Acids Res* **32**, D493-6 (2004).

3.	Kent, W.J. *et al.* The human genome browser at UCSC. *Genome Res* **12**, 996-1006 (2002).

4.	Firth, H.V. *et al.* DECIPHER: Database of Chromosomal Imbalance and Phenotype in Humans Using Ensembl Resources. *American journal of human genetics* **84**, 524-533 (2009).

5.	Petrovski, S., Wang, Q., Heinzen, E.L., Allen, A.S. & Goldstein, D.B. Genic Intolerance to Functional Variation and the Interpretation of Personal Genomes. *PLOS Genetics* **9**, e1003709 (2013).

6.	Online Mendelian Inheritance in Man, OMIM®. (McKusick-Nathans Institute of Genetic Medicine, Johns Hopkins University, Baltimore, MD).

7.	Hinrichs, A.S. *et al.* The UCSC Genome Browser Database: update 2006. *Nucleic acids research* **34**, D590-D598 (2006).

8.	Peterson, R.A. & Cavanaugh, J.E. Ordered quantile normalization: a semiparametric transformation built for the cross-validation era. *Journal of Applied Statistics* **47**, 2312-2327 (2020).

9.	McCarroll, S.A. *et al.* Integrated detection and population-genetic analysis of SNPs and copy number variation. *Nature Genetics* **40**, 1166-1174 (2008).

10.	Perry, G.H. *et al.* The fine-scale and complex architecture of human copy-number variation. *Am J Hum Genet* **82**, 685-95 (2008).

11.	Favorov, A. *et al.* Exploring Massive, Genome Scale Datasets with the GenometriCorr Package. *PLOS Computational Biology* **8**, e1002529 (2012).

12.	Quinlan, A.R. & Hall, I.M. BEDTools: a flexible suite of utilities for comparing genomic features. *Bioinformatics* **26**, 841-842 (2010).

13.	Wetzel, A.S. & Darbro, B.W. A Comprehensive List of Human Microdeletion and Microduplication Syndromes. (2022).

14.	MacDonald, J.R., Ziman, R., Yuen, R.K., Feuk, L. & Scherer, S.W. The Database of Genomic Variants: a curated collection of structural variation in the human genome. *Nucleic Acids Res* **42**, D986-92 (2014).

15.	Lappalainen, I. *et al.* DbVar and DGVa: public archives for genomic structural variation. *Nucleic Acids Res* **41**, D936-41 (2013).

16.	Collins, R.L. *et al.* A structural variation reference for medical and population genetics. *Nature* **581**, 444-451 (2020).

17.	Iafrate, A.J. *et al.* Detection of large-scale variation in the human genome. *Nature Genetics* **36**, 949-951 (2004).

18.	Cooper, G.M. *et al.* A copy number variation morbidity map of developmental delay. *Nature Genetics* **43**, 838-846 (2011).

19.	Coe, B.P. *et al.* Refining analyses of copy number variation identifies specific genes associated with developmental delay. *Nat Genet* **46**, 1063-71 (2014).

20.	Frazer, K.A. *et al.* A second generation human haplotype map of over 3.1 million SNPs. *Nature* **449**, 851-861 (2007).

21.	The International HapMap Project. *Nature* **426**, 789-96 (2003).

22.	Auton, A. *et al.* A global reference for human genetic variation. *Nature* **526**, 68-74 (2015).

23.	Cavalli-Sforza, L.L. The Human Genome Diversity Project: past, present and future. *Nature Reviews Genetics* **6**, 333-340 (2005).

24.	Zarrei, M., MacDonald, J.R., Merico, D. & Scherer, S.W. A copy number variation map of the human genome. *Nat Rev Genet* **16**, 172-83 (2015).
